# Cognitive and executive impairments in Parkinson’s Disease psychosis: a Bayesian meta-analysis

**DOI:** 10.1101/2022.12.24.22283774

**Authors:** Sara Pisani, Luca Gosse, Rita Wieretilo, Dominic ffytche, Latha Velayudhan, Sagnik Bhattacharyya

**Affiliations:** Division of Academic Psychiatry, Department of Psychosis, Institute of Psychiatry, Psychology and Neuroscience, King’s College London, London, SE5 8AF, United Kingdom; Department of Psychology, Institute of Psychiatry, Psychology and Neuroscience, King’s College London, London, SE5 8AF, United Kingdom; Faculty of Medicine, Dentistry and Health, Medical School, University of Sheffield, Sheffield, S10 2RX, United Kingdom; Department of Old Age Psychiatry, Division of Academic Psychiatry, Institute of Psychiatry, Psychology and Neuroscience, King’s College London, London, SE5 8AF, United Kingdom

**Keywords:** Parkinson’s disease, psychosis, cognition, executive functions, Bayesian meta-analysis

## Abstract

**Background:** Cognitive and executive deficits lead to worsening of quality of life and are a risk factor for developing dementia in Parkinson’s Disease (PD) patients with psychosis. However, which key cognitive domains are differentially affected in PD patients with psychosis (PDP) compared to those without (PDnP), remains unclear. Here we examined this using a Bayesian meta-analytic approach.

**Methods:** Searches were conducted on PubMed, Web of Science, SCOPUS, Medline and PsycINFO. Hedges’ g effect-size estimates were extracted from eligible studies as a measure of standard mean differences between PDP and PDnP patients. Meta-analyses were conducted separately for each cognitive domain and sub-domain, we examined the effect of age, PD medications, PD duration and severity, depression and psychosis severity for all major domains with meta-regressions.

**Results:** Effect-size estimates suggest worse performance on all major domains (k=105 studies) in PDP compared to PDnP patients, with global cognition (k=103 studies, *g*=-0.57), processing speed (k=29 studies, *g*=-0.58), executive functions (k=33, *g*=-0.56), episodic memory (k=30 studies, *g*=-0.58), and perception (k=34 studies, *g*=-0.55) as the most likely affected domains. Age, depression, and PD duration had moderating effects on task-related performance across most of the major nine domains.

**Conclusions:** We report extensive deficits across nine domains as well as sub-domains in PD psychosis, with global cognition, processing speed and executive functions as the most likely impaired. Presence of depression may influence task-related performance in PDP, alongside age and PD duration, but not dose of dopamine replacement treatments.

## Introduction

Psychotic symptoms in Parkinson’s Disease (PD) are one of the most common non-motor symptoms, manifesting typically as visual hallucinations and delusions^1^, are distressing and associated with increased risk of hospitalisation, loss of independence and mortality^2-2^. Several studies have shown that patients with PD psychosis (PDP) have worse performance compared to PD patients without psychosis in cognitive domains such as memory ^5^, attention, visuo-spatial abilities and other executive skills^6, 7^. Cognitive deficits are associated with poor quality of life, and with the onset of dementia as PD progresses^8^. However, which key cognitive domains are differentially affected in PD patients with psychosis compared to those without, remains unclear. Montagnese et al.^9^ investigated this by taking a meta-analytic approach and found that across measures of episodic memory, global cognition (e.g., Mini-Mental State Examination, MMSE^10^), language, attention, visual perception, working memory and executive functioning, PD patients with visual hallucinations performed worse compared to PD patients without visual hallucination, with the greatest impairments being in executive functions and attention. They also reported that only age, but not PD medications, duration and severity of PD, gender or general cognitive status, was associated with performance on general cognitive tests, working memory and executive functions^9^.

However, which cognitive domains might be most likely to be differentially impaired in PD patients with psychosis compared to those without psychosis remains unclear. This is important for a better understanding of the relationship between neurocognitive impairments and psychotic symptoms, and their potential role in the development of these symptoms, which in turn may help develop more effective interventions. Hence, we wanted to address these issues using two approaches. Instead of taking a frequentist approach such as that adopted by Montagnese et al. ^9^, we have adopted a Bayesian approach to allow us to estimate the likelihood or probability of impairment in task performance above a certain threshold in each cognitive domain between PDP patients. Further, we compared the magnitude of impairment in all the domains by restricting our analyses to those studies that have reported across all cognitive domains of interest to identify domains for which the evidence of impairment is most robust. Unlike before, we also examined impairment in sub-domains of broader cognitive domains, for example working memory sub-tests measuring manipulation and maintenance of information, category- or letter-based fluency, the ability of copying and drawing, verbal and non-verbal retrieval ^11-11^. We also updated the evidence from Montagnese et al. ^9^ with additional studies including PDP patients with a wider range of psychotic manifestations, e.g., multimodal hallucinations, hallucinations with/without insights and more severe symptoms (e.g., delusions), and summarised evidence on different cognitive domains based on recommended approaches ^14^. Finally, we explored the association between age, PD medications, depression, and severity of psychosis symptoms on the probability of reporting such task-related performance differences using a Bayesian meta-regression and also report pooled effect-size estimates after accounting for the effect of confounding variables that were significantly associated with task performance.

## Methods

### Search strategy

Searches were conducted on PubMed, Web of Science, SCOPUS, Medline and PsycINFO (Ovid) on 14^th^ September 2021 (protocol registered on PROSPERO; CRD42021260475). Full search strategy can be found in Supplementary Material 1. In brief, terms for the clinical condition (e.g., “Parkinson*”, “Parkinson psychosis”, “Parkinson disease psychosis”) and for cognitive deficits (e.g., “memory impair*”, “attention*”, “language impair*”) were used. Systematic review and meta-analysis were used as additional resources to identify articles. We report following the Preferred Reporting Items for Systematic Reviews and Meta-analyses (PRISMA) guidelines^15^.

### Eligibility criteria

Studies were included in this review if a) they included PD psychosis patients and compared their performance on neuropsychological assessments with that of PD patients without psychosis (PDnP); b) patients with PD had a diagnosis of psychosis after PD onset; c) PD patients did not have a diagnosis of PD dementia; d) they were written in English, peer-reviewed and original pieces of research; and e) if they had available data on the neuropsychological assessments for each group. Studies that examined cognitive deficits in PD patients were included on the grounds that they provided subgroup analyses between PD psychosis and PD patients without psychosis. Grey literature, reviews, books, and conference abstracts were excluded. We did not include studies with PD dementia patients, and longitudinal studies were included on the grounds that they provided baseline assessments for use in the present analyses.

### Data synthesis

Data extraction procedure is reported in Supplementary Material 1. The nature of the cognitive assessments examined in this review measured a variety of cognitive and executive areas and were classified into separate domains as recommended ^14^. We conducted separate meta-analyses for each category and sub-category of cognitive and executive domains. First, we extracted Hedges’ *g* effect-size estimates and associated standard error for each cognitive task for each study representing standardised mean differences between PD psychosis and PDnP. These were computed in R (version 4.0.3)^16^ using the *metafor* package ^17^. Subsequently, Hedges’ *g* effect sizes from individual studies were synthesized in a Bayesian meta-analysis using a random-effects approach with the *brms* package ^18^ based on the STAN probabilistic programming language ^19^, accounting for the fact that some studies contributed to multiple domains or sub-domains, thus violating the requirement of independence of effect sizes ^20^. The package *brms* provides an estimate of between- and within-study variance which assesses the presence of variability across different studies. We applied a normal distribution for the population-level parameter (mean=0, SD=1)^21^, with a half-*t* distribution as τ, known as Half Cauchy, as recommended ^22^ and also the Hamiltonian Monte Carlo No-U-Turn sampling procedure^23, 24^. Several cognitive tasks involve multiple domains. For example, letter-based or category-based fluency tests require processing speed as well as language ability; similarly, the Stroop tests ^25, 26^ calls upon both selective attention and it requires the ability to switch from one task instruction to a different one, and processing information in a timely manner. When such tests were employed in a study, they were included under all the applicable domains ^14^. Perception-based assessments were classified into sub-domains based on the work from Montagnese et al.^9^. Publication bias was assessed by regressing the effect-size standard error on the effect sizes for each analysis, and we applied cluster-robust variance estimation to control for the lack of independent effect size ^27, 28^.

In the first instance, separate Bayesian analyses were conducted for each domain. In order to identify the cognitive domains most likely affected in PDP, we estimated the empirical cumulative distribution function (ECDF) for each domain using a threshold of effect-size = -0.3. A negative effect-size indicated that PDP patients performed worse compared to PDnP patients. Using this threshold, the ECDF gives the probability of impairment (i.e., PDP worse than PDnP) equivalent to an effect-size difference of 0.3 or more. We reasoned that any cognitive domains affected to an extent equal to or greater than this threshold were most likely to represent the key domains that may underlie psychosis in PD, as opposed to general cognitive impairments associated with PD itself. Finally, in order to allow a systematic comparison of the magnitude of impairment across all the cognitive domains investigated thus far in the literature, we also carried out a separate analysis restricting to those studies that have reported across all cognitive domains of interest. Below, effect sizes are reported in Hedges’ *g*, as estimate of standard mean difference between PDP and PDnP patients, and 95% credible intervals (95% CI). In all analyses, convergence was successful as represented by the potential scale reduction factor 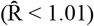.

Bayesian meta-regressions were conducted where at least 10 or more studies were available to examine the effect of key confounding factors of interest such as depression, dopamine-replacement medications (expressed in Levodopa equivalent daily dose, LEDD), motor symptoms (expressed with the Unified Parkinson’s Disease Rating Scale (MDS-UPDRS) part III scores, as a measure of PD severity), and severity of psychosis symptoms on the performance on neuropsychological assessments. To summarise neuropsychiatric scores for depression and psychosis severity from different assessments, we computed standardised scores by dividing the mean with the standard deviation reported in each study for each patient’s group respectively for depression and psychosis severity. We assessed the quality of included studies using the Effective Public Health Practice Project (EPHPP) ^29^ scale (see Supplementary Material 1 for description).

## Results

### Study characteristics

From an initial list of 5144 studies, we identified a total of 69 studies after removing duplicates and full-text screening, and a further 36 from systematic reviews and meta-analysis ^9^ leading to a total of 105 studies reporting on 2912 PDP patients (mean age ± SD = 68.92 ± 4.34, 48.8% male) and 6056 PDnP patients (mean age ± SD = 66.88 ± 4.28, 57.1% male). Basic sample characteristics are reported in Table 1 (for description of sample feature please see Supplementary Materials 2-3). Figure 1 reports the flowchart with the steps that led to the final study pool.

**Table 1.**
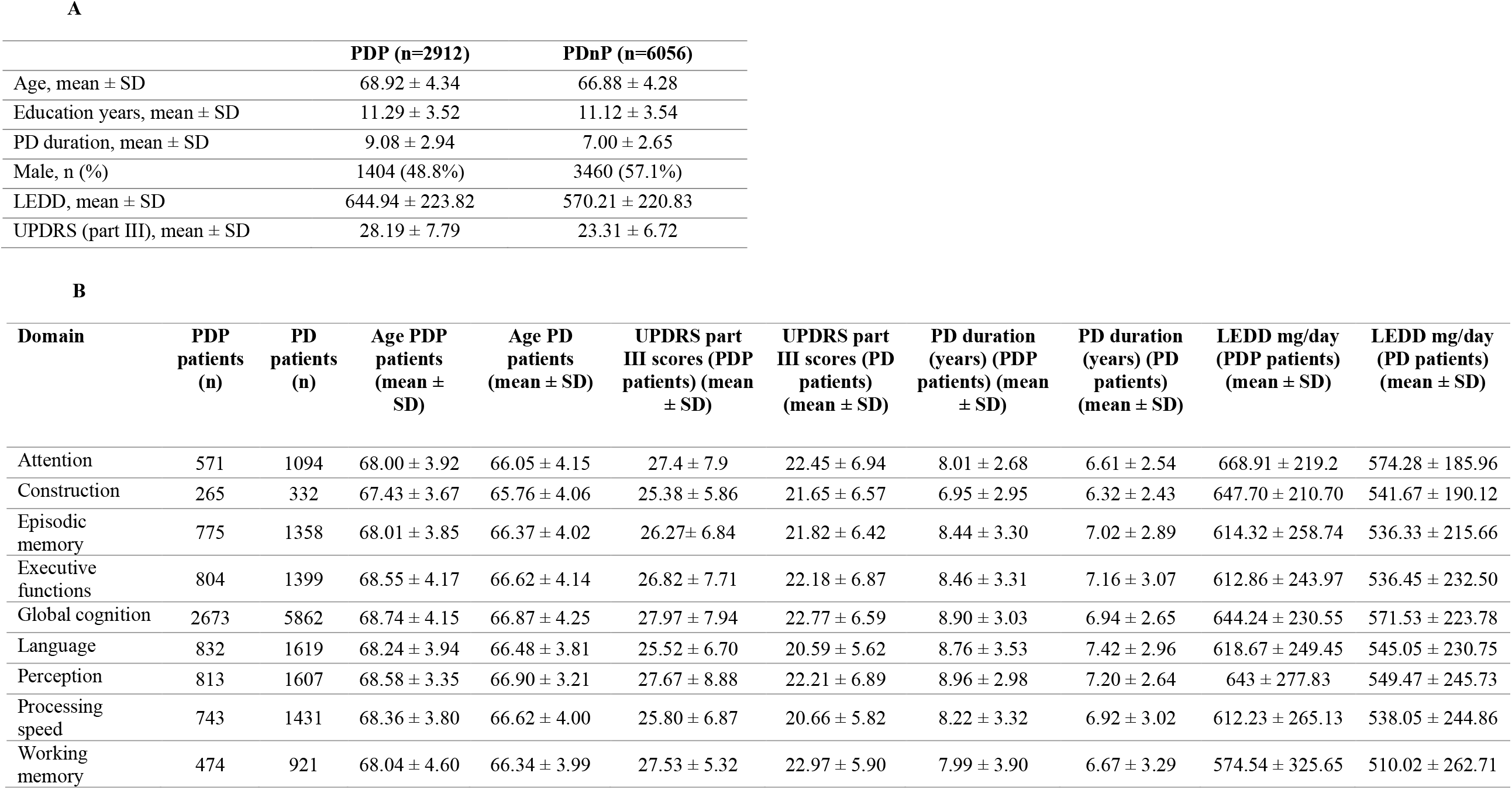
Sample characteristics (A), and sample characteristics per cognitive domain analysis (B). Motor scores are measured using the MDS-UPDRS part III scale (and previous versions), PD medications are reported in levodopa equivalent daily dose (LEDD) mg per day and PD duration is reported in mean years.

**Figure 1.**
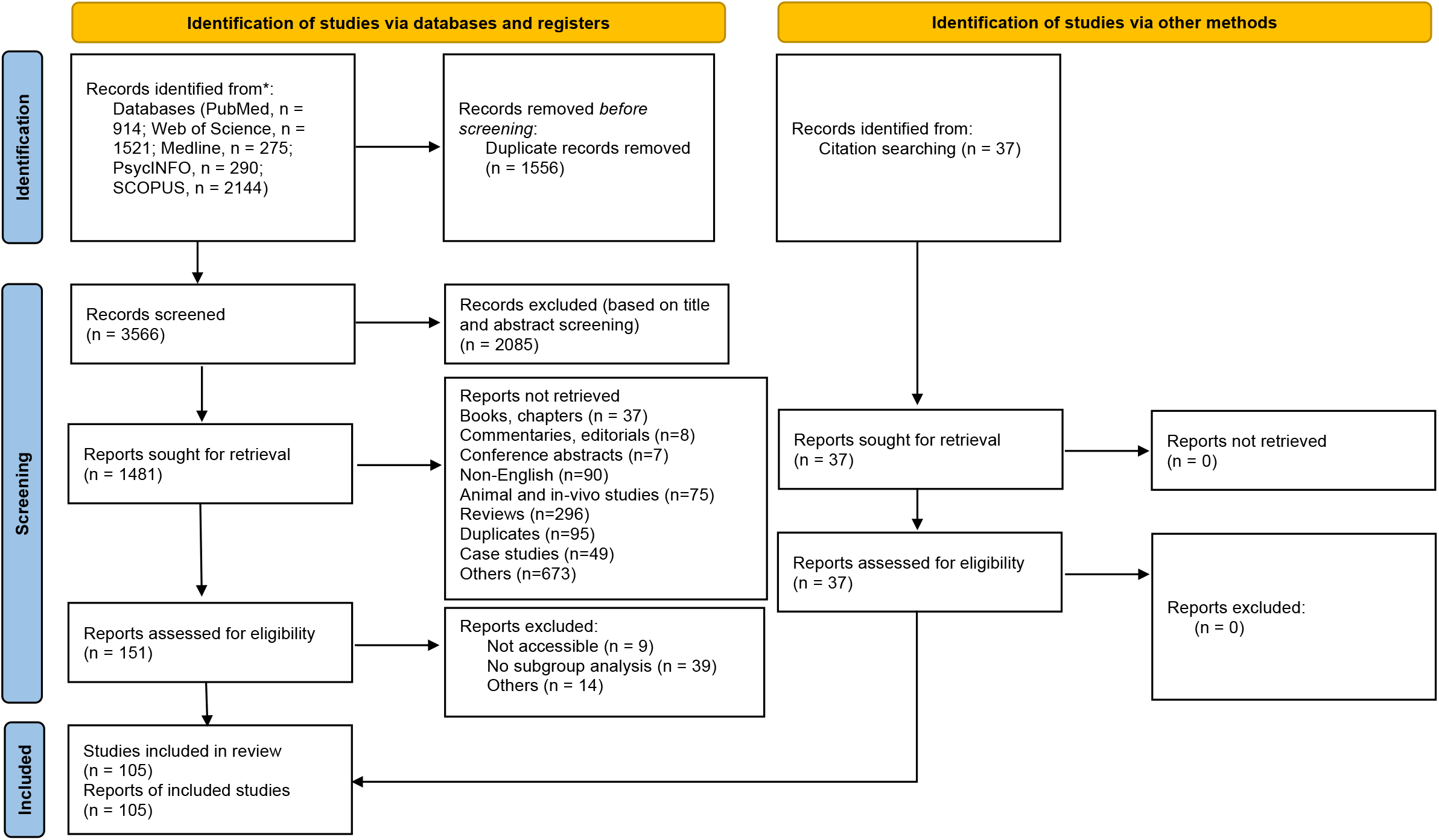
PRISMA flowchart reporting study selection procedure.

### Pooled effect-size estimates of task performance in PDP versus PDnP across different cognitive domains: Bayesian meta-analysis

Figure 2A reports the forest plot with posterior probability draws for each major cognitive domain. A negative effect-size estimate indicates that PDP patients performed worse while a positive estimate indicates that they performed better compared to PDnP patients in that specific cognitive domain. All the effect-size estimates across all nine major domains indicate that task performance across all these cognitive domains tested was worse in PDP compared to PDnP patients, with the largest effect sizes observed in construction, processing speed, episodic memory, global cognition and executive functions Overall results for domains and sub-domains are reported in Table 2. Presence of both between- and within-study heterogeneity were detected across all analyses, both in the domains and sub-domains related analyses. Publication bias was also present in the majority of the analyses. See Supplementary Material 4 for funnel plots and posterior probability forest plots for each domain and sub-domain (eFigures 2-43).

**Figure 2.**
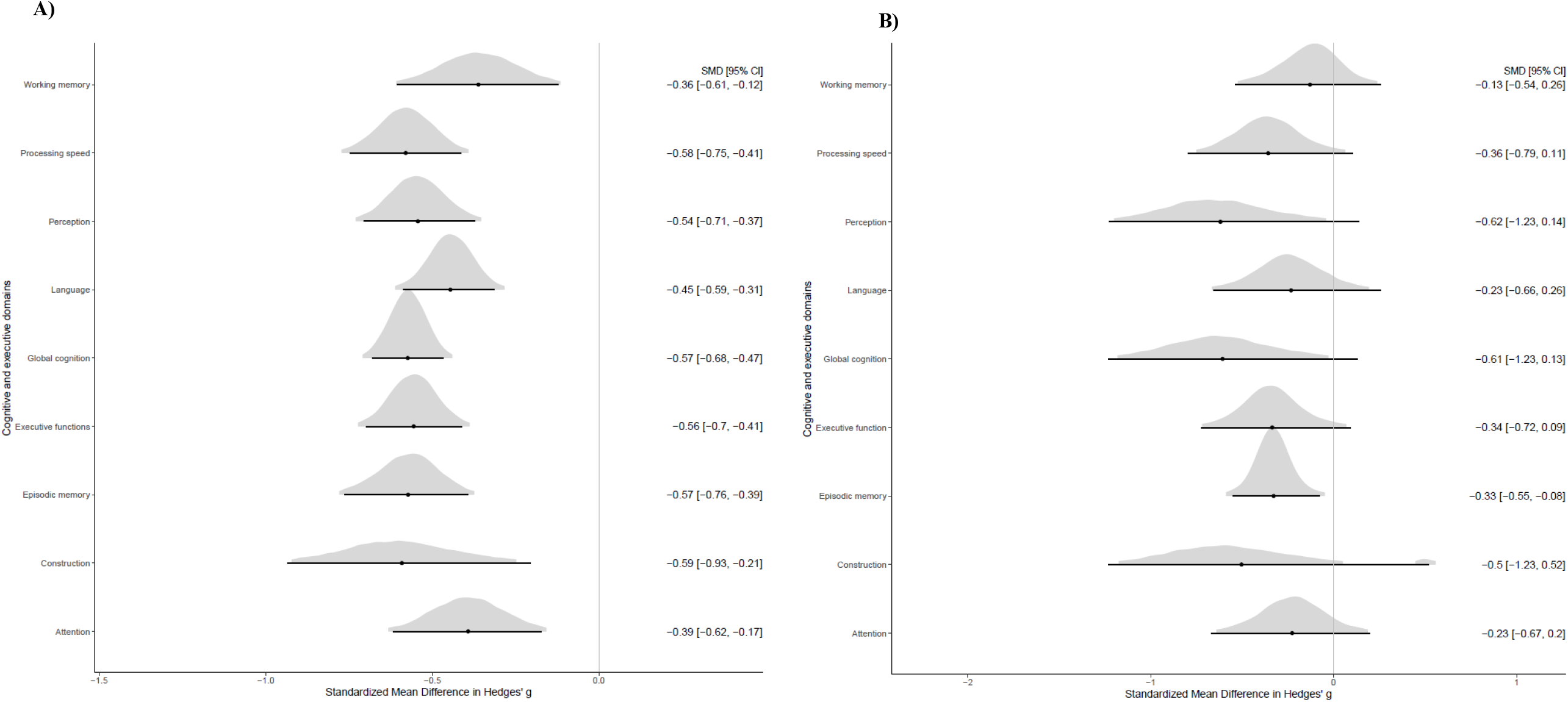
A) Forest plot showing the probability distribution for standard mean difference (SMD, followed by 95% credible intervals) for each domain (k=105 studies; PDP, n=2912, PDnP, n=6056). B) Forest plot showing the probability distribution for standard mean difference (SMD, followed by 95% credible intervals) for each domain across the same sample of PDP (n=124) and PDnP patients (n=139) (k=4 studies).

**Table 2.** Standard mean difference (SMD) expressed in Hedges’ g, with respective 95% credible intervals (95% CI) and cumulative probability, are reported here for all domains. Between and within-study variabilities is also reported with associated 95% CI.

| Domain | SMD, $g$ (95% CI) [cumulative probability of SMD < -0.3] | Variability (expressed in tau, $\tau$, with 95% CI) |
| --- | --- | --- |
| **Language (k=34)** | -0.45 (-0.59 to -0.31) [98.6%] | Between-study: 0.29 (0.12 to 0.44)<br>Within-study: 0.19 (0.02 to 0.36) |
| Reading (k=7) | -0.27 (-0.71 to 0.17) | Between-study: 0.25 (0.01 to 0.72)<br>Within-study: 0.45 (0.08 to 0.90) |
| Phonemic fluency (k=17) | -0.47 (-0.71 to -0.25) | Between-study: 0.29 (0.03 to 0.58)<br>Within-study: 0.19 (0.01 to 0.46) |
| Semantic fluency (k=19) | -0.63 (-0.89 to -0.39) | Between-study: 0.50 (0.34 to 0.74)<br>Within-study: 0.05 (0 to 0.14) |
| Naming (k=10) | -0.28 (-0.57 to -0.03) | Between-study: 0.19 (0.01 to 0.53)<br>Within-study: 0.23 (0.01 to 0.55) |
| **Construction (k=11)** | -0.59 (-0.93 to -0.21) [93.7%] | Between-study: 0.28 (0.01 to 0.70)<br>Within-study: 0.58 (0.32 to 0.90) |
| Drawing (k=5) | -0.64 (-1.53 to 0.33) | Between-study: 0.51 (0.02 to 1.56)<br>Within-study: 0.97 (0.36 to 1.96) |
| Copying (k=7) | -0.74 (-1.08 to -0.41) | Between-study: 0.16 (0.01 to 0.51)<br>Within-study: 0.26 (0.01 to 0.75) |
| **Working memory (k=16)** | -0.36 (-0.61 to -0.12) [69.8%] | Between-study: 0.40 (0.21 to 0.67)<br>Within-study: 0.22 (0.08 to 0.36) |
| Manipulation (k=10) | -0.41 (-0.82 to 0.01) | Between-study: 0.59 (0.29 to 1.05)<br>Within-study: 0.12 (0 to 0.36) |
| Maintenance (k=10) | -0.32 (-0.64 to 0.00) | Between-study: 0.39 (0.13 to 0.73)<br>Within-study: 0.24 (0.02 to 0.50) |
| **Episodic memory (k=30)** | -0.57 (-0.76 to -0.39) [99.6%] | Between-study: 0.43 (0.21 to 0.70)<br>Within-study: 0.29 (0.17 to 0.42) |
| Encoding (k=12) | -0.78 (-1.21 to -0.37) | Between-study: 0.56 (0.06 to 1.08)<br>Within-study: 0.25 (0.01 to 0.74) |
| Non-verbal retrieval (k=12) | -0.61 (-0.88 to -0.34) | Between-study: 0.15 (0.01 to 0.44)<br>Within-study: 0.46 (0.25 to 0.73) |
| Verbal retrieval (k=26) | -0.52 (-0.78 to -0.29) | Between-study: 0.52 (0.22 to 0.88)<br>Within-study: 0.17 (0.01 to 0.38) |
| **Perception (k=34)** | -0.54 (-0.71 to -0.37) [99.6%] | Between-study: 0.19 (0.01 to 0.40)<br>Within-study: 0.63 (0.51 to 0.77) |
| Visual acuity (k=8) | -0.19 (-0.64 to 0.25) | Between-study: 0.35 (0.02 to 0.87)<br>Within-study: 0.31 (0.01 to 0.80) |
| Dorsal stream (k=19) | -0.44 (-0.65 to -0.22) | Between-study: 0.21 (0.02 to 0.45)<br>Within-study: 0.46 (0.29 to 0.66) |
| Ventral stream (k=8) | -1.03 (-1.60 to -0.43) | Between-study: 0.60 (0.03 to 1.40)<br>Within-study: 0.45 (0.03 to 1.09) |
| Dorsal stream/Ventral stream (k=7) | -0.59 (-1.31 to 0.17) | Between-study: 0.78 (0.05 to 1.81)<br>Within-study: 0.58 (0.05 to 1.33) |
| Low level vision apperception (k=9) | -0.87 (-1.38 to -0.37) | Between-study: 0.30 (0.01 to 0.90)<br>Within-study: 0.81 (0.48 to 1.26) |
| Imagery (k=2) | 0.35 (-0.60 to 1.32) | Between-study: 0.69 (0.03 to 2.59) |
|  |  | Within-study: 0.17 (0.01 to 0.55) |
| Visual contrast (k=4) | - 0.41 (-0.87 to 0.14) | Between-study: 0.27 (0.01 to 0.92)<br>Within-study: 0.28 (0.02 to 0.70) |
| **Executive functions** (k=33) | -0.56 (-0.70 to -0.41) [99.9%] | Between-study: 0.30 (0.12 to 0.47)<br>Within-study: 0.30 (0.19 to 0.41) |
| **Processing speed** (k=29) | -0.58 (-0.75 to -0.41) [99.9%] | Between-study: 0.37 (0.25 to 0.53)<br>Within-study: 0.8 (0.00 to 0.22) |
| **Attention** (k=24) | -0.39 (-0.62 to -0.17) [79.7%] | Between-study: 0.40 (0.11 to 0.64)<br>Within-study: 0.26 (0.02 to 0.55) |
| **Global cognition** (k=103) | -0.57 (-0.68 to -0.47) [100%] | Between-study: 0.41 (0.29 to 0.52)<br>Within-study: 0.28 (0.14 to 0.42) |

### Cumulative probability of the most likely impaired domains

To identify the most likely domains affected in PDP, we applied the empirical cumulative distribution function and set effect-size of -0.3 as threshold to the estimates reported above. Of all the domains tested, only the domains of global cognition, processing speed, executive function, episodic memory and perception were found to have over 99.5% probability of impairment equivalent to an effect size of at least -0.3 or worse (Table 2).

### Task-related performance in the same patient sample

In addition, data from 4 studies that assessed all nine domains were meta-analysed ^12, 30-32^ separately, so as to compare relative impairments across all the domains in the same group of participants. Figure 2B reports the posterior probability distribution across the nine domains for these four studies. PDP patients were more likely to show a worse performance on only episodic memory tests compared to PDnP patients (*g*=-0.33 (95% CI -0.55, -0.08; between-study variability, 0.16, 95% CI 0.01, 0.53; within-study variability 0.10, 95% CI 0.00, 0.28)). Presence of between-study heterogeneity was detected in this analysis. There were no differences between PDP and PDnP patients included in these four studies and those included in the general study pool (k=101) (see Supplementary Material 5).

### Effect of potential confounding factors: Bayesian meta-regression

Meta-regression results are shown in Table 3. In brief, the likelihood that LEDD may be associated with cognitive and executive deficits was negligible across all domains. Similarly, psychosis severity was reported in global cognition but may be less likely to be associated with task-related performance. Age was likely associated with task-related performance on all domains, except for attention. PD duration was likely to influence performance on global cognitive assessments, construction, executive functions and working memory. Severity of depression likely moderates performance in global cognition, executive functions, processing speed and language. All three moderators were inversely associated with task performance, such that, the longer the disease duration, the more severe the depression and the greater the age, the worse the performance on cognitive domains.

**Table 3.** Number of studies, regression coefficients (or β values), with respective 95% credible intervals (95% CI) are reported here for the nine major cognitive domains for depression and psychosis severity, severity of PD (measured with MDS-UPDRS part III), age, duration of PD and PD medications (expressed in LEDD).

| Cognitive domains | Depression (beta value, 95% CI) | Psychosis Severity (beta value, 95% CI) | UPDRS part III Motor symptoms (beta value, 95% CI) | Age (beta value, 95% CI) | PD duration (years) (beta value, 95% CI) | LEDD (beta value, 95% CI) |
| --- | --- | --- | --- | --- | --- | --- |
| Global cognition | -0.08 (-0.16 to -0.01), k=49 | -0.09 (-0.22 to 0.03), n=16 | -0.00 (-0.02 to 0.01), k=79 | -0.04 (-0.06 to -0.02), k=95 | -0.06 (-0.10 to -0.03), k=92 | 0.00 (0.00-0.00), k=90 |
| Episodic memory | -0.21 (-0.51 to 0.04), k=14 | NA | 0.00 (-0.04 to 0.04), k=23 | -0.04 (-0.08 to -0.01), k=30 | -0.06 (-0.13 to 0.00), k=28 | 0.00 (0.00-0.00), k=25 |
| Attention | 0.27 (0.01 to 0.55), k=14 | NA | -0.01 (-0.03 to 0.01), k=18 | 0.01 (-0.04 to 0.06), k=23 | 0.02 (-0.08 to 0.11), k=20 | 0.00 (0.00-0.00), k=22 |
| Construction | NA | NA | 0.03 (-0.07 to 0.14), k=9 | -0.08 (-0.16 to -0.02), k=11 | -0.13 (-0.27 to -0.01), k=10 | 0.00 (0.00-0.00), k=10 |
| Executive functions | -0.26 (-0.38 to -0.14), k=16 | NA | -0.01 (-0.02 to 0.02), k=24 | -0.05 (-0.08 to -0.02), k=32 | -0.06 (-0.10 to -0.01), k=28 | 0.00 (0.00-0.00), k=29 |
| Processing speed | -0.25 (-0.35 to -0.14), k=14 | NA | -0.00 (-0.03 to 0.03), k=22 | -0.06 (-0.10 to -0.02), k=28* | -0.05 (-0.11 to 0.00), k=24 | 0.00 (0.00-0.00), k=26 |
| Working memory | -0.06 (-0.23 to 0.16), k=10 | NA | -0.02 (-0.05 to 0.02), k=12 | -0.06 (-0.11 to -0.01), k=15 | -0.07 (-0.13 to -0.01), k=14 | 0.00 (0.00-0.00), k=15 |
| Language | -0.13 (-0.22 to -0.05), k=18 | NA | 0 (-0.02 to 0.03), k=24 | -0.04 (-0.07 to -0.02), k=33 | -0.02 (-0.07 to 0.02), k=29 | 0.00 (0.00-0.00), k=30 |
| Perception | -0.09 (-0.31 to 0.14), k=17 | NA | -0.01 (-0.03 to 0.02), k=26 | -0.08 (-0.12 to -0.04), k=33* | -0.01 (-0.08 to 0.06), k=30 | 0.00 (0.00-0.00), k=28 |

NA: not applicable
\*Divergence of N>100 present in the analysis.

## Discussion

Here we endeavoured to uncover what the probable estimate of cognitive performance might be in PDP patients using a Bayesian meta-analysis. Previous studies reported cognitive deficits in PDP patients on a range of cognitive or executive measures, but what the true effect is likely to be remained unanswered. We found that compared to those without psychosis, PD patients with psychosis are likely to show a worse task-related performance on all major domains, and sub-domains with the largest effect sizes on measures assessing construction, episodic memory, processing speed, global cognition, and executive functions. We further observed deficits in several specific sub-domains such as semantic and phonemic fluency, and naming, copying, learning, verbal and nonverbal retrieval, and higher order visual processing (i.e., dorsal and ventral streams, and low-level vision apperception). This is in line with previous evidence, indicating extensive cognitive impairments in PD psychosis also encompassing sub-areas of cognitive, perceptual and executive abilities ^11, 12, 33, 34^. In addition, using a threshold of impairment equivalent to an effect-size estimate of at least -0.3 or worse, we found that although the probability of impairment for any of the nine domains was higher than 69%, the domains of general cognitive abilities, executive function, processing speed, memory, and perception were the most likely to be impaired in PDP with probabilities > 99.5%. This is in line with and extends on previous findings suggesting that although overall cognitive and executive functions may be disrupted in PDP ^9^, global cognitive and executive functioning, processing speed and episodic memory (and its sub-domains) may be the key domains affected. We observed similar effect sizes to those reported in Montagnese et al. ^9^, with some discrepancy in magnitude of impairment estimated. This may be related to the fact that we applied a non-frequentist approach to this meta-analysis, which allows computation of pooled effect-size (per cognitive domain) based on probability distribution drawn from a prior (based on previous established knowledge) and the data from the 105 studies ^35, 36^. A number of cognitive sub-domains appeared impacted in PDP, showing a similar pattern of results favouring the presence of extensive impairments. Finally, when we restricted our analyses to those four studies that reported on all domains, we found that episodic memory was the only domain that was significantly impaired in PDP patients compared to PDnP patients. However, whether this indicates that episodic memory deficits underlie the emergence of psychotic symptoms in PD, whereas involvement of other domains reflect progression of PD in general, remains to be tested.

Age was the most common moderator likely associated with impaired performance, across all major domains except for attention, progressing age being associated with worse performance, in contrast to Montagnese et al.^9^, who reported an effect of age only for global cognition, executive functions and working memory. PD duration also moderated performance on global cognition, construction, executive functions and working memory, with longer duration associated with worse performance. Dopamine-replacement medications (expressed in LEDD) were unlikely to affect task-related performance in PDP patients, in accordance with previous evidence ^37, 38^. We also observed an inverse relationship between performance on global cognition, executive functions, processing speed, and language, and depression indicating that, like with PD duration and age, the greater the depression the worse the cognitive performance. This is in agreement with previous literature ^39, 40^, highlighting the effect of neuropsychiatric symptoms on cognitive functioning in PD as well as PD psychosis patients.

Our findings are to some extent in agreement with the attentional network dysfunction model of psychosis in PD ^41^, specifically in relation to processing speed and episodic memory. This model suggests that visual hallucinations in PD may be a by-product of malfunctional information processing across the ventral and dorsal attentional networks, and Default Mode Network. Consistent with this, we observed possible deficits in measurements assessing processing speed, as well as general cognitive and executive functions, which are associated with these cortical networks. However, based on the present findings, it may be challenging to propose one model to explain psychosis in PD. Rather it may be related to impairment in a range of cognitive processes and structure and/ or function of neural substrates subserving those processes. Nevertheless, these results suggest that future research should focus on episodic memory and its sub-categories (i.e., recognition, cued and free recall, false positive error in recognition tests) and its relationship with psychosis in PD. Further, future studies may employ neuroimaging approaches in conjunction with memory tasks or memory-related cognitive activation paradigms to examine whether altered memory-related brain function may underlie psychosis in PD.

### Limitations and future research

Results reported here are to be considered in light of a range of limitations some of which are related to the specific analytic approach that we have used here, and others pertain to limitations of the meta-analytic approach in general. Focusing on the former category, firstly, we used a somewhat non-informative prior in our analyses. Specifically, we used a normal distribution for the population-level parameter, i.e., where we would expect a potential effect size to be, and the Half-Cauchy prior for the distribution parameter, tau (τ). Half-Cauchy distribution has been recommended in research where sample sizes are relatively small and for its properties ^22^. Although this population-level prior does not provide extensive information, it may be useful especially in initial investigations on a specific problem based on the belief that all probabilities are equal ^42^ and can set the foundation for exploratory analyses. Using a meta-analytic approach, we were not able to examine an exhaustive list of cognitive domains. Instead, we were only able to synthesize the effect-size of cognitive and executive domains that have been investigated in the included studies, applying recommended guidelines for the domain classification ^14^. Therefore, it is plausible that cognitive impairments other than those examined in these 105 studies may be impaired in people with PD psychosis. Nevertheless, it is worth noting that we also examined sub-domains ^14^ (e.g., manipulation, encoding, semantic fluency), and it may be possible that deficits in the overall domain (e.g., language) might be actually due to impairments in the respective sub-domains such as, for example, category- and letter-based fluency as well as naming abilities instead of difficulties in comprehension (i.e., reading). Understanding where the putative impairments lie may lead to better understanding of the neurocognitive profile of PD psychosis and the extent to which each domain contributes to psychotic symptoms in PD. One other limitation relates to the task performance in the same patient sample across the nine cognitive domains. We found only four studies that examined all domains and due to the very small sample (PDP, n=129 patients; PDnP, n=139 patients) it may be challenging to compare these results with those from the larger group of studies (k=101), as they may not be as representative. However, we tested for discrepancies in key characteristics, e.g., age, motor symptoms, PD medications, and both samples were comparable, thus suggesting that differences between the samples of the two sets of studies may not explain the discrepant result. Moreover, presence of between- and within-study heterogeneity and publication bias across the analyses warrant cautious interpretation of the effect-size estimates. Such variability and publication bias were also reported in Montagnese et al. ^9^. Another limitation relates to the fact that we primarily examined the effect of PD medications expressed in LEDD in the meta-regressions. LEDD is one of the most common clinical parameters to measure the amount of dopaminergic medications (i.e., dopamine agonists, levodopa, levodopa intestinal gel, etc.) ^43^. PDP patients may however be on different other medications including antipsychotics, anticholinergic drugs, as well as antidepressants ^44, 45^. We observed a lack of consistent reporting, for example studies have reported the number or percentage of patients on a given medication, e.g., apomorphine, donepezil, rivastigmine, whilst others report just the category of the medication (either in number or percentage and at times in mg per day). This limited our ability to more comprehensively account for the role of medications in cognitive impairment in PDP. In this work, we mainly focused on cross-sectional studies that examined PDP patients at one single time points. Although this is informative, PD psychosis is a risk factor for more severe PD symptoms, worsening of quality of life, increased hospitalisations as well as PD dementia ^4, 11^. Future research could attempt to examine the relationship between psychosis and impaired cognitive performance in longitudinal.

## Conclusion

This systematic review and meta-analysis examined the probable cognitive and executive domains affected in PD psychosis patients using a Bayesian approach. Our findings suggest extensive deficits across the major cognitive domains in PD psychosis compared to PD patients without psychosis, with global cognitive functions, processing speed, executive functions and memory as the most likely affected domains. Of those, episodic memory may be the most robustly impaired domain as suggested by analyses from the set of studies investigating all domains. PD medications did not affect the relationship between PDP and task-related performance; however, age, PD duration and severity of depression may also contribute to worsening of cognitive performance on the majority of these domains. These findings are to be considered preliminary and warrant replication in future studies incorporating a prospective longitudinal design to investigate whether these impairments in these cognitive domains may be causally associated with psychotic symptoms in people with PD.

## Data Availability

Data availability statement: This publication is a systematic review and meta-analysis, and as such it includes data that have already been published. The data that support the findings of this study are available from the corresponding author upon request.

## Acknowledgements

None

## References

1. Ffytche DH, Creese B, Politis M, et al. The psychosis spectrum in Parkinson disease. Nat Rev Neurol 2017;13(2):81–95.

2. Goetz CG, Stebbins GT. Risk factors for nursing home placement in advanced Parkinson’s disease. Neurology 1993;43(11):2227–2229.

3. McKinlay A, Grace RC, Dalrymple-Alford JC, Anderson T, Fink J, Roger D. A profile of neuropsychiatric problems and their relationship to quality of life for Parkinson’s disease patients without dementia. Parkinsonism & Related Disorders 2008;14(1):37–42.

4. Martinez-Martin P, Rodriguez-Blazquez C, Forjaz MJ, et al. Neuropsychiatric symptoms and caregiver’s burden in Parkinson’s disease. Parkinsonism Relat Disord 2015;21(6):629–634.

5. Moustafa AA, Krishna R, Frank MJ, Eissa AM, Hewedi DH. Cognitive correlates of psychosis in patients with Parkinson’s disease. Cognitive Neuropsychiatry 2014;19(5):381–398.

6. Factor SA, Scullin MK, Sollinger AB, et al. Cognitive correlates of hallucinations and delusions in Parkinson’s disease. J Neurol Sci 2014;347(1-2):316–321.

7. Barnes J, Boubert L. Executive functions are impaired in patients with Parkinson’s disease with visual hallucinations. Journal of Neurology Neurosurgery and Psychiatry 2008;79(2):190–192.

8. Morgante L, Colosimo C, Antonini A, et al. Psychosis associated to Parkinson’s disease in the early stages: relevance of cognitive decline and depression. Journal of Neurology, Neurosurgery & Psychiatry 2012;83(1):76–82.

9. Montagnese M, Vignando M, Ffytche D, Mehta MA. Cognitive and visual processing performance in Parkinson’s disease patients with vs without visual hallucinations: A meta-analysis. Cortex 2022;146:161–172.

10. Folstein MF, Folstein SE, McHugh PR. “Mini-mental state”: a practical method for grading the cognitive state of patients for the clinician. Journal of psychiatric research 1975;12(3):189–198.

11. Santangelo G, Trojano L, Vitale C, et al. A neuropsychological longitudinal study in Parkinson’s patients with and without hallucinations. Movement Disorders 2007;22(16):2418–2425.

12. Shin S, Lee JE, Hong JY, Sunwoo MK, Sohn YH, Lee PH. Neuroanatomical substrates of visual hallucinations in patients with non-demented Parkinson’s disease. J Neurol Neurosurg Psychiatry 2012;83(12):1155–1161.

13. Williams-Gray CH, Foltynie T, Lewis SJ, Barker RA. Cognitive deficits and psychosis in Parkinson’s disease. CNS drugs 2006;20(6):477–505.

14. Harvey PD. Domains of cognition and their assessment Dialogues Clin Neurosci 2019;21(3):227–237.

15. Page MJ, McKenzie JE, Bossuyt PM, et al. The PRISMA 2020 statement: an updated guideline for reporting systematic reviews. BMJ 2021;372:n71.

16. Team RC. R: A language and environment for statistical computing. 2013.

17. Viechtbauer W. Conducting meta-analyses in R with the metafor package. Journal of statistical software 2010;36(3):1–48.

18. Bürkner P-C. brms: An R Package for Bayesian Multilevel Models Using Stan. Journal of Statistical Software 2017;80(1):1 –28.

19. Team SD. Stan: A C++ library for probability and sampling. Online: http://mc-stanorg 2014.

20. Van den Noortgate W, López-López JA, Marín-Martínez F, Sánchez-Meca J. Three-level meta-analysis of dependent effect sizes. Behavior Research Methods 2013;45(2):576–594.

21. Gelman A. Prior distributions for variance parameters in hierarchical models (comment on article by Browne and Draper). Bayesian Analysis 2006;1(3):515-534, 520.

22. Williams D, Rast P, Bürkner P-C. Bayesian Meta-Analysis with Weakly Informative Prior Distributions 2018.

23. Hoffman MD, Gelman A. The No-U-Turn sampler: adaptively setting path lengths in Hamiltonian Monte Carlo. J Mach Learn Res 2014;15(1):1593–1623.

24. Betancourt M, Girolami M. Hamiltonian Monte Carlo for hierarchical models. Current trends in Bayesian methodology with applications 2015;79(30):2–4.

25. Stroop JR. Studies of interference in serial verbal reactions. Journal of experimental psychology 1935;18(6):643.

26. Ivnik RJ, Malec JF, Smith GE, Tangalos EG, Petersen RC. Neuropsychological tests’ norms above age 55: COWAT, BNT, MAE Token, WRAT-R Reading, AMNART, STROOP, TMT, and JLO. Clinical Neuropsychologist 1996;10(3):262–278.

27. Lin L, Chu H. Quantifying publication bias in meta-analysis. Biometrics 2018;74(3):785–794.

28. Konstantopoulos S. Fixed effects and variance components estimation in three-level meta-analysis. Res Synth Methods 2011;2(1):61–76.

29. Ephpp P. Quality assessment tool for quantitative studies. Hamilton: The Effective Public Health Practice Project URL: http://www.ephppca/PDF/Quality%20Assessment2010;20.

30. Llebaria G, Pagonabarraga J, Martínez-Corral M, et al. Neuropsychological correlates of mild to severe hallucinations in Parkinson’s disease. Mov Disord 2010;25(16):2785–2791.

31. Hepp DH, da Hora CC, Koene T, et al. Cognitive correlates of visual hallucinations in non-demented Parkinson’s disease patients. Parkinsonism & Related Disorders 2013;19(9):795–799.

32. Lefebvre S, Baille G, Jardri R, et al. Hallucinations and conscious access to visual inputs in Parkinson’s disease. Scientific Reports 2016;6(1):36284.

33. Grossi D, Trojano L, Pellecchia MT, Amboni M, Fragassi NA, Barone P. Frontal dysfunction contributes to the genesis of hallucinations in non-demented Parkinsonian patients. Int J Geriatr Psychiatry 2005;20(7):668–673.

34. Lenka A, Hegde S, Arumugham S, Yadav R, Pal P. Cognitive dysfunction in patients with Parkinson’s disease and psychosis. Movement Disorders 2019;v34.

35. McNeish D. On Using Bayesian Methods to Address Small Sample Problems. Structural Equation Modeling: A Multidisciplinary Journal 2016;23(5):750–773.

36. Chung Y, Rabe-Hesketh S, Dorie V, Gelman A, Liu J. A nondegenerate penalized likelihood estimator for variance parameters in multilevel models. Psychometrika 2013;78(4):685–709.

37. Growdon J, Kieburtz K, McDermott M, Panisset M, Friedman J. Levodopa improves motor function without impairing cognition in mild non-demented Parkinson’s disease patients. Neurology 1998;50(5):1327–1331.

38. Morrison C, Borod J, Brin M, Hälbig T, Olanow C. Effects of levodopa on cognitive functioning in moderate-to-severe Parkinson’s disease (MSPD). Journal of Neural Transmission 2004;111(10):1333–1341.

39. Weintraub D, Morales KH, Duda JE, Moberg PJ, Stern MB. Frequency and correlates of co-morbid psychosis and depression in Parkinson’s disease. Parkinsonism & related disorders 2006;12(7):427–431.

40. Starkstein SE, Bolduc PL, Mayberg HS, Preziosi TJ, Robinson RG. Cognitive impairments and depression in Parkinson’s disease: a follow up study. Journal of Neurology, Neurosurgery & Psychiatry 1990;53(7):597–602.

41. Shine JM, Halliday GM, Naismith SL, Lewis SJG. Visual Misperceptions and Hallucinations in Parkinson’s Disease: Dysfunction of Attentional Control Networks? Movement Disorders 2011;26(12):2154–2159.

42. Kamary K, Robert CP. Reflecting about selecting noninformative priors. arXiv preprint arXiv:14026257 2014.

43. Tomlinson CL, Stowe R, Patel S, Rick C, Gray R, Clarke CE. Systematic review of levodopa dose equivalency reporting in Parkinson’s disease. Mov Disord 2010;25(15):2649–2653.

44. Rezak M. Current Pharmacotherapeutic Treatment Options in Parkinson’s Disease. Disease-a-Month 2007;53(4):214–222.

45. Mills KA, Greene MC, Dezube R, Goodson C, Karmarkar T, Pontone GM. Efficacy and tolerability of antidepressants in Parkinson’s disease: a systematic review and network meta-analysis. International journal of geriatric psychiatry 2018;33(4):642–651.

