## Supplementary Material for "Cognitive and executive impairments in Parkinson’s Disease psychosis: a Bayesian meta-analysis"

### Supplementary Material 1

Search conducted on Web of Science on the 14th Sept 2021

7
**#6 AND #1**
Edit
Add to Search
1,521

6
**#5 OR #4**
Edit
Add to Search
6,156

5
**#2 AND #3**
Edit
Add to Search
6,156

4
**((TS=(Parkinson's disease psychosis)) OR TS=(Parkinson's psychosis)) OR TS=(Parkinson* psychosis)**
Edit
Add to Search
3,335

3
**((((((TS=(psychosis)) OR TS=(Psychosis spectrum disorder*)) OR TS=(psychosis symptom*)) OR TS=(halluci*)) OR TS=(delusion*)) OR TS=(paranoi*)) OR TS=(multimodal halluci*)**
Edit
Add to Search
161,174

2
**((TS=(Parkinson* Disease)) OR TS=(Parkinson's disease)) OR TS=(Parkinson*)**
Edit
Add to Search
238,375

1
**((((((((((((((((((((((TS=(Cognitive impairment*)) OR TS=(cognitive deficit*)) OR TS=(cognitive dysfunction*)) OR TS=(executive impairment*)) OR TS=(executive deficit*)) OR TS=(executive dysfunction*)) OR TS=(memory loss)) OR TS=(memory impair*)) OR TS=(memory deficit*)) OR TS=(attention problem*)) OR TS=(attentional deficit*)) OR TS=(language problem*)) OR TS=(language impair*)) OR TS=(visuospatial dysfunction*)) OR TS=(visuospatial impair*)) OR TS=(visuoperceptive impair*)) OR TS=(decision making)) OR TS=(orientation impair*)) OR TS=(orientation difficult*)) OR TS=(planning impair*)) OR TS=(planning deficit*)) OR TS=(working memory)) OR TS=(task-switching impair*)) OR TS=(task-shifting impair*)) OR TS=(fluency impair*)) OR TS=(fluency dysfunction*)**
Edit
Add to Search
1,686,485

**Search conducted on Medline and PsycINFO (Ovid) n 14th September 2021**

1. exp Cognitive Impairment/

2. cognitive deficit*.mp. [mp=title, abstract, heading word, table of contents, key concepts, original title, tests & measures, mesh]

3. cognitive dysfunction*.mp. [mp=title, abstract, heading word, table of contents, key concepts, original title, tests & measures, mesh]

4. executive impairment*.mp. [mp=title, abstract, heading word, table of contents, key concepts, original title, tests & measures, mesh]

5. executive deficit*.mp. [mp=title, abstract, heading word, table of contents, key concepts, original title, tests & measures, mesh]

6. executive dysfunction*.mp. [mp=title, abstract, heading word, table of contents, key concepts, original title, tests & measures, mesh]

7. memory loss.mp. [mp=title, abstract, heading word, table of contents, key concepts, original title, tests & measures, mesh]

8. memory impair*.mp. [mp=title, abstract, heading word, table of contents, key concepts, original title, tests & measures, mesh]

9. memory deficit*.mp. [mp=title, abstract, heading word, table of contents, key concepts, original title, tests & measures, mesh]

10. attention problem*.mp. [mp=title, abstract, heading word, table of contents, key concepts, original title, tests & measures, mesh]

11. attentional deficit*.mp. [mp=title, abstract, heading word, table of contents, key concepts, original title, tests & measures, mesh]

12. language problem*.mp. [mp=title, abstract, heading word, table of contents, key concepts, original title, tests & measures, mesh]

13. language impair*.mp. [mp=title, abstract, heading word, table of contents, key concepts, original title, tests & measures, mesh]

14. visuospatial dysfunction*.mp. [mp=title, abstract, heading word, table of contents, key concepts, original title, tests & measures, mesh]

15. visuospatial impair*.mp. [mp=title, abstract, heading word, table of contents, key concepts, original title, tests & measures, mesh]

16. visuoperceptive impair*.mp. [mp=title, abstract, heading word, table of contents, key concepts, original title, tests & measures, mesh]

17. decision making.mp. [mp=title, abstract, heading word, table of contents, key concepts, original title, tests & measures, mesh]

18. orientation impair*.mp. [mp=title, abstract, heading word, table of contents, key concepts, original title, tests & measures, mesh]

19. orientation difficult*.mp. [mp=title, abstract, heading word, table of contents, key concepts, original title, tests & measures, mesh]

20. planning impair*.mp. [mp=title, abstract, heading word, table of contents, key concepts, original title, tests & measures, mesh]

21. planning deficit*.mp. [mp=title, abstract, heading word, table of contents, key concepts, original title, tests & measures, mesh]

22. working memory.mp. [mp=title, abstract, heading word, table of contents, key concepts, original title, tests & measures, mesh]

23. task-switching impair*.mp. [mp=title, abstract, heading word, table of contents, key concepts, original title, tests & measures, mesh]

24. fluency impair*.mp. [mp=title, abstract, heading word, table of contents, key concepts, original title, tests & measures, mesh]

25. fluency dysfunction*.mp. [mp=title, abstract, heading word, table of contents, key concepts, original title, tests & measures, mesh]

26. 1 or 2 or 3 or 4 or 5 or 6 or 7 or 8 or 9 or 10 or 11 or 12 or 13 or 14 or 15 or 16 or 17 or 18 or 19 or 20 or 21 or 22 or 23 or 24 or 25

27. Parkinson* disease.mp. [mp=title, abstract, heading word, table of contents, key concepts, original title, tests & measures, mesh]

28. exp Parkinson's Disease/

29. Parkinson*.mp. [mp=title, abstract, heading word, table of contents, key concepts, original title, tests & measures, mesh]

30. psychosis.mp. [mp=title, abstract, heading word, table of contents, key concepts, original title, tests & measures, mesh]

31. psychosis symptom*.mp. [mp=title, abstract, heading word, table of contents, key concepts, original title, tests & measures, mesh]

32. psychosis spectrum disorder*.mp. [mp=title, abstract, heading word, table of contents, key concepts, original title, tests & measures, mesh]

33. halluci*.mp. [mp=title, abstract, heading word, table of contents, key concepts, original title, tests & measures, mesh]

34. delusion*.mp. [mp=title, abstract, heading word, table of contents, key concepts, original title, tests & measures, mesh]

35. paranoi*.mp. [mp=title, abstract, heading word, table of contents, key concepts, original title, tests & measures, mesh]

36. multimodal halluci*.mp. [mp=title, abstract, heading word, table of contents, key concepts, original title, tests & measures, mesh]

37. 30 or 31 or 32 or 33 or 34 or 35 or 36

38. 27 or 28 or 29

39. 37 and 38

40. Parkinson's disease psychosis.mp. [mp=title, abstract, heading word, table of contents, key concepts, original title, tests & measures, mesh]

41. Parkinson's psychosis.mp. [mp=title, abstract, heading word, table of contents, key concepts, original title, tests & measures, mesh]

42. Parkinson* psychosis.mp. [mp=title, abstract, heading word, table of contents, key concepts, original title, tests & measures, mesh]

43. 40 or 41 or 42

44. 39 or 43

45. 26 and 44

Medline results = 275

PsycINFO results = 290

Search conducted on SCOPUS, 14th September 2021

( ( ( ( TITLE-ABS-KEY ( parkinson* ) OR TITLE-ABS-KEY ( parkinson's AND disease ) ) ) AND ( ( TITLE-ABS-KEY ( psychosis ) OR TITLE-ABS-KEY ( halluci* ) OR TITLE-ABS-KEY ( delusion* ) OR TITLE-ABS-KEY ( paranoi* ) OR TITLE-ABS-KEY ( psychosis AND spectrum AND disorder ) OR TITLE-ABS-KEY ( multimodal AND halluci* ) OR TITLE-ABS-KEY ( psychosis AND symptom* ) ) ) ) OR ( ( ( TITLE-ABS-KEY ( parkinson* AND psychosis ) OR TITLE-ABS-KEY ( parkinson's AND disease AND psychosis ) OR TITLE-ABS-KEY ( parkinson's AND psychosis ) ) ) ) ) AND ( ( ( TITLE-ABS-KEY ( cognitive AND impairment* ) OR TITLE-ABS-KEY ( cognitive AND deficit* ) OR TITLE-ABS-KEY ( cognitive AND dysfunction* ) OR TITLE-ABS-KEY ( executive AND impairment* ) OR TITLE-ABS-KEY ( executive AND deficit* ) OR TITLE-ABS-KEY ( executive AND dysfunction* ) OR TITLE-ABS-KEY ( memory AND loss ) OR TITLE-ABS-KEY ( memory AND impair* ) OR TITLE-ABS-KEY ( memory AND deficit* ) OR TITLE-ABS-KEY ( attention AND problem* ) OR TITLE-ABS-KEY ( attentional AND deficit* ) OR TITLE-ABS-KEY ( language AND problem* ) OR TITLE-ABS-KEY ( language AND impair* ) OR TITLE-ABS-KEY ( visuospatial AND dysfunction* ) OR TITLE-ABS-KEY ( visuospatial AND impair* ) OR TITLE-ABS-KEY ( visuoperceptive AND impair* ) OR TITLE-ABS-KEY ( decision AND making ) OR TITLE-ABS-KEY ( orientation AND impair* ) OR TITLE-ABS-KEY ( orientation AND difficult* ) OR TITLE-ABS-KEY ( planning AND impair* ) OR TITLE-ABS-KEY ( planning AND deficit* ) OR TITLE-ABS-KEY ( working AND memory ) OR TITLE-ABS-KEY ( task-switching AND impair* ) OR TITLE-ABS-KEY ( task-shifting AND impair* ) OR TITLE-ABS-KEY ( fluency AND impair* ) OR TITLE-ABS-KEY ( fluency AND dysfunction* ) ) ) )

Scopus results = 2144

**Search conducted on PubMed on 14th September 2021**

| Search | Actions | Query | Results | Time |
|---|---|---|---|---|
| #7 | | Search: **(#6) AND (#1)** Filters: **English** | 914 | 05:01:48 |

| Search | Actions | Query | Results | Time |
|---|---|---|---|---|
| #6 | | Search: **(#5) OR (#4)** Filters: **English** | 3,875 | 05:01:21 |
| #5 | | Search: **(#2) AND (#3)** Filters: **English** | 3,819 | 05:01:05 |
| #4 | | Search: **((Parkinson's disease psychosis) OR (Parkinson* psychosis)) OR (Parkinson's psychosis)** Filters: **English** | 2,175 | 05:00:53 |
| #3 | | Search: **((((((Psychosis) OR (Psychosis symptom*)) OR (psychosis spectrum disorder*)) OR (halluci*)) OR (delusion*)) OR (multimodal halluci*)) OR (paranoi*)** Filters: **English** | 122,197 | 05:00:27 |
| #2 | | Search: **((Parkinson's disease) OR (Parkinson* disease)) OR (Parkinson*)** Filters: **English** | 135,647 | 04:59:57 |
| #1 | | Search: **(((((((((((((((((((((((((Cognitive impairment*) OR (Cognitive deficit*)) OR (cognitive dysfunction*)) OR (executive impairment*)) OR (executive deficit*)) OR (executive dysfunction*)) OR (memory loss)) OR (memory impair*)) OR (memory deficit*)) OR (attention problem*)) OR (attentional deficit*)) OR (language problem*)) OR (language impair*)) OR (visuospatial dysfunction*)) OR (visuospatial impair*)) OR (visuoperceptive impair*)) OR (decision making)) OR (orientation impair*)) OR (orientation difficult*)) OR (planning impair*)) OR (planning deficit*)) OR (working memory)) OR (task-switching impair*)) OR (task-shifting impair*)) OR (fluency impair*)) OR (fluency dysfunction*)** Filters: **English** | 753,178 | 04:59:38 |

#### *Data extraction*

Means and standard deviations were extracted from neuropsychological assessments for PD psychosis (PDP) and PDnP patients. Data on cognitive assessments conducted on healthy controls were not included in the analysis. When median and range, or median and interquartile range were provided, these were converted into means and standard deviations [1 2]. In addition, data on socio-demographics (i.e., age, gender, years of educations), on PD (i.e., disease duration, motor symptoms, e.g., assessed with the Movement Disorder Society Unified Parkinson's Disease Rating Scale, MDS-UPDRS [3]), medications expressed in levodopa equivalent daily dose (LEDD), and other clinical symptoms (i.e., depression, sleep disorders, etc.) were extracted. Data extraction was conducted independently by three researchers (SP, LG, RW). Discrepancies were addressed through discussion and consensus with senior researchers. Studies that conducted sub-group analysis based on different psychosis symptoms were considered as independent studies.

#### *Assessment of study quality*

We assessed the quality of included studies using the Effective Public Health Practice Project (EPHPP) scale, a 6-item quality rating scale which assesses: selection bias, study design, confounders, blinding, validity and reliability of data collection, withdrawals and dropout. Each domain is assigned a rating of "Strong", "Moderate" or "Weak". Given the nature of the studies included in the analysis, the EPHPP was adapted to match the case-control study design, therefore the "blinding" component was removed from the quality rating. Quality ratings were conducted by two authors, and discrepancies were resolved through discussion and consensus with senior researchers.

### Supplementary Material 2

The table below reports study characteristics for the 105 included studies: sample size for each group (with number of males, M) and years of education (mean unless otherwise specified), mean age, PD onset age (mean), clinical variables about PD (i.e., duration, severity and medications), diagnostic tool for PDP and depression severity.

| Study | Sample (n PDP, (male); n PDnP, (male); years of education (mean) | Age (Mean, PDP; PDnP) | PD onset (PDP; PDnP) | PD duration in years (PDP; PDnP) | UPDRS part III (mean ± SD, PDP; PDnP) | LEDD in mg/day (mean), PDP; PDnP) | Depression (mean ± SD, PDP; PDnP) |
|---|---|---|---|---|---|---|---|
| *Aarsland et al[4], Norway Group: PD patients with psychosis | 14 (6M), 138 (7M); NR | 77.1; 72.5 | 65; 63.7 | 12; 8.8 | 46.1 ± 18.9; 24 ± 13.4 | 507; 463 | MADRS 16 ± 9.1; 6.6 ± 5.8 |
| *Aarsland et al.[4] , Norway Group: PD patients with hallucinations and insights | 23 (12M), 138 (7M); NR | 78.8; 72.5 | 68.5; 63.7 | 10.3; 8.8 | 44.7 ± 15.4; 24 ± 13.4 | 509; 463 | MADRS 12.0 ± 4.9; 6.6 ± 5.8 |
| Ballanger et al.[5], Canada | 7 (4M), 7 (6M); NR | 69.2; 66.7 | NR | 4.1; 10.3 | 15.963 ± 2.198; 24.241 ± 10.627 | 464.3; 778.6 | BDI 8.93 ±6.60; 12.20 ± 7.70 |
| Barnes & Boubert[6], UK | 17 (NR), 20 (NR); NR | 67.52; 63.73 | NR | 11.93; 9.72 | NR | 510; 465 | BDI 19.12 ± 4.52; 16.48 ± 4.43 |
| Barnes & Boubert [7], UK | 19 (NR), 20 (NR); NR | 68.45; 67.43 | NR | 10; 9.53 | NR | 567; 545 | BDI 19.65 ± 5.33; 16.93 ± 5.03 |
| Barnes & David [8], UK | 21 (7M), 23 (9M); NR | 67.62; 63.23 | NR | 11.76; 8.3 | NR | 578; 670 | BDI 18.24 ± 4.5; 16.6 ± 5.4 |
| Barnes et al.[9], UK | 17 (6M), 20 (8M); NR | 67.88; 62.75 | NR | 11.88; 8.75 | NR | 498; 457 | BDI 18.24 ± 4.54; 16.6 ± 5.4 |
| Barrett et al.[10], USA | 33 (22M), 68 (38M); 16, 16 | 67.4; 67.3 | 60.8;61.5 | 6.7; 5.8 | 28.865 ± 13.949; 24.588 ± 14.769 | 635.764; 527.347 | BDI -II 11.49 ± 8.53; 7.65 ± 5.30 |
| Bejr-kasem et al.[11], Spain | 18 (10M), 14 (10M); 12.5, 11.6 | 70.4; 65.8 | NR | 5.2; 4 | 21.9 ± 8.6; 25.8 ± 9.2 | 697.2; 601.1 | HADS-D 2.2 ± 1.8; 3.3 ± 3.1 |
| Boecker et al. [12], Germany | 8 (5M), 11 (8M); NR | 72.88; 70.56 | NR | 11; 8.05 | 46.25 ± 15.98; 32.73 ± 9.00 | 667; 617 | NR |
| *Boubert & Barnes[13], UK Group: Internally driven hallucinations | 17 (NR), 20 (NR); NR | 76.42; 72.18 | NR | 12.93; 10.72 | NR | 608; 581 | BDI 17.88 ± 4.17; 16.11 ± 4.91 |
| *Boubert & Barnes [13], UK Group: Externally driven hallucinations | 18 (NR), 20 (NR); NR | 73.45; 72.18 | NR | 12.02; 10.72 | NR | 650; 581 | BDI 18.92 ± 3.23; 16.11 ± 4.91 |
| Chang et al.[14], Taiwan | 12 (7M), 23 (16M); 9.27, 10.88 | 67.79; 66.36 | NR | 11.73; 6.2 | 27.92 ± 13; 14.2 ± 8.42 | 863.8; 311.2 | HAM-D 5 ± 4.67; 4.125 ± 4.11 |
| Cho et al. [15], Canada | 11 (6M), 8 (8M); 16.2, 15.9 | 64.1; 63 | NR | 8.7; 6.7 | 24.5 ± 15.1; 14.3 ± 7 | 965.2; 687.1 | BDI 13.1 ± 6.8; 7.9 ± 6.5 |

| | | | | | | | |
|---|---|---|---|---|---|---|---|
| Chung et al.[16], Korea | 26 (NR), 32 (NR); 6.7, 7.3 | 68.4; 64.8 | 62; 58.5 | 6.458; 6.42 | NR | 710.6; 560.4 | BDI 28.2 ± 12.6; 21.2 ± 11.1 |
| Clegg et al. [17], UK | 34 (27M), 120 (73M); 12.2, 12.9 | 65.7; 66.6 | NR | 0.52; 0.5 | 28.3 ± 12; 26.5 ± 12.1 | 186.9; 175.5 | GDS-15 3.9 ± 3.1; 2.5 ± 2.3 |
| Creese et al.[18], UK | 24 (13M), 45 (27M); Education level (median) 2, 2 | 67.84; 65.68 | NR | 5.38; 3.45 | NR | 522.5; 385 | NR |
| Dauwan et al.[19], The Netherlands | 20 (13M), 20 (14M); NR | 72.15; 70.5 | NR | 8.31; 5.61 | NR | 900.98; 822.13 | BDI 14.91 ± 7.78; 9.91 ± 7.78 |
| Dave et al.[20], PPMI study (International) | 30 (21M), 355 (234M); NR | 64.07; 61.35 | NR | 8.31; 5.62 | 22.83 ± 9.6; 20.05 ± 8.9 | 526.92; 631.34 | NR |
| Debs et al.[21], France | 105 (56M), 314 (182M); NR | 70; 68 | 63; 64 | 7; 5 | 21 ± 11; 18 ± 11 | 1128; 890 | NR |
| Diederich et al.[22], USA | 14 (4M), 21 (12M); NR | 67.27; 65.31 | NR | 11.79; 10.14 | 25.71 ± 10; 21.33 ± 12.6 | 303.57; 288.09 | NR |
| Doe de Mainderille et al.[23], France | 19 (7M), 55 (39M); NR | 69.6; 66.5 | 61.4; 58.7 | NR | 17.6 ± 10; 12.4 ± 8.5 | 524; 552.3 | CES-D 24.6 ± 10.4; 15.6 ± 10.2 |
| *Factor et al.[24], USA Group: Delusions with/without hallucinations | 23 (13M), 96 (63M); 15.4, 16 | 67.8; 64.5 | 60.3; 58.1 | 8.1; 7.3 | 19.4 ± 7.4; 17 ± 8.2 | NR | NR |
| *Factor et al.[24], USA Group: Hallucinations without delusions | 25 (17M), 96 (63M); 15.4, 16 | 62.8; 64.5 | 54.6; 58.1 | 9.4; 7.3 | 18.8 ± 6.7; 17 ± 8.2 | NR | NR |
| Factor et al.[25], USA | 48 (30M), 96 (63M); 15.4, 16 | 65.2; 64.5 | 57.4; 58.1 | 8.8; 7.3 | 19.1 ± 7; 17 ± 8.2 | NR | NR |
| Fénelon et al.[26], France | 48 (27M), 130 (74M); NR | 73.9; 67.5 | 61.2; 58.9 | 12.9; 8.5 | 20.8 ± 10.3; 14 ± 7.6 | 766; 711 | CES-D 21.9 ± 9.7; 17.5 ± 9.2 |
| Fernandez et al.[27], UK | 30 (NR), 20 (NR); NR | 65; 54 | NR | 12.5; 11.15 | NR | 695; 731 | NR |
| Ffytche et al.[28], UK | 115 (71M), 286 (192M); NR | 60.77; 61.97 | NR | 0.577; 0.543 | 20.08 ± 9; 21.1 ± 8.8 | NR | GDS 2.72 ± 2.5; 2.18 ± 2.4 |
| Firbank et al.[29], UK | 17 (13M), 19 (17M); 11.6, 11.1 | 75.5; 72.3 | NR | 11; 9.6 | 55.9 ± 19.3; 34.7 ± 18.8 | 717.3; 673.5 | NR |
| Forsaa et al.[30], Norway | 70 (37M), 160 (76M); 8.3, 9.5 | 75.7; 72.6 | 65.2; 64.8 | 10.5; 7.8 | 38.7 ± 18.3; 24.3 ± 12.8 | 507; 463 | NR |
| Franciotti et al.[31], Italy | 15 (NR), 15 (NR); 10, 10 | 70; 68 | NR | 11.3; 12 | 36.3 ± 8.2; 35.6 ± 7.4 | 636; 645 | GDS-15 8.8 ± 2.6; 9.3 ±2.7 |
| Gallagher et al.[32], UK | 30 (24M), 64 (41M); NR | 70.216; 66.5 | NR | 10.737; 4.353 | 38.18 ± 15.18; 25.57 ± 12.741 | 723.619; 384.378 | HDRS 5 ± 4.67; 2.35 ± 3.79 |
| Gama et al.[33] , Brazil | 11 (6M), 28 (20M); NR | 70.6; 65.7 | 63.1; 58 | 7.4; 6.5 | 22.7 ± 7.4; 15.4 ± 8.3 | 716.6; 723 | BDI 21.9 ± 10.9; 17.5 ± 13 |
| Garofalo at al.[34] , UK | 12 (6M), 17 (10M); NR | 60.83; 63.29 | NR | 16.42; 10.94 | NR | 714.03; 614.81 | BDI 12.66 ± 7.83; 8.88 ± 4.94 |
| Gobel et al.[35], Switzerland | 7 (NR), 8 (NR); 13.29, 14 | NR | NR | 13.29; 14 | NR | 978.71; 1126.86 | NR |

| | | | | | | | |
|---|---|---|---|---|---|---|---|
| Goetz at al.[36], US | 29 (NR), 60 (NR); NR | NR | NR | NR | 32.6 ± 12.3; 26.3 ± 10.1 | 524; 480 | NR |
| Goetz et al.[37], USA | 15 (NR), 19 (NR); NR | NR | NR | NR | 28 ± 10.8; 27.3 ± 11.9 | 769; 629 | NR |
| Goldman et al.[38], USA | 25 (17M), 25 (18M); 15.4, 15.7 | 74.8; 75.4 | NR | 13.1; 10.8 | 39 ± 13.8; 43.5 ± 13.2 | 808.3; 787.8 | NR |
| Gordon et al.[39] , Brazil | 16 (15M), 52 (37M); 5.2, 6.7 | 69; 61.9 | 58.6; 54.4 | 10.1; 7.1 | 85.5 ± 22.8; 62.7 ± 23.8 | 647.5; 597.9 | NR |
| *Graham et al.[40] , UK<br>Group: Early hallucinators | 13 (7M), 54 (25M); NR | NR | 62.3; 59.3 | NR | 29.8 ± 10.8; 22.4 ± 12.7 | 457; 433 | BDI 13.4 ± 7.7; 9.9 ± 7.6 |
| *Graham et al.[40] , UK<br>Group: Late hallucinators | 13 (7M), 42 (21M); NR | NR | 52; 50 | NR | 32.3 ± 14.5; 26.1 ± 12.2 | 614; 667 | BDI 8.4 ± 2.9; 13 ± 7.9 |
| Grossi et al.[41] , Italy | 14 (9M), 34 (20M); 12.71, 8.53 | 67.36; 66.85 | NR | 10.38; 6.29 | NR | 450; 437.9 | NR |
| Grossi et al.[42] , Italy | 19 (11M), 19 (NR); 8.4, NR | 68.3; NR | NR | NR | NR | NR | NR |
| Haeske-Dewick et al.[43], Germany | 16 (9M), 20 (14M); 10.5, 10 | 72.13; 67.25 | NR | 10.5; 5.5 | NR | 400; 400 | GDS 14 ± 4.5; 9.5 ± 5.5 |
| Hall et al.[44], Australia | 25 (NR), 28 (NR); NR | 69.28; 66.65 | NR | 6.74; 6.18 | 34.63 ± 15.26; 28.15 ± 11.95 | 688.5; 590.5 | BDI-II 12.64 ± 11.64; 7.3 ± 6.38 |
| Hepp at al.[45], The Netherlands | 15 (11M), 40 (21M); NR | 69; 67 | NR | 12; 11 | 37 ± 9; 30 ± 10 | 1081; 1008 | NR |
| Hepp et al.[46], Netherlands | 31 (18M), 31 (20M); Verhage education score: 5.2/4.9 | 66; 65 | NR | 7; 8 | 32 ± 15; 26 ± 11 | 508; 602 | BDI 17 ± 9; 12 ± 10 |
| Holroyd et al.[47], USA | 26 (NR), 72 (NR); NR | NR | NR | NR | 47.8 ± 21.4; 38.5 ± 16.5 | 648.1; 593.1 | GDS 7.9 ±6.5; 5 ± 4.5 |
| Ibarretxe-Bilbao et al.[48], Spain | 12 (3M), 14 (5M); 7.5, 8.3 | 73.3; 71.1 | NR | 12.1; 11.9 | 27.2 ± 12.1; 22.2 ± 13.4 | 846.4; 879.3 | HAM-D 7 ± 4.6; 3.3 ± 3.1 |
| Ibarretxe-Bilbao et al.[49], Spain | 16 (5M), 19 (8M); 7.6, 7.8 | 73.5; 72.5 | NR | 12.9; 10.9 | 29.7 ± 12.8; 24.7 ± 14.3 | NR | HAM-D 7.4 ± 4.4; 3.6 ± 2.8 |
| Imamura et al.[50], Japan | 11 (NR), 23 (NR); NR | 74.2; 69.3 | NR | 9.5 (median); 5.7 (median) | 28.9 ± 9; 24.6 ± 7.1 | 94.4; 101 | SDS 46 ± 9.9; 40.1 ± 5.9 |
| Jacobson et al. [51], US | 38 (10M), 33 (7M); NR | 80.2; 80.5 | NR | NR | NR | 665; 884 | NR |
| Janzen et al.[52], Netherlands | 13 (6M), 16 (9M); NR | 66; 64.3 | 54.5; 61.3 | 11.5; 3.1 | 29.1 ± 8.4; 23.6 ± 11.5 | 712.2; 170.2 | NR |
| *Katzen et al.[53], USA<br>Group: VH | 47 (29M), 105 (73M); 14, 14.1 | 65.6; 64.6 | 53.9; 54.8 | 11.7; 9.9 | NR | NR | BDI-II 15.8 ± 10.4; 11.5 ± 7.5 |
| *Katzen et al.[53], USA<br>Group: Only VH | 35 (22M), 105 (73M); 14.1, 14.1 | 66.3; 64.6 | 54.8; 54.8 | 11.5; 9.9 | NR | NR | BDI-II 14.9 ± 10.6; 11.5 ± 7.5 |
| *Katzen et al.[53], USA<br>Group: VH (plus another modality) | 12 (7M), 105 (73M); 14, 14.1 | 63.5; 64.6 | 51.3; 54.8 | 12.3; 9.9 | NR | NR | BDI-II 18.9 ± 9.5; 11.5 ± 7.5 |

| | | | | | | | |
|---|---|---|---|---|---|---|---|
| Kiferle et al.[54], Italy | 60 (28M), 62 (31M); NR | 62.69; 66.03 | 54.94; 58.13 | 6.77; 7.14 | 14.74 ± 10.41; 15.13 ± 10.02 | 259; 278.2 | NR |
| Kiferle et al.[55], Italy | 18 (NR), 18 (NR); NR | 75; 76.6 | 69.5; 70.8 | 5.5; 5.775 | 27.3 ± 7.6; 25.8 ± 9.7 | 547.3; 496.2 | NR |
| Koerts et al.[56], The Netherlands | 14 (9M), 14 (10M); 4.4, 4.1 | 69; 67.11 | NR | 10.7; 6 | NR | 943.6; 693.2 | NR |
| Kopal et al.[57], Czech Republic | 18 (10M), 34 (18M); NR | 71.5; 67.7 | NR | 10.17; 7.12 | 28.06 ± 9.85; 16.65 ± 6.61 | NR | NR |
| Lee et al.[58], USA | 41 (31M), 150 (106M); 16.1, 16.5 | 64; 62.8 | NR | 8.5; 5.8 | 24.7 ± 10.4; 20.8 ± 10.2 | 620.4; 480 | IDS 25.3 ± 13; 16.6 ± 11.9 |
| Lee et al.[59], Korea | 20 (9M), 25 (9M); NR | 69.46; 69.21 | 63.217; 63.341 | NR | 26.013 ± 12.311; 21.52 ± 10.82 | 721.07; 567.542 | NR |
| Lee et al.[60], Korea | 10 (7M), 14 (5M); NR | 69.2; 66.1 | NR | 7.2; 7.3 | 22.9 ± 5.1; 20.3 ± 7.8 | 981.2; 843.4 | NR |
| Lee et al.[61], Korea | 10 (7M), 21 (9M); NR | 69.4; 68.6 | 62.2; 59.3 | 7.2; 7 | 22.5 ± 5.8; 16.4 ± 5.1 | 1031.2; 805.2 | GDS 16.6 ± 6.6; 15.5 ± 7.2 |
| Lefebvre et al.[62], France | 18 (11M), 16 (12M); 12.44, 13.38 | 63.5; 62.69 | NR | 9.06; 8 | 25 ± 8.44; 21.81 ± 7.93 | 859.72; 804.25 | MADRS 3.56 ± 2.99; 3.06 ± 2.88 |
| Lenka et al.[63], India | 42 (35M), 48 (41M); 11.3, 12.1 | 58.5; 57.9 | NR | 6.5; 5.7 | 36.3 ± 8.1; 35.2 ± 8.4 | 722.8; 577.5 | NR |
| Lenka et al.[64], India | 42 (36M), 51 (43M); NR | 58.7; 57.8 | 52.1; 50.9 | 6.6; 5.8 | 12.4 ± 5.7; 12.1 ± 5.8 | 729.1; 565.4 | HAM-D 8.6 ± 5.5; 6.9 ± 4.8 |
| Leu-Semenescu et al.[65], France | 26 (18M), 74 (48M); NR | 62.5; 63.9 | 55; 57 | 7; 6.9 | 25.8 ± 12.5; 17.7 ± 10.1 | 742; 620 | BDI 12.9 ± 6.1; 10.5 ± 6.7 |
| *Llebaria et al.[66], Spain Group: Patients with VH | 11 (NR), 28 (NR); 9.7, 8.4 | 71.1; 72.7 | NR | 7.6; 7.9 | 26.1 ± 4; 26.7 ± 3 | 987; 775.9 | NR |
| *Llebaria et al.[66], Spain Group: Patient with VH with insight | 10 (NR), 28 (NR); 8.5, 8.4 | 75.8; 72.7 | NR | 8.5; 7.9 | 24.3 ± 3; 26.7 ± 3 | 854; 775.9 | NR |
| *Llebaria et al.[66] , Spain Group: Patients with VH without insight | 8 (NR), 28 (NR); 7, 8.4 | 79.2; 72.7 | NR | 10.2; 7.9 | 22.2 ± 5; 26.7 ± 3 | 815.6; 775.9 | NR |
| Mack et al.[67] , USA | 65 (40M), 40 (27M); 15.6, 16.6 | 64.2; 69.9 | 54; 63.69 | 10.2; 6 | 16.2 ± 1.3; 17.7 ± 1.7 | 606.3; 583.3 | HAM-D 7.7 ± 0.6; 3.3 ± 0.2 |
| Manganelli et al.[68], Italy | 10 (6M), 12 (7M); NR | 70.4; 65.5 | NR | 8.7; 9 | 16.1 ± 6.9; 17.6 ± 4.2 | 535.9; 697.9 | NR |
| Matsui et al.[69], Japan | 10 (5M), 9 (1M); NR | 69.3; 67.5 | NR | 14.4; 4.4 | NR | 355; 252.8 | NR |
| Matsui et al.[70], Japan | 31 (8M), 39 (14M); NR | 71.1; 69 | NR | 10.9; 6.7 | 34 ± 14.3; 31.5 ± 16.6 | 449; 288 | NR |
| Meppelink et al.[71], The Netherlands | 9 (5M), 14 (11M); 5.7, 5.7 | 61.2; 64.6 | NR | 8.1; 8.7 | 21.4 ± 7; 20.4 ± 7.3 | 855; 794 | NR |

| | | | | | | | |
|---|---|---|---|---|---|---|---|
| Meral et al.[72] , Turkey | 17 (10M), 26 (16M); NR | 68.9; 66.7 | 63; 60.9 | 5.3; 5.8 | 20.1 ± 12; 15.8 ± 10.2 | 386.7; 307.9 | NR |
| Miloserdov et al.[73], Germany | 16 (11M), 16 (12M); 3.53, 3.59 | 70.5; 70.19 | NR | 9.78; 4.61 | 26.63 ± 8.67; 20.94 ± 10.64 | 797.98; 385.53 | NR |
| Morgante et al.[74], Italy | 37 (24M), 443 (276M); NR | 68.3; 66.1 | 62.4; 61.4 | 4.2; 3.3 | 16.6 ± 8.3; 18.1 ± 9.1 | 0.649; 0.571 | HDRS 5 (3.0-10.00); 5 (3.0-8.0) |
| Moustafa et al.[75] , Egypt | 21 (14M), 23 (16M); 14.1, 13.6 | 68.2; 66.7 | NR | 11.1; 9.61 | 21.8 ± 6.3; 20.8 ± 5.6 | 943.5; 883.1 | BDI 7.9 ± 2.3; 7.6 ± 1.5 |
| Muller et al. [76], Australia | 18 (12M), 15 (10M); 15, 13 | 67.1; 62.6 | 59.9; 55.6 | 5.8; 4.7 | 32 ± 34; 21 ± 41 | 708; 548 | BDI 9 ± 20; 6.5 ± 24 |
| Nagano-Saito et al.[77] , Japan | 8 (NR), 11 (NR); NR | 67.6; 66 | NR | 8.6; 5.1 | 43.1 ± 15.5; 39.2 ± 10.3 | 322; 255 | NR |
| *Nishio et al.[78], Japan<br>Group: Patients with kinetopsia | 24 (12M), 53 (22M); 13, 11.2 | 68.5; 65.7 | NR | 8.5; 6.7 | 23.1 ± 10.8; 16.2 ± 6.6 | 53.2; 436.4 | 8 people with depression; 6 people with depression |
| *Nishio et al.[78], Japan<br>Group: Patients with object misidentification illusions | 17 (7M), 53 (22M); 13.4, 11.2 | 67.3; 65.7 | NR | 6.5; 6.7 | 18.7 ± 10.8; 16.2 ± 6.6 | 516.3; 436.4 | 5 people with depression; 6 people with depression |
| *Nishio et al.[78], Japan<br>Group: Patients with VH | 19 (11M), 53 (22M); 12.7, 11.2 | 69.4; 65.7 | NR | 7.3; 6.7 | 27.2 ± 11; 16.2 ± 6.6 | 612.4; 436.4 | 5 people with depression; 6 people with depression |
| Oka et al.[79], Japan | 31 (20M), 37 (26M); NR | 68.5; 66.6 | NR | 4.9; 4.1 | 25.4 ± 6.1; 22.6 ± 5.2 | 276; 281 | NR |
| Ozer et al.[80], Turkey | 33 (20M), 30 (21M); NR | 67.39; 65.35 | 60.21; 59.57 | 6.79; 5.87 | 21.9 ± 11.5; 16.5 ± 11.1 | 413.79; 290.34 | NR |
| *Papapetropoulos et al.[81], Greece<br>Group: Patients with benign VH | 19 (14M), 104 (52M); NR | NR | 57.63; 59.68 | 8.55; 6.8 | 29.68 ± 17.1; 32.63 ± 21.8 | NR | NR |
| *Papapetropoulos et al.[81] , Greece<br>Group: Patients with malignant VH | 15 (7M), 104 (52M); NR | NR | 59.07; 59.68 | 13.33; 6.8 | 29.4 ± 16.7; 32.63 ± 21.8 | NR | NR |
| Papapetropoulos et al.[82], USA | 31 (21M), 39 (25M); 13.2, 12.8 | 63.9; 64.3 | 54.4; 55.8 | 9.5; 8.5 | 26.1 ± 8.1; 19.1 ± 10.3 | NR | BDI 17.9 ± 14.7; 15.1 ± 11.1 |
| Park et al.[83], South Korea | 7 (3M), 13 (8M); 9.1, 9.2 | 71; 66.3 | NR | 5.4; 5.1 | 26.8 ± 13.4; 22.1 ± 12.2 | 537.1; 616.9 | GDS 15.4 ± 8.1; 13.3 ± 7.7 |
| Pereira et al.[84], Spain | 18 (6M), 18 (6M); 7.5, 8.4 | 73.7; 73.8 | NR | 12.9; 12.8 | 28 ± 11.9; 25.4 ± 14.2 | 702.8; 671.2 | NR |
| Porter et al.[85], UK | 23 (9M), 102 (52M); NR | 75; 74.4 | NR | 8; 4.8 | 38.9 ± 13.61; 28.7 ± 12.4 | NR | HAD 12.6 ± 5.83; 10.1 ± 5.94 |
| Ramirez-Ruiz et al. [86], Spain | 24 (10M), 21 (9M); 7.3, 7.7 | 74.7; 73.3 | NR | NR | 30.6 ± 14.5; 24.9 ± 13.7 | NR | NR |

| | | | | | | | |
|---|---|---|---|---|---|---|---|
| Ramirez-Ruiz et al. [87], Spain | 18 (7M), 20 (8M); 7.6, 7.5 | NR | NR | 12.6; 10.6 | 29.3 ± 11.7; 24.5 ± 14 | 723.6; 647.5 | NR |
| Ramirez-Ruiz et al.[88], Spain | 10 (4M), 10 (4M); 7.8, 6.5 | 73; 72.5 | NR | 11.1; 11 | 28.8 ± 4.2; 26.8 ± 3.4 | 637.5; 585 | HAM-D 7.8 ± 1.5; 3.5 ± 1.1 |
| Sanchez-Ramos et al.[89], USA | 55 (40M), 158 (90M); NR | 70; 66 | NR | 8.6; 6.3 | NR | 426; 443 | NR |
| Santangelo et al.[90], Italy | 9 (NR), 15 (NR); 11.7, 10.3 | 72.1; 70.1 | NR | 10.5; 6.2 | 25.8 ± 11.6; 19 ± 9.8 | 844.8; 499.2 | NR |
| Sawada et al. [91], Japan | 28 (13M), 83 (33M); NR | 73.1; 68.6 | NR | 9; 7.7 | 28.78 ± 14.84; 20 ± 9.05 | 493.8; 414.8 | NR |
| Sawczak et al.[92], Canada | 30 (19M), 30 (19M); 15.6, 15.1 | 63.7; 62.7 | NR | 2.3; 1.7 | 25.7 ± 11.3; 21 ± 10 | 410.9; 370.2 | NR |
| Schumacher-Schuh et al.[93], Brazil | 50 (32M), 146 (68M); 5.8, 6.1 | 66.2; 68.5 | 56.3; 60.2 | 10; 8.3 | NR | 959.2; 740.7 | NR |
| Schumacher-Schuh et al. [94], Brazil | 54 (34M), 151 (71M); NR | NR | 56.58; 59.67 | 9.62; 8.13 | 41.26 ± 19.59; 32.81 ± 16.55 | 771.23; 677.35 | NR |
| Shin et al. [95], South Korea | 46 (23M), 64 (26M); 8.3, 8.4 | 71.3; 70.7 | NR | 0.275; 0.233 | 24.1 ± 10.4; 21.6 ± 11 | 482.4; 501.4 | NR |
| Shine et al. [96], Australia | 23 (NR), 22 (NR); NR | 67.3; 60.1 | NR | 9; 5.6 | 32.3 ± 16.8; 27.1 ± 16.1 | 644.3; 542 | BDI-II 9.6 ± 6.5; 11.5 ± 9.7 |
| Shine et al.[97], Australia | 9 (NR), 13 (NR); NR | 65.7; 61.5 | NR | 6.7; 4.9 | 27.6 ± 14.2; 21.1 ± 9.1 | 1081.9; 1264 | BDI-II 9.2 ± 6.1; 8 ± 7.4 |
| Shine et al.[98], Australia | 10 (NR), 9 (NR); NR | 69.5; 67.1 | NR | 6.9; 4.4 | 34 ± 15; 32 ± 15 | 819.5; 512.5 | BDI-II 15.5 ± 14; 8.8 ± 7 |
| Shine et al.[99], Australia | 86 (NR), 111 (NR); 14.1, 14 | 70.5; 68.6 | NR | 7.7; 7.2 | 34.5 ± 18; 30.3 ± 15.6 | 749.6; 720.6 | BDI-II 11.3± 10.1; 7.5 ± 6.6 |
| Shine et al.[100], USA | 21 (14M), 14 (10M); NR | 69.3; 66.3 | NR | 6; 4.7 | 40.3 ± 9; 34.9 ± 8 | 960.2; 1116.3 | BDI-II 14.1 ± 11; 18.9 ± 11 |
| Stebbins et al. [101], USA | 12 (NR), 12 (NR); NR | 71.08; 73.25 | NR | 13.92; 11.17 | 30.42 ± 10.71; 31.8 ± 14.81 | NR | NR |
| Straughan et al. [102], UK | 16 (10M), 20 (10M); 10.06, 10.05 | 71.69; 71.1 | NR | 8.45; 5.94 | 31.31 ± 13.17; 19.05 ± 6.81 | NR | NR |
| Thota et al.[103], India | 34 (30M), 35 (28M); NR | 58.7; 55.7 | 51.4; 50 | NR | 13.2 ± 5.4; 12.4 ± 6.6 | 736; 625.2 | NR |
| Uchiyama et al. [104], Japan | 11 (5M), 42 (17M); 13.5, 12.3 | 68.3; 65.5 | NR | 6.2; 6.3 | 21.2 ± 2.8; 16.4 ± 1.2 | 535.5; 368.6 | NR |
| Weintraub et al.[105], USA | 34 (NR), 96 (NR); 14.2, 14.6 | 69.9; 71.9 | NR | 8.5; 6.5 | 24.8 ± 11; 22.1 ± 11.2 | 578.5; 392.3 | NR |
| Yao et al.[106], UK | 12 (10M), 15 (10M); 7.2, 6.3 | NR | NR | 9.1; 7.1 | 20.704 ± 17.82; 20.99 ± 13.9 | 986.9; 689.7 | MADRS-S 1.64 ± 2.31; 0.09 ± 0.20 |
| Zarkali et al.[107], UK | 19 (6M), 86 (49M); 17.1, 17.1 | 64.6; 64.5 | NR | 4.8; 4.1 | 29.2 ± 20.8; 22.2 ± 11.5 | 434.9; 460.5 | HADS-D 4.8 ± 3.2; 3.8 ± 2.9 |

| | | | | | | | |
|---|---|---|---|---|---|---|---|
| Zhu et al.[108], China | 72 (49M), 299 (199M); NR | 67.75; 64.57 | 60; 59.06 | 7.74; 5.58 | 29.67 ± 18.53; 21.31 ± 12.24 | 580.63; 493.52 | HAM-D 15.97 ± 10.36; 11.83 ± 8.96 |

*Studies with more than one group of PDP psychosis patients, e.g., PDP patients with visual hallucinations with/without delusions.

BDI: Beck Depression Inventory; CES-D: Centre for Epidemiologic Studies depression self-rating scale; GDS: Geriatric Depression Scale; HADS-D: Hospital Anxiety and depression scale; HAM-D: Hamilton Depression Scale; HDRS: Hamilton Depression Rating Scale; IDRS: Inventory for Depressive Symptomatology score; MADRS: Montgomery-Asberg Depression Rating Scale; MDS-UPDRS: Movement disorders society – unified Parkinson's Disease Rating Scale; NPI: Neuropsychiatric Inventory; NR: Not reported; SDS: Self-rating depression scale; VH: visual hallucinations.

#### *Study characteristics*

The majority of the studies included PD patients with visual hallucinations (n= 94.3%). One study examined pareidolia in PD patients [104], eight studies included PD patients with different degrees of psychosis severity, i.e., PD patients with psychosis [4, 24], PDP patients experiencing hallucinations with and without insight [66], PDP patients with externally or internally derived hallucinations [13], PDP patients with different variations of hallucinations and illusions [53, 78], PDP patients divided based on whether they developed psychosis early or late in their PD diagnosis [40], and PDP patients with benign or malignant visual hallucinations [81]. Two studies employed a previous version of the Unified Parkinson's Disease Rating Scale (UPDRS) part I which included an item on thought disorders [4, 21]. The question probes the presence of a continuum of psychosis phenomena from mild to severe hallucinations and delusions. Diagnosis of PDP relied on neuropsychiatric scales (e.g., Neuropsychiatric Inventory, NPI; Scales for Outcomes in Parkinson's Disease – Psychiatric Complications, SCOPA-PC; UPDRS part I question 1.2), interview and on defined clinical criteria.

Overall, the majority of the studies, 50% received a global rating of "Strong", 35% received a global rating of "Moderate", and 15% of "Weak". Quality ratings are presented using the Robvis tool which was adapted to match the methodological dimensions of the EPHPP rating tool. The main methodological weaknesses were reported in the "Confounder" domain whereby some studies did not report the factors controlled for in their analyses, however most studies used a match design approach to control for demographics and other relevant clinical variables between PDP and PDnP patients. Full quality ratings are reported in eFigure1 A-C.

### Supplementary Material 3

eFigure 1 A and B. Quality ratings plots reporting the rating for each domain based on the EPHPP tool. Methodological domains are “Selection bias” (D1), “Study design” (D2), “Confounder” (D3), “Data collection” (D4), and “Withdrawals” (D5). The overall column reports the global rating for each study. “Strong”, “Moderate” or “Weak” are assigned to each domain. C. Summary of quality rating across the domains.

A

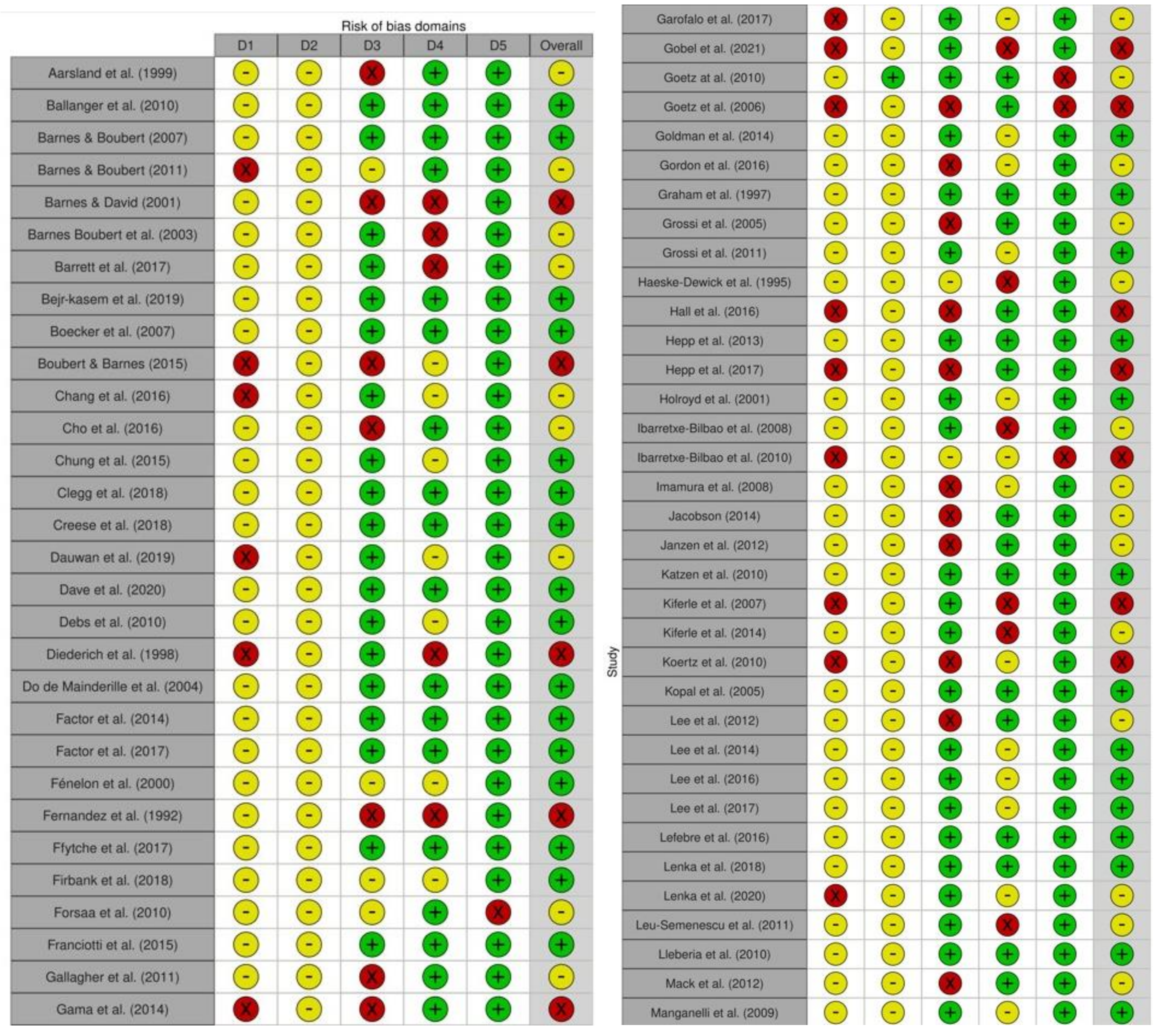

B

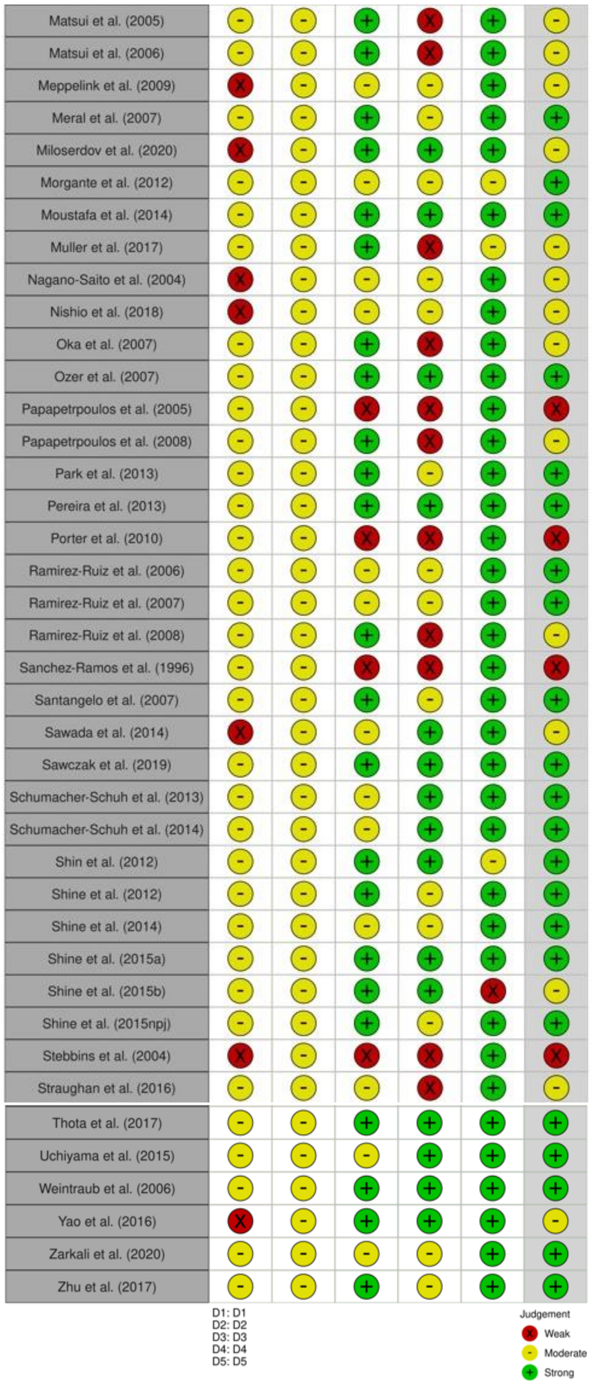
Matsui et al. (2005)
Matsui et al. (2006)
Meppelink et al. (2009)
Meral et al. (2007)
Miloserdov et al. (2020)
Morgante et al. (2012)
Moustafa et al. (2014)
Muller et al. (2017)
Nagano-Saito et al. (2004)
Nishio et al. (2018)
Oka et al. (2007)
Ozer et al. (2007)
Papapetrpoulos et al. (2005)
Papapetrpoulos et al. (2008)
Park et al. (2013)
Pereira et al. (2013)
Porter et al. (2010)
Ramirez-Ruiz et al. (2006)
Ramirez-Ruiz et al. (2007)
Ramirez-Ruiz et al. (2008)
Sanchez-Ramos et al. (1996)
Santangelo et al. (2007)
Sawada et al. (2014)
Sawczak et al. (2019)
Schumacher-Schuh et al. (2013)
Schumacher-Schuh et al. (2014)
Shin et al. (2012)
Shine et al. (2012)
Shine et al. (2014)
Shine et al. (2015a)
Shine et al. (2015b)
Shine et al. (2015npj)
Stebbins et al. (2004)
Straughan et al. (2016)
Thota et al. (2017)
Uchiyama et al. (2015)
Weintraub et al. (2006)
Yao et al. (2016)
Zarkali et al. (2020)
Zhu et al. (2017)
D1: D1
D2: D2
D3: D3
D4: D4
D5: D5
Judgement
Weak
Moderate
Strong

C

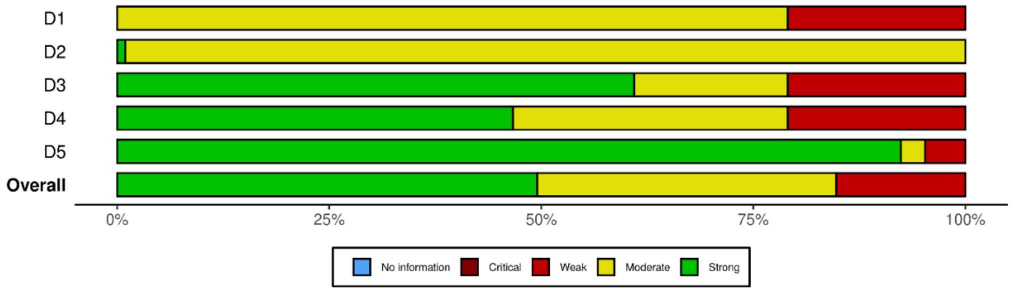
D1
D2
D3
D4
D5
Overall
0%
25%
50%
75%
100%
No information
Critical
Weak
Moderate
Strong

### Supplementary Material 4

Forest plots showing posterior probability distribution for standard mean difference (expressed in Hedges' *g*) and funnel plots for publication bias for:

*Global cognition*

*eFigure2. Posterior probability forest plot for global cognition*

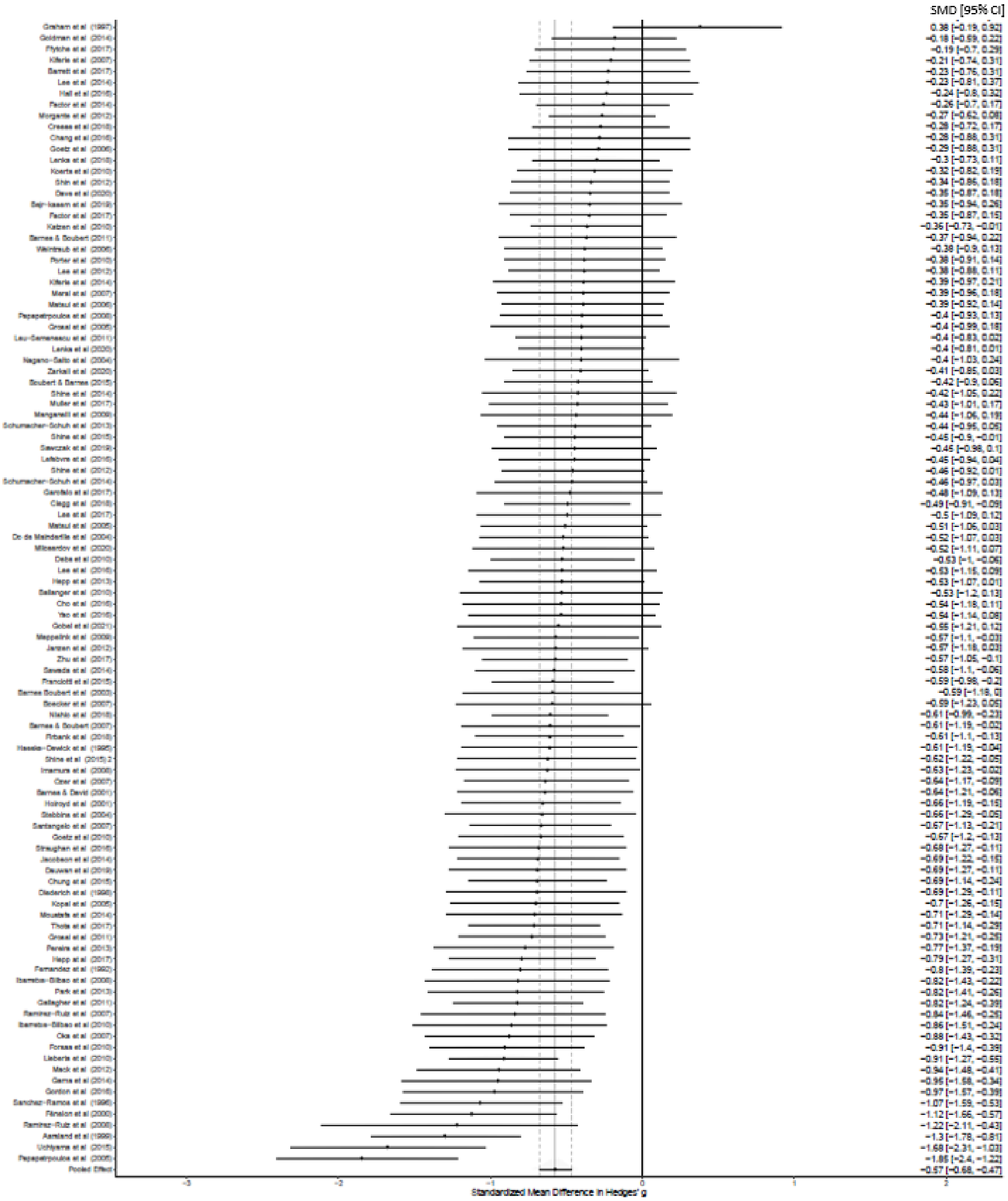

*eFigure3. Funnel plot for global cognition*

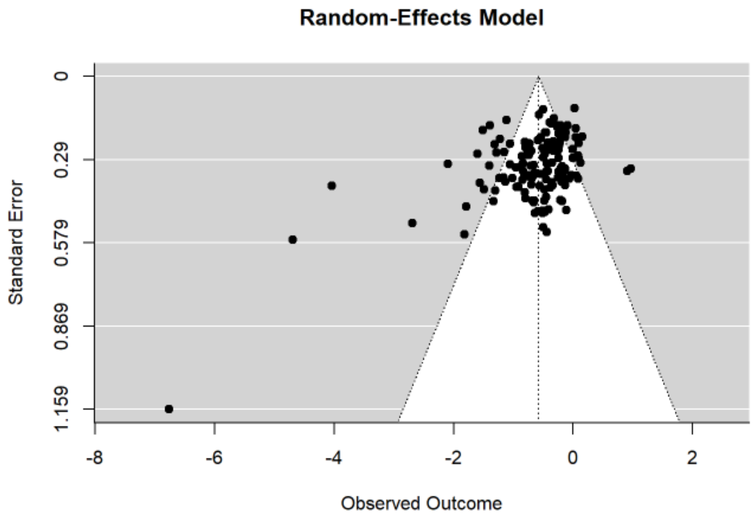

Publication bias, t = -10.963, p <0.001

*Construction*

*eFigure4. Posterior probability forest plot for construction*

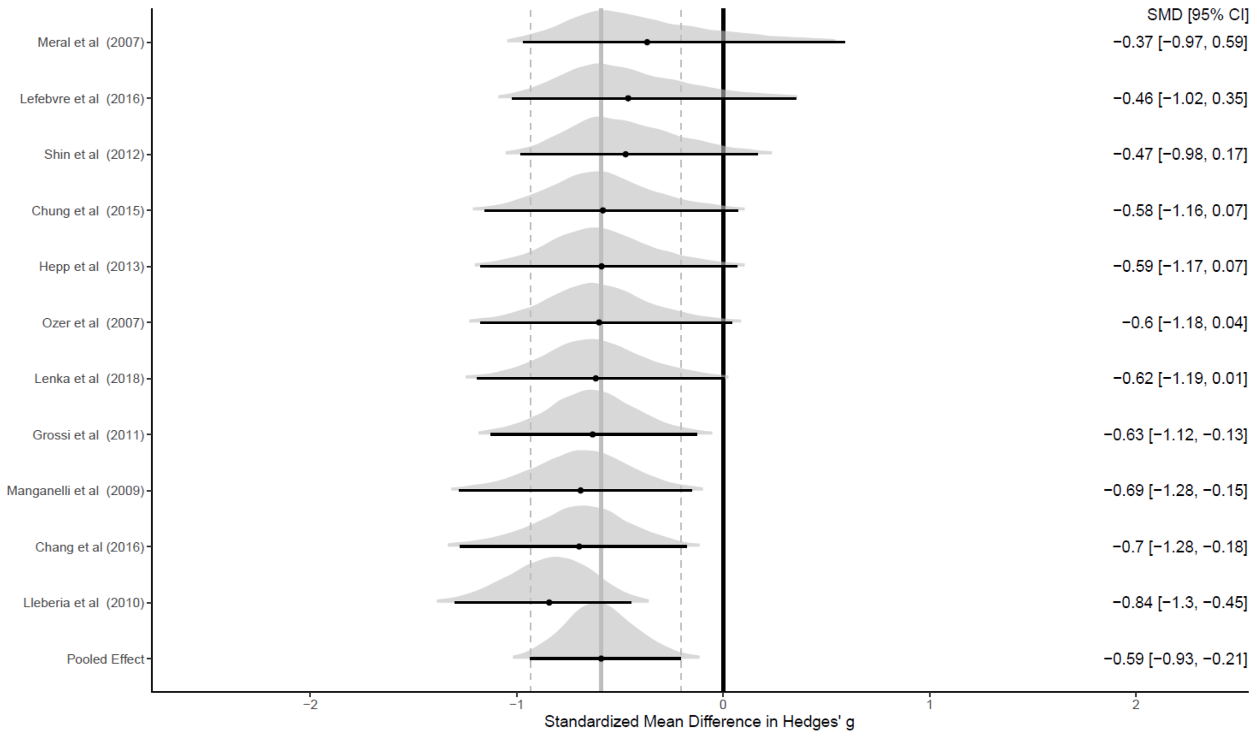

*eFigure5. Funnel plot for construction*

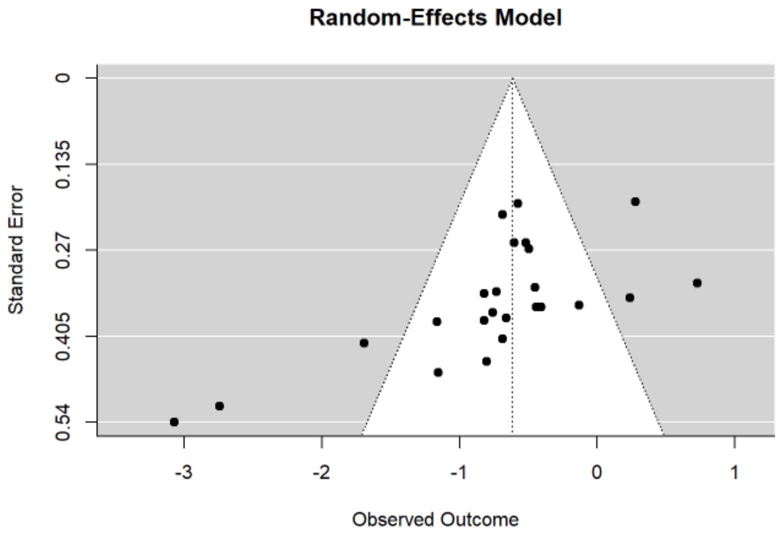

Publication bias, t = -14.893, p = 0

*Construction – Drawing*

*eFigure6. Posterior probability forest plot for drawing (construction)*

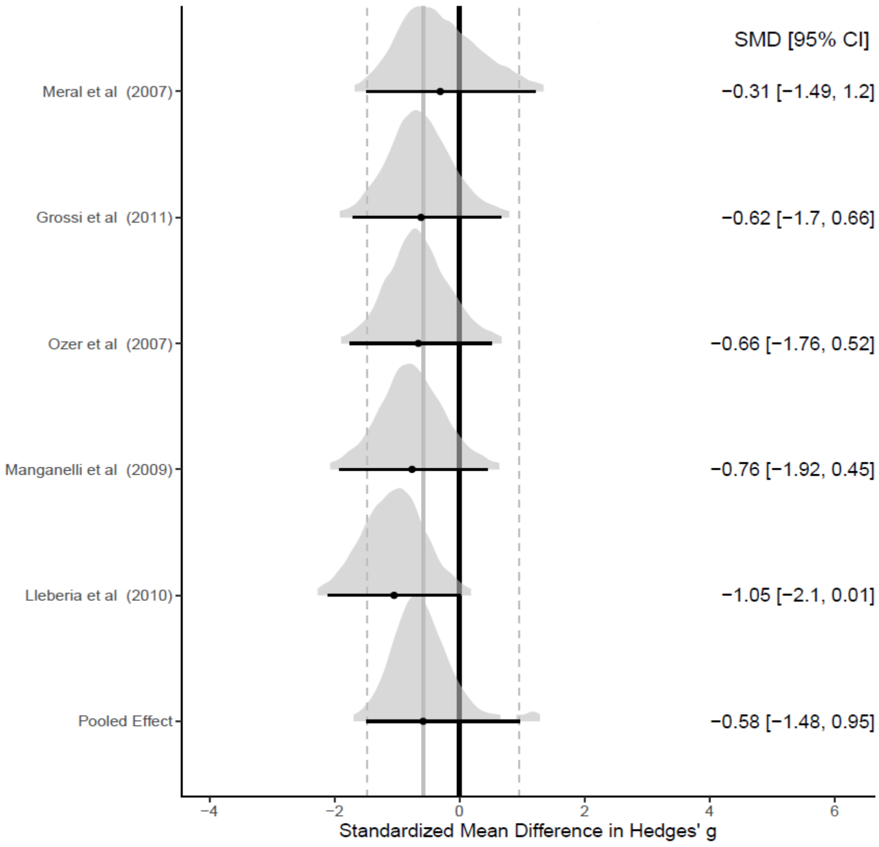

*eFigure7. Funnel plot for drawing (construction)*

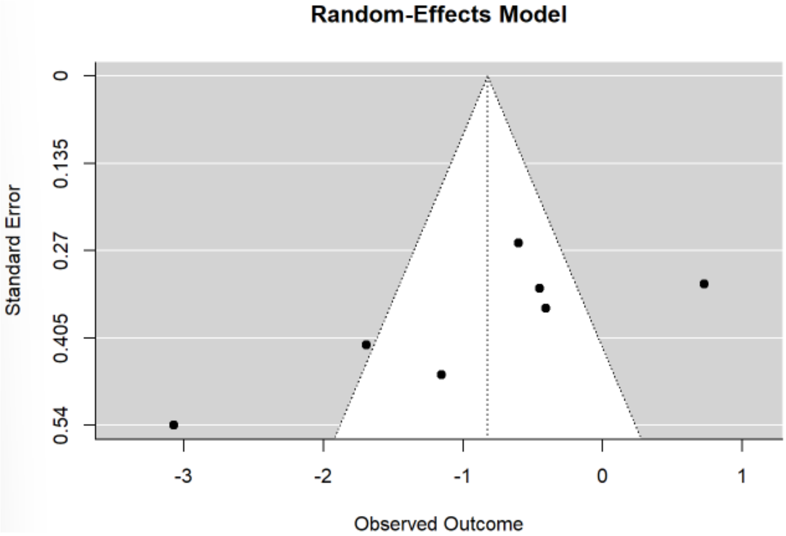

Publication bias, t = -4.744, p < 0.001

*Construction – Copying*

*eFigure8. Posterior probability forest plot for copying (construction)*

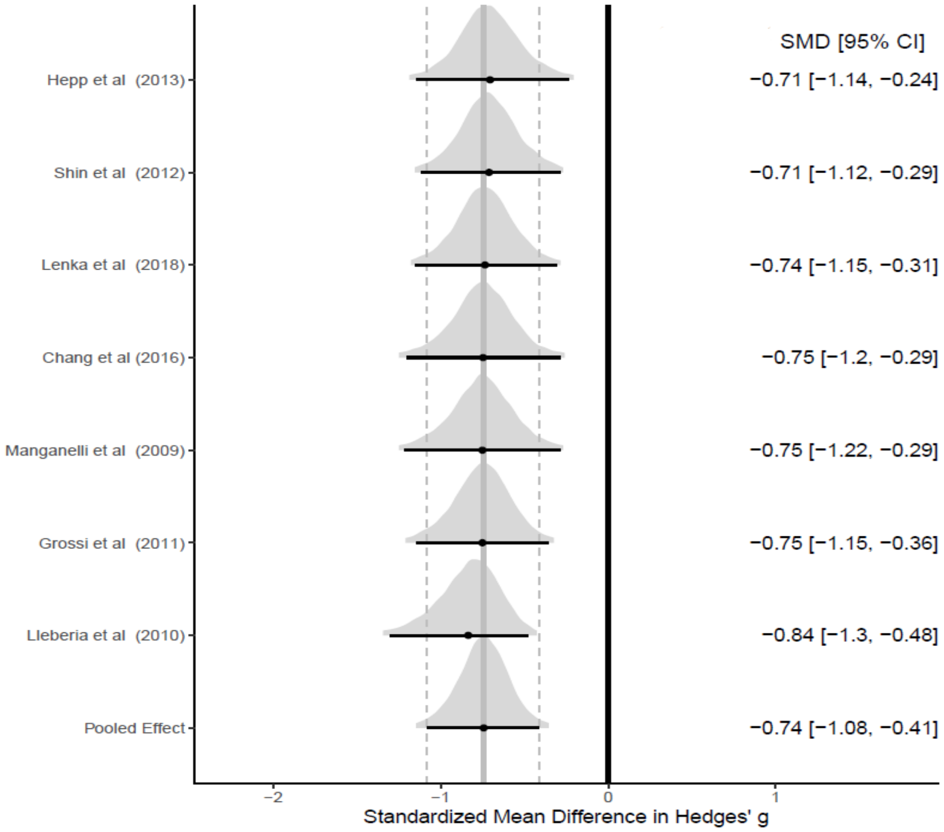

*eFigure9. Funnel plot for copying (construction)*

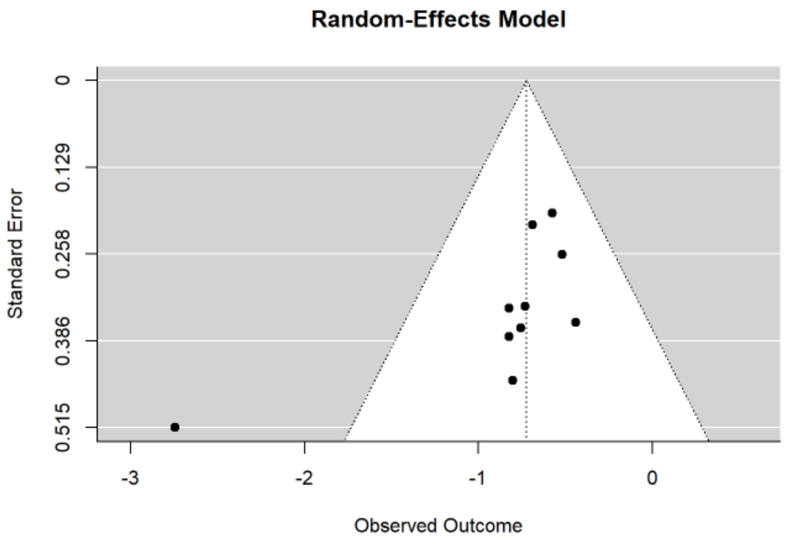

Publication bias, t = -3.514, p < 0.001

*Working memory*

*eFigure10. Posterior probability forest plot for working memory*

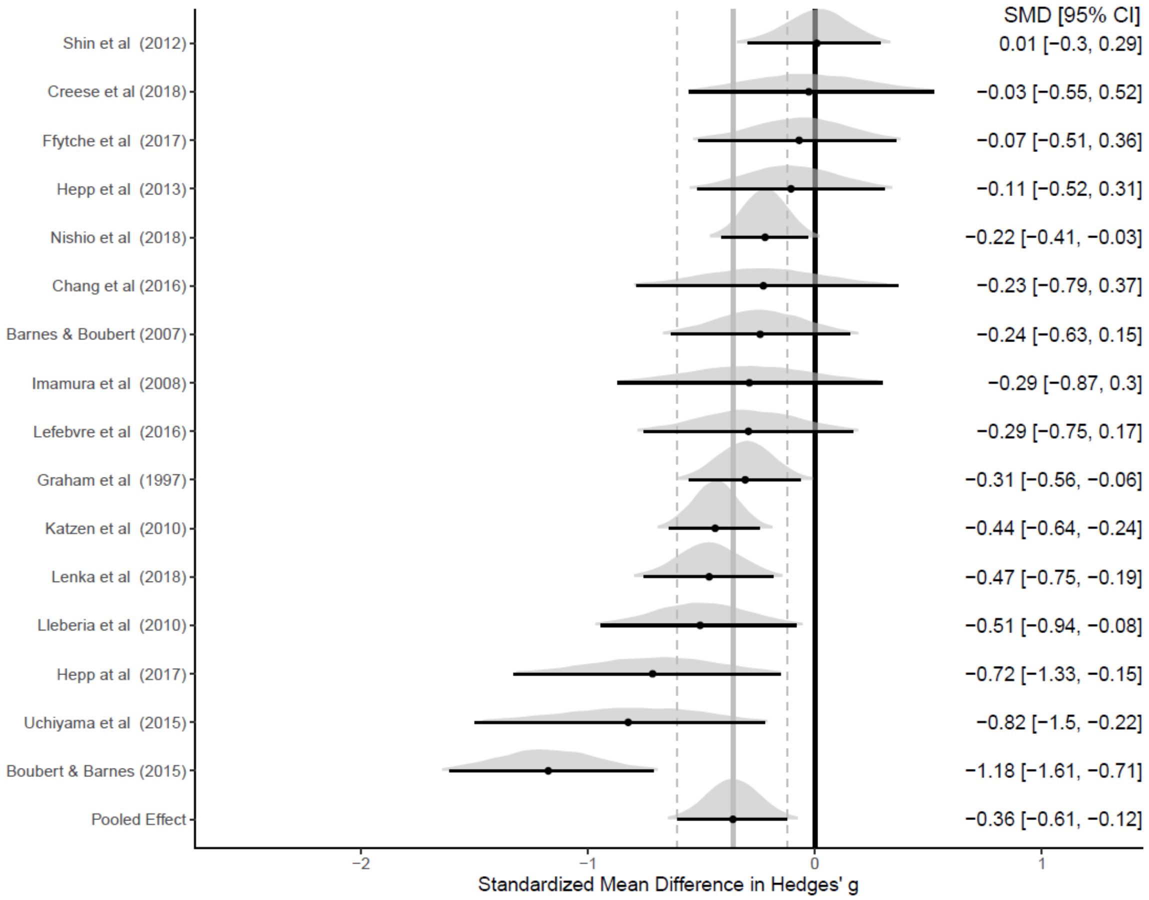

*eFigure11. Funnel plot for working memory*

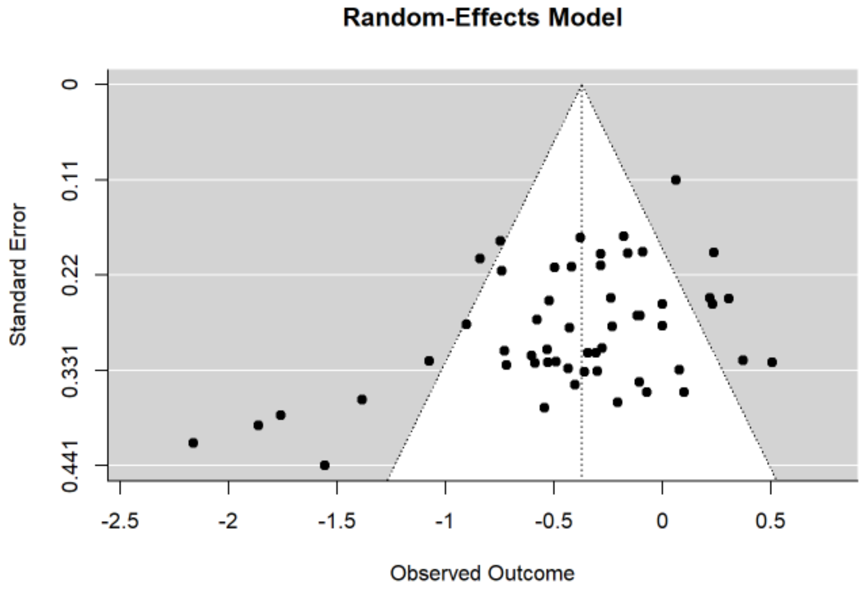

Publication bias, t = -4.400, p < 0.001

*Working memory – maintenance*

*eFigure12. Posterior probability forest plot for maintenance (working memory)*

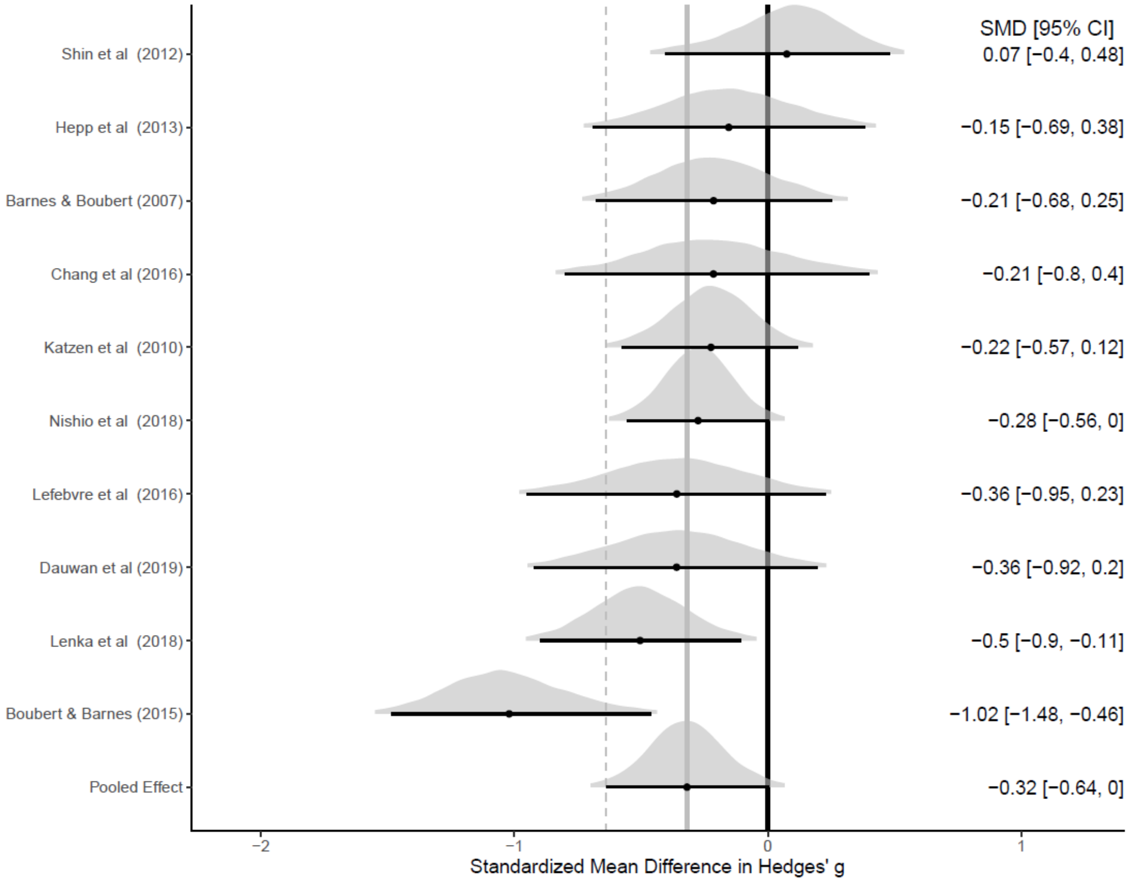

*eFigure13. Funnel plot for maintenance (working memory)*

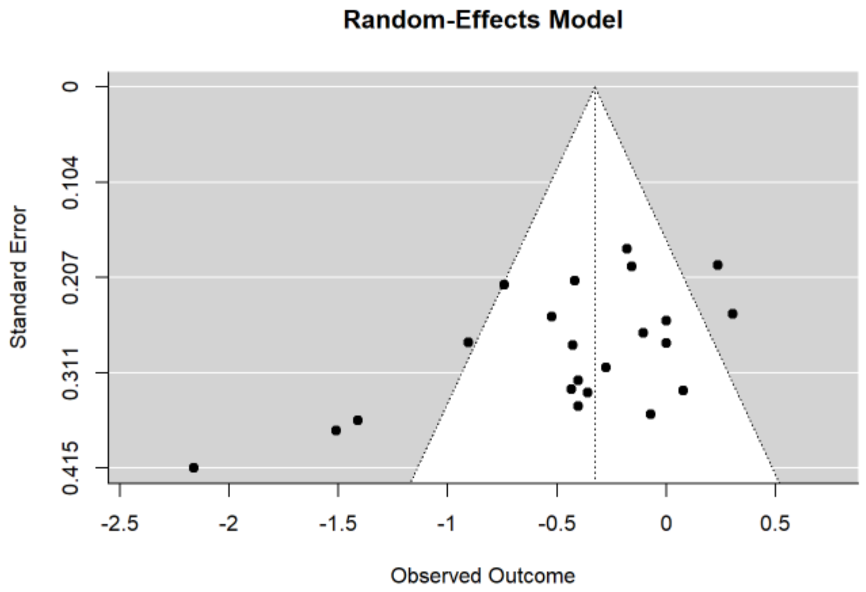

Publication bias, t = -3.11, p = 0.002

*Working memory – manipulation*

*eFigure14. Posterior probability forest plot for manipulation (working memory)*

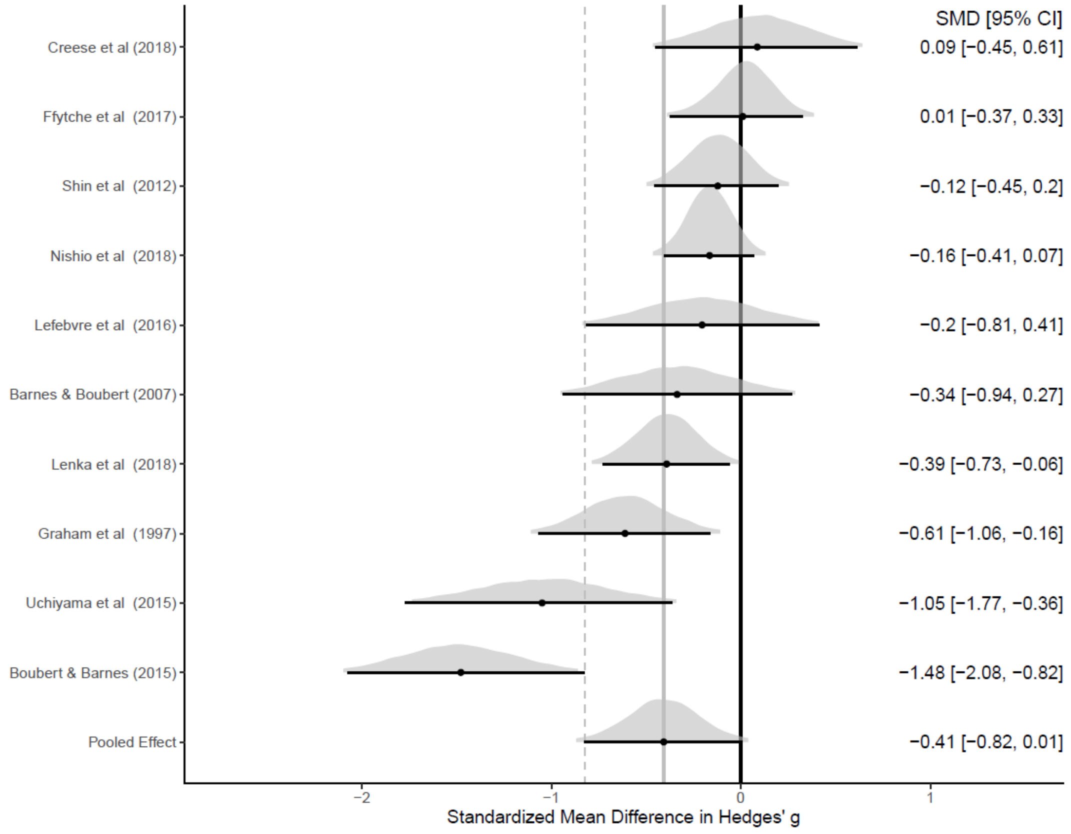

*eFigure15. Funnel plot for manipulation (working memory)*

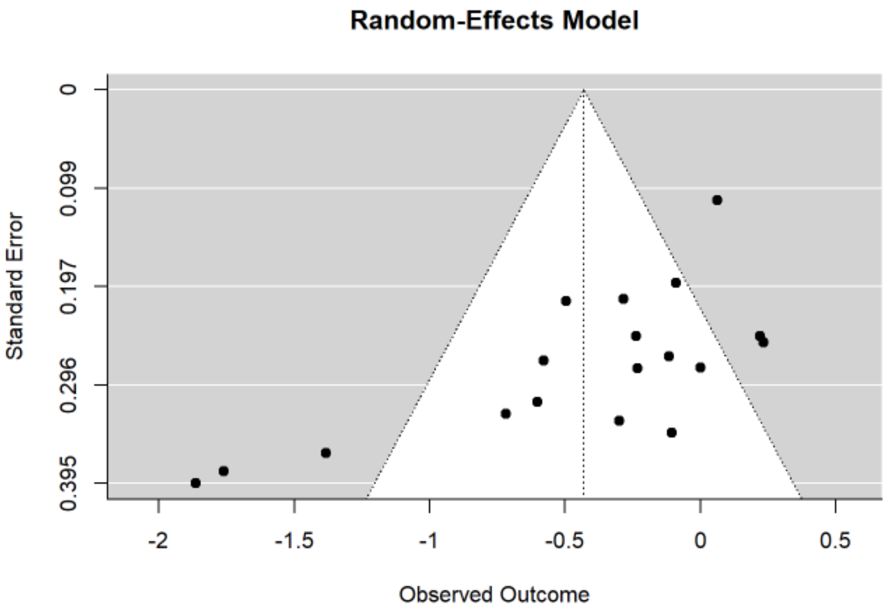

Publication bias, t = -2.907, p = 0.003

*Episodic memory*

*eFigure16. Posterior probability forest plot for episodic memory*

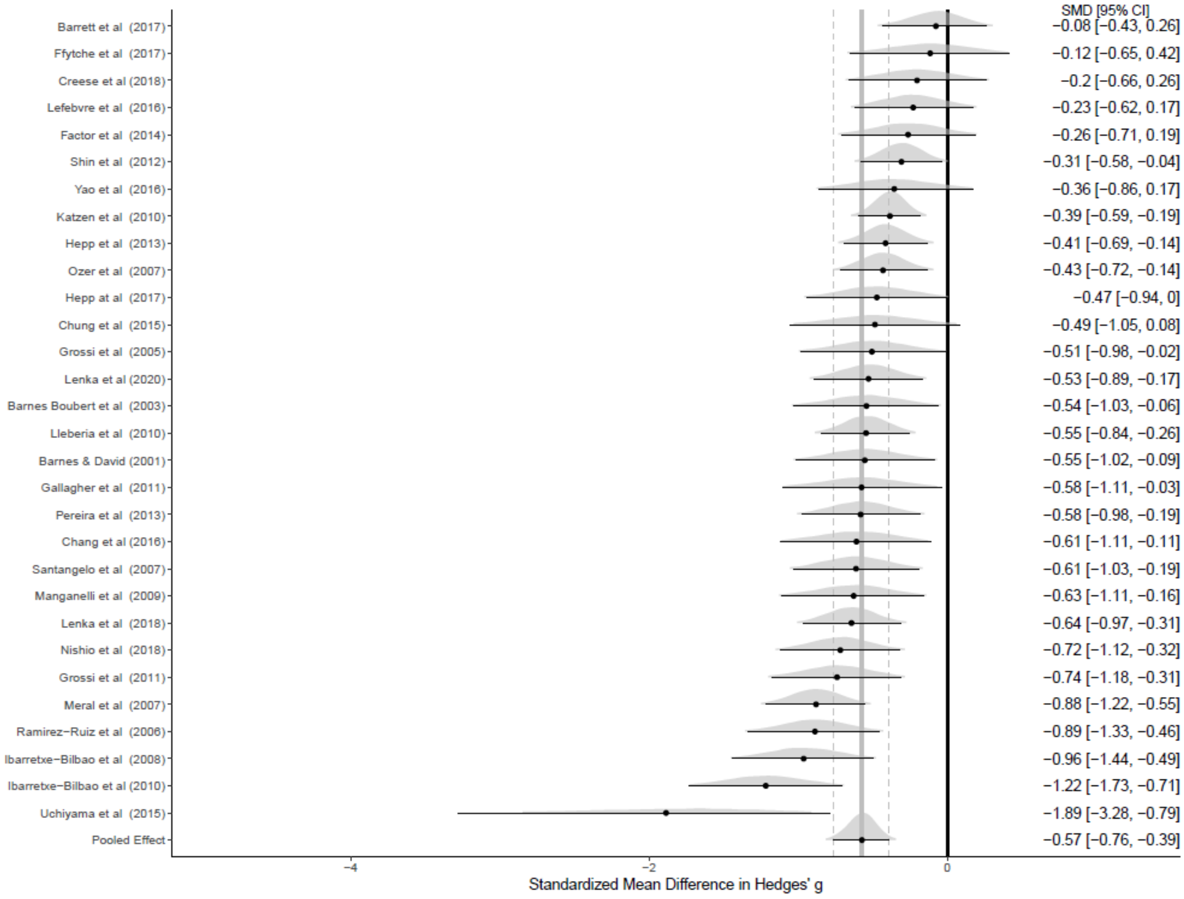

*eFigure17. Funnel plot for episodic memory*

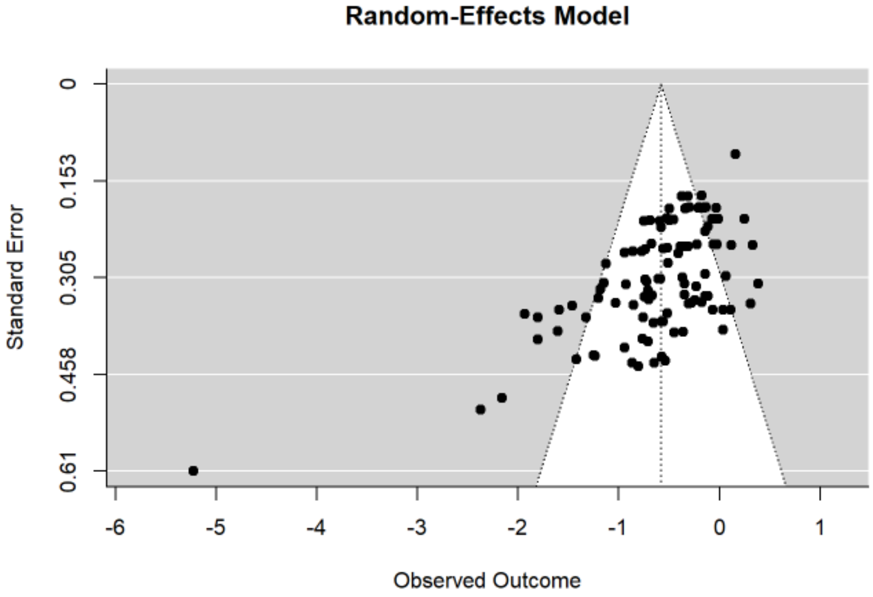

Publication bias, t = -10.995, p <0.001

*Episodic memory – nonverbal retrieval*

*eFigure18. Posterior probability forest plot for nonverbal retrieval (episodic memory)*

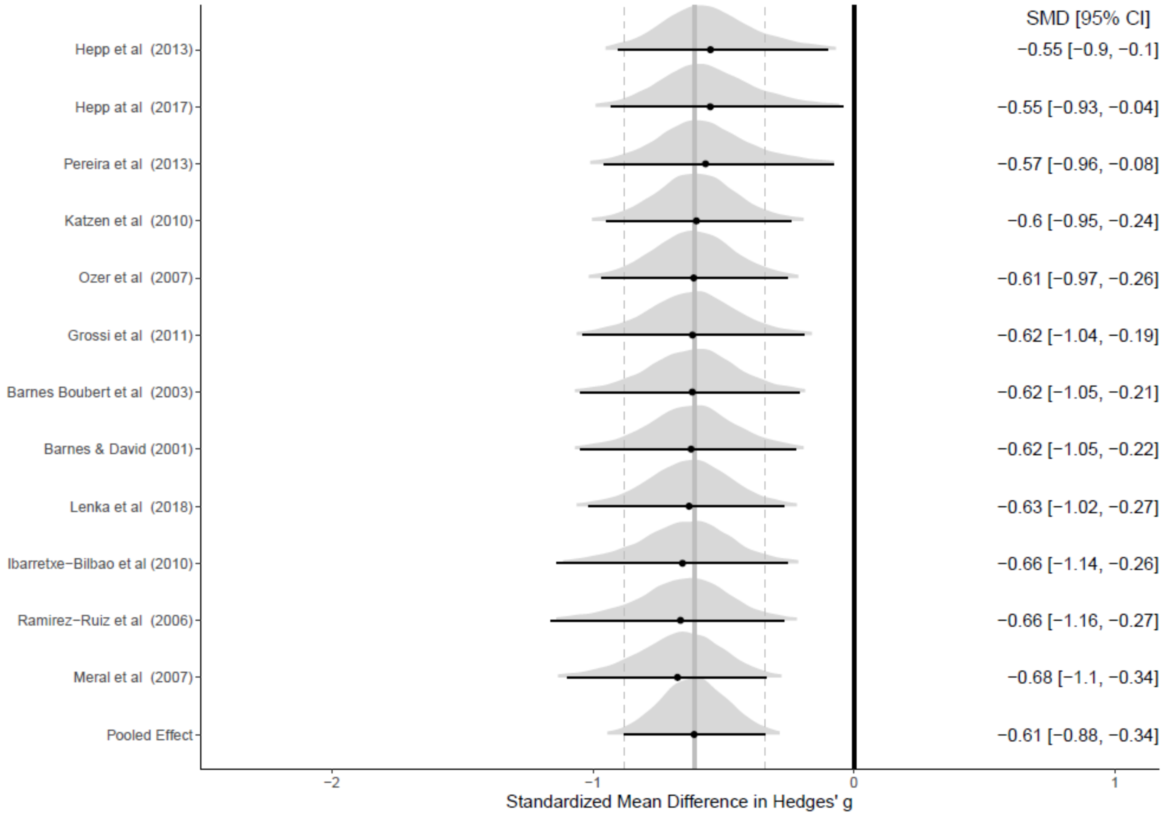

*eFigure19. Funnel plot for nonverbal retrieval (episodic memory)*

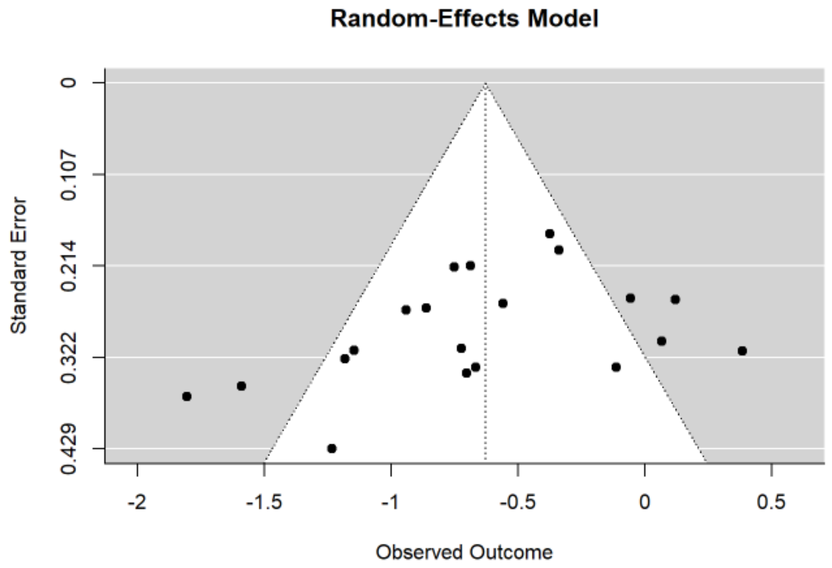

Publication bias, t = -2.390, p = 0.017

*Episodic memory - Verbal retrieval*

*eFigure20. Posterior probability forest plot for verbal retrieval (episodic memory)*

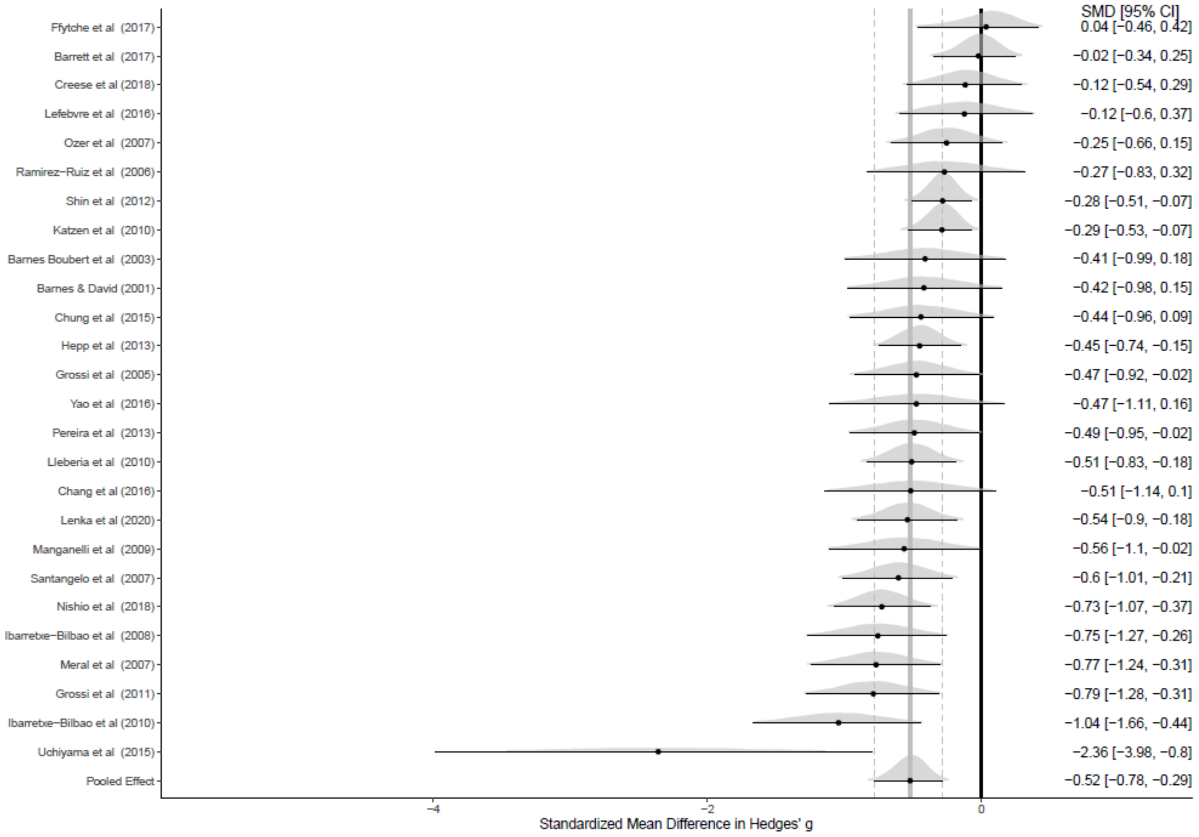

*eFigure21. Funnel plot for verbal retrieval (episodic memory)*

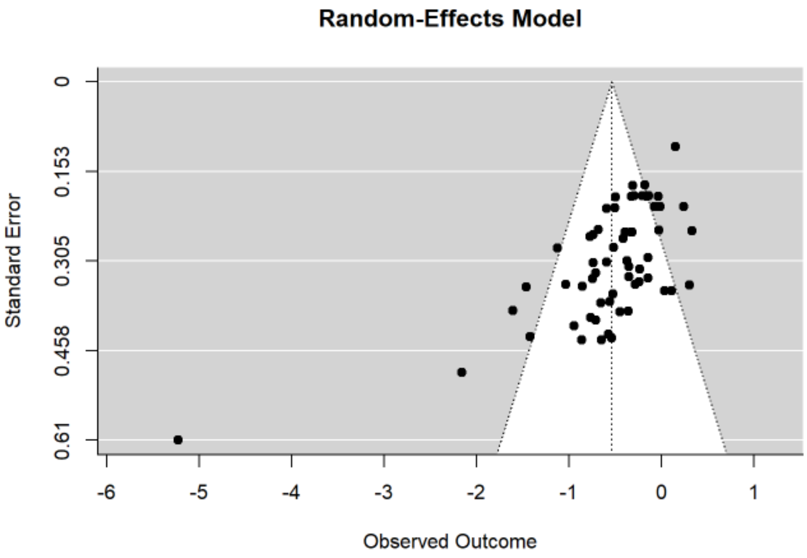

Publication bias, t = -8.886, p < 0.001

*Episodic memory – Encoding*

*eFigure22. Posterior probability forest plot for encoding (episodic memory)*

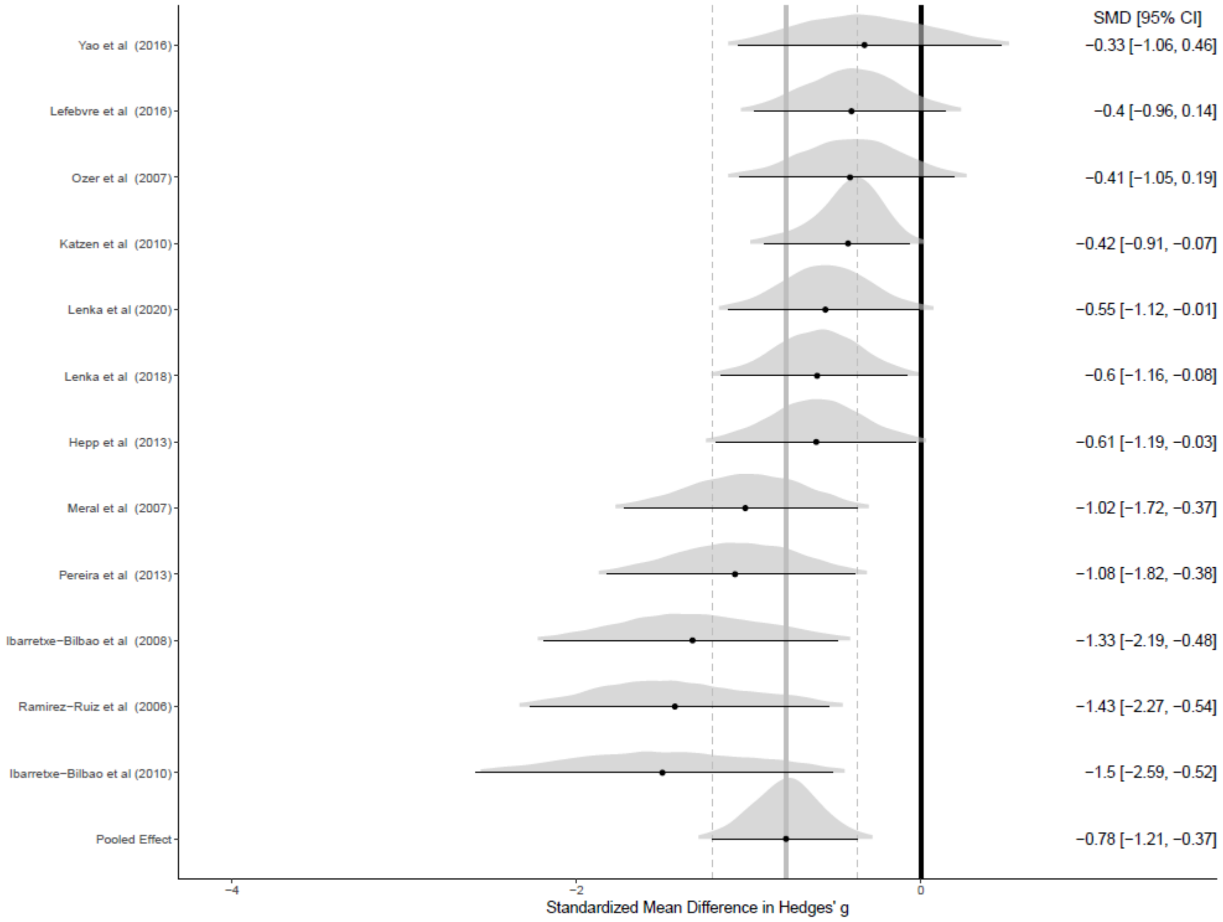

*eFigure23. Funnel plot for encoding (episodic memory)*

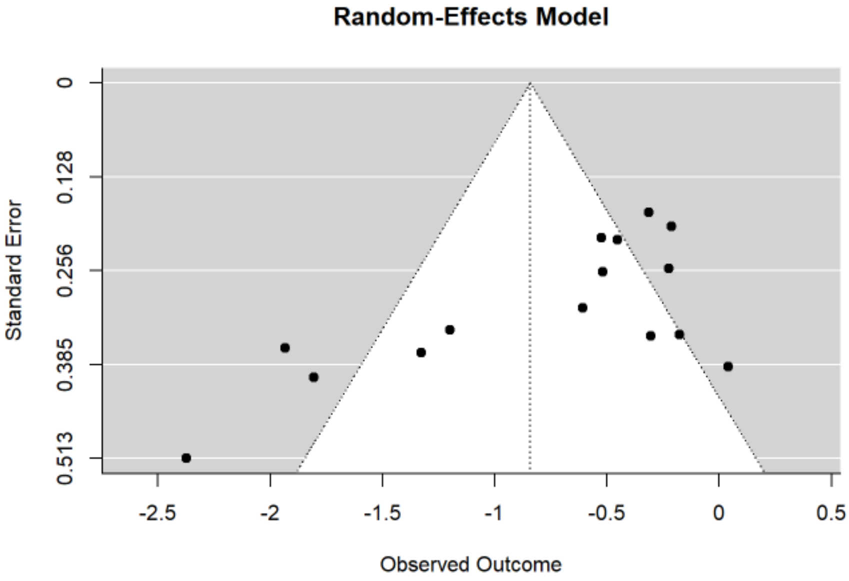

Publication bias, t = -3.619, p < 0.001

*Language*

*eFigure24. Posterior probability forest plot for language*

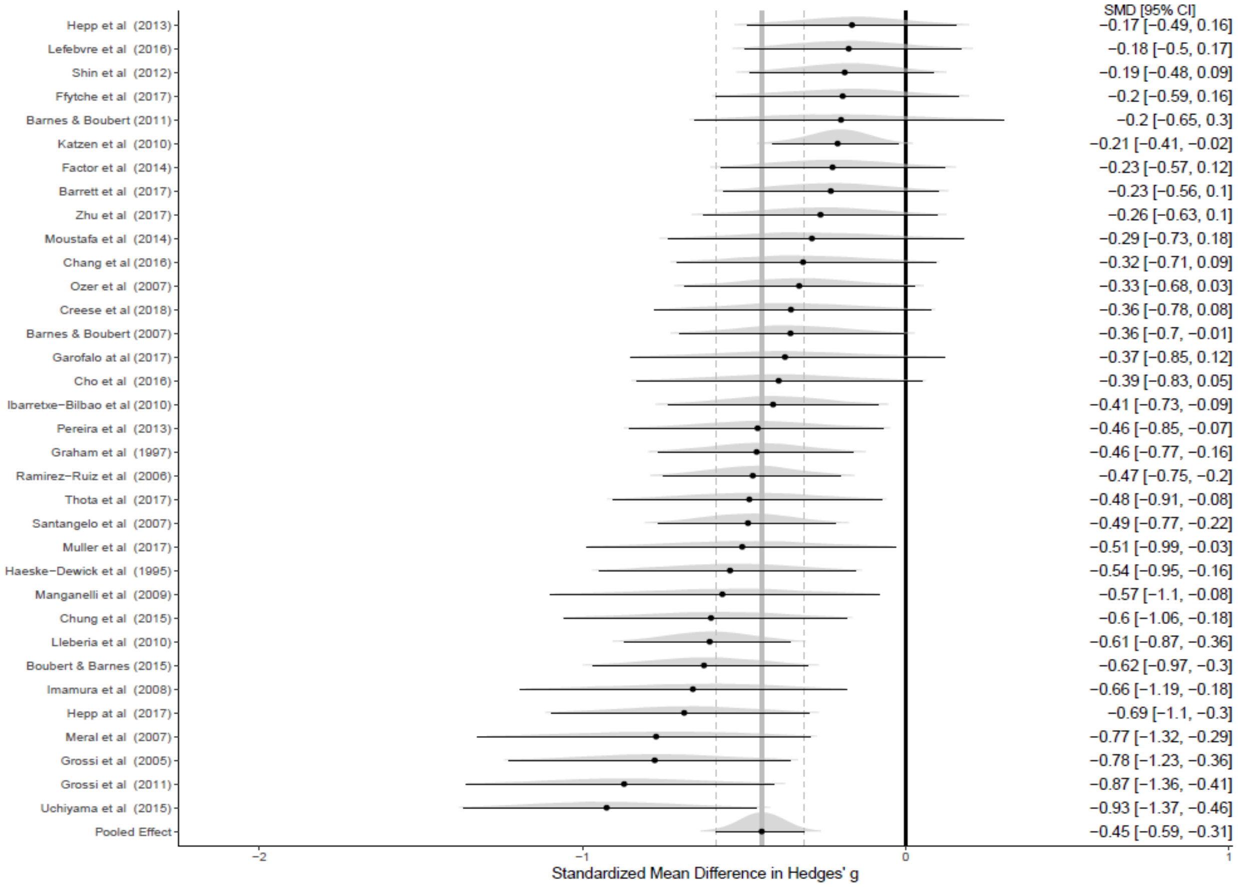

*eFigure25. Funnel plot for language*

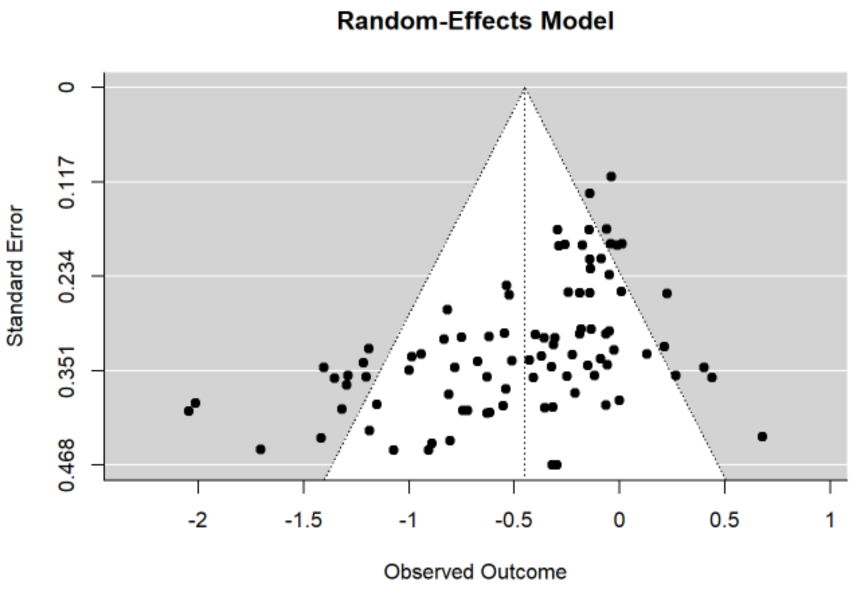

Publication bias, t = -4.298, p < 0.001

*Language - Semantic fluency*

*eFigure26. Posterior probability forest plot for semantic fluency (language)*

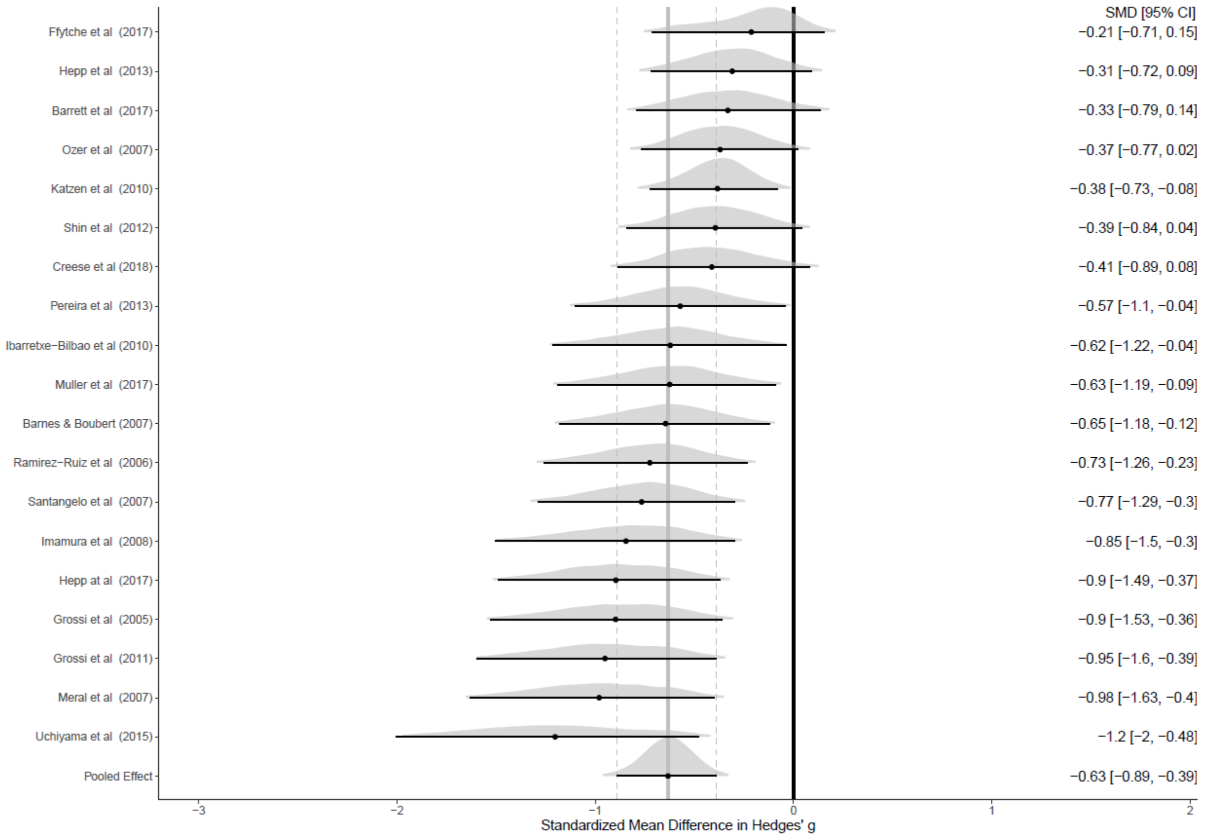

*eFigure27. Funnel plot for semantic fluency (language)*

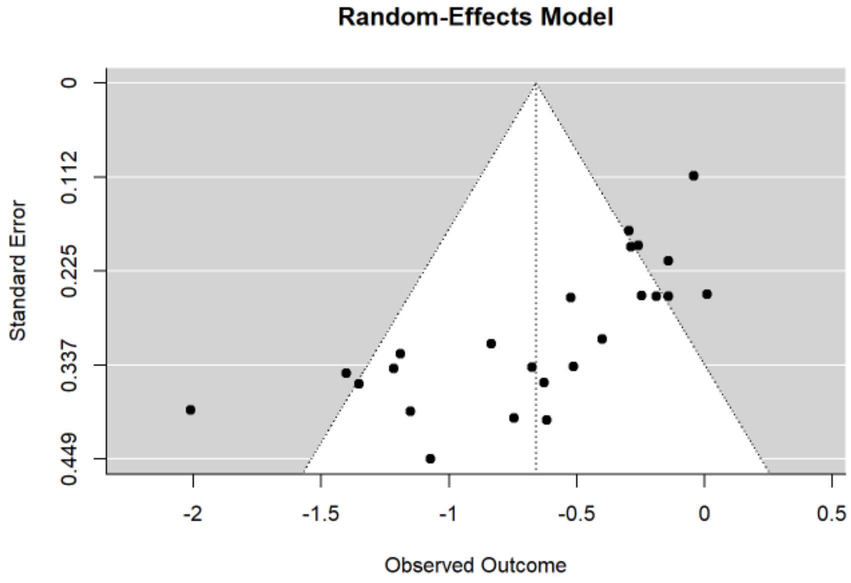

Publication bias, t = -3.961, p < 0.001

*Language - Phonemic fluency*

*eFigure28. Posterior probability forest plot for phonemic fluency (language)*

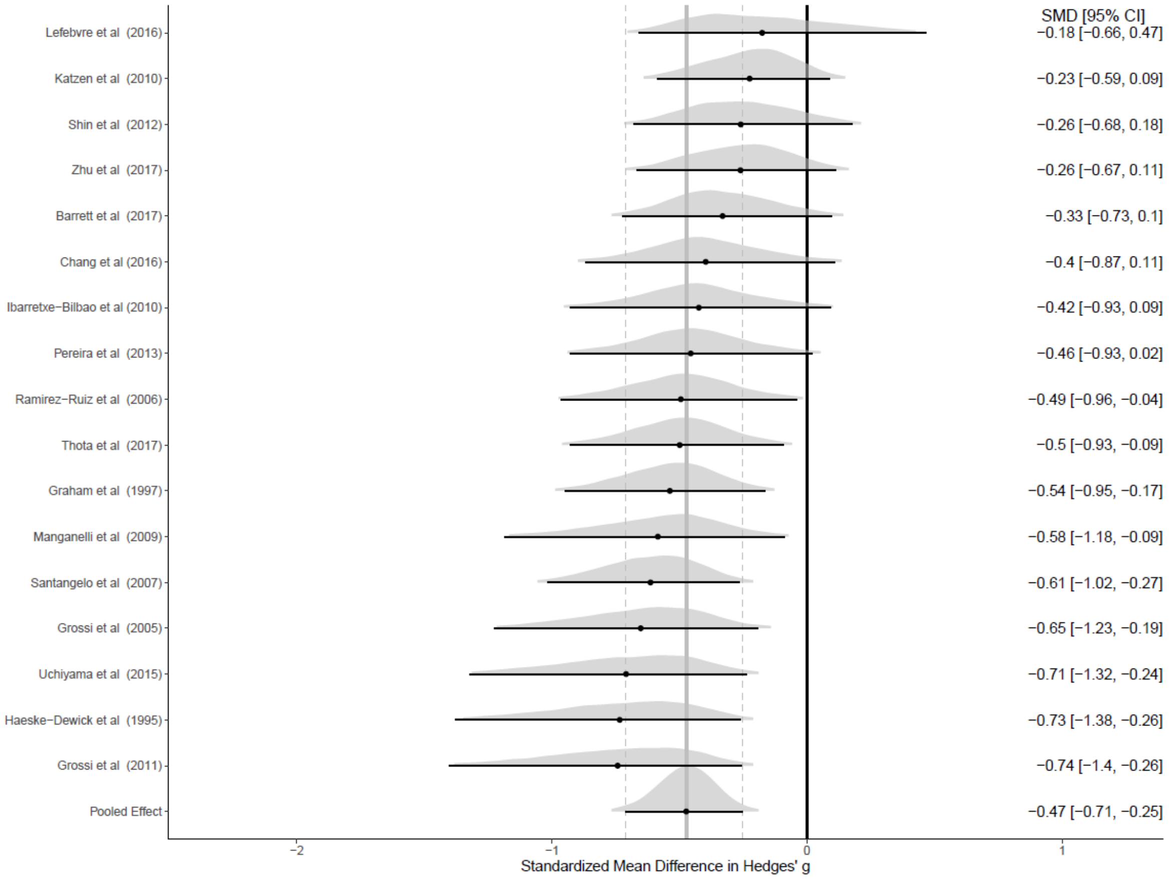

*eFigure29. Funnel plot for phonemic fluency (language)*

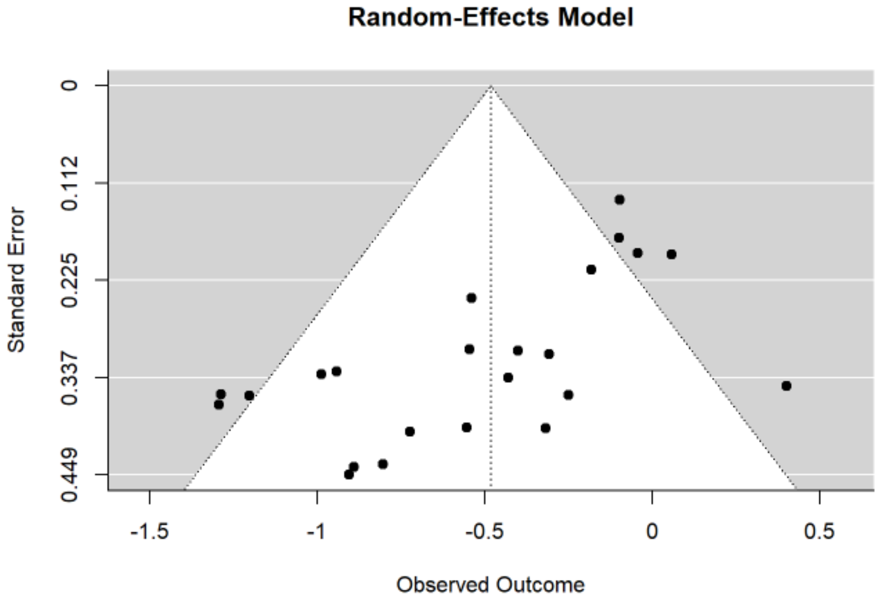

Publication bias, t = -2.952, p = 0.003

*Language – Naming*

*eFigure30. Posterior probability forest plot for naming (language)*

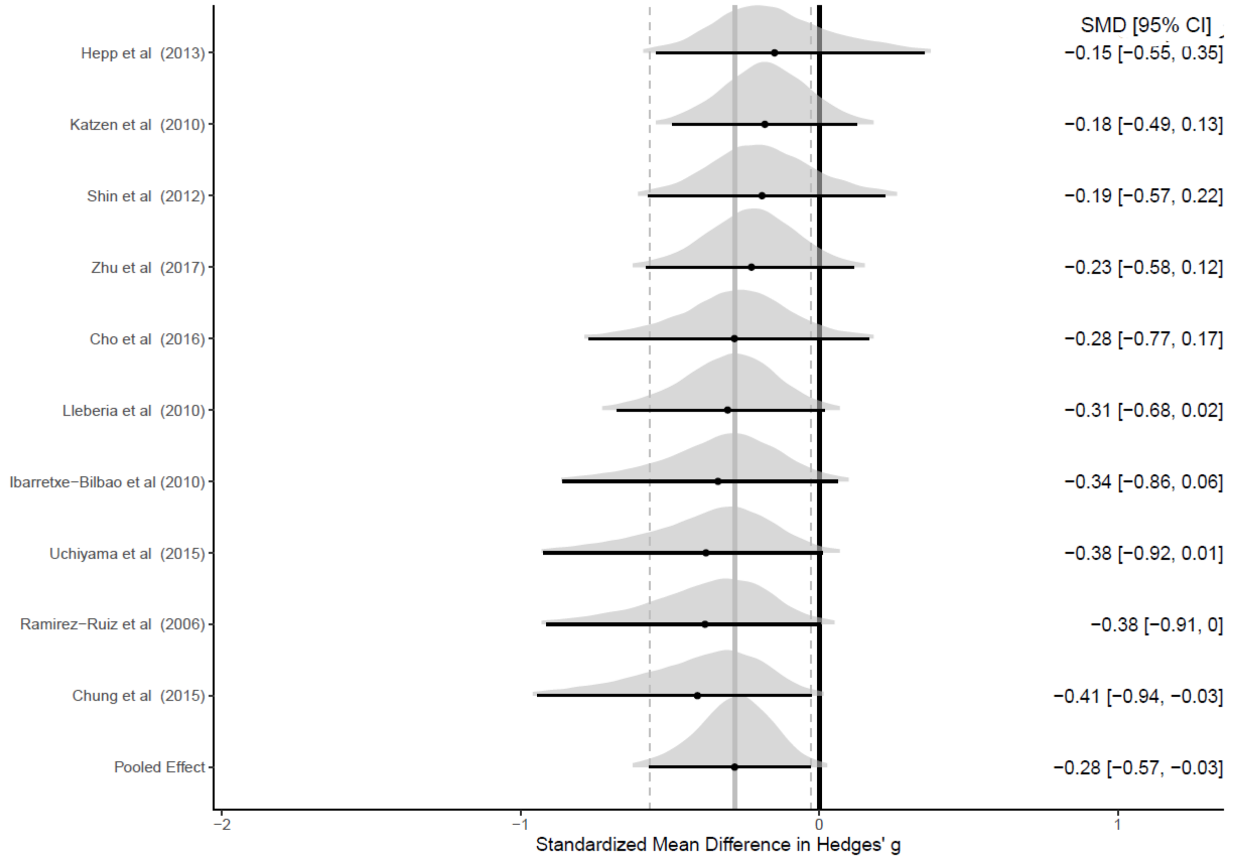

*eFigure31. Funnel plot for naming (language)*

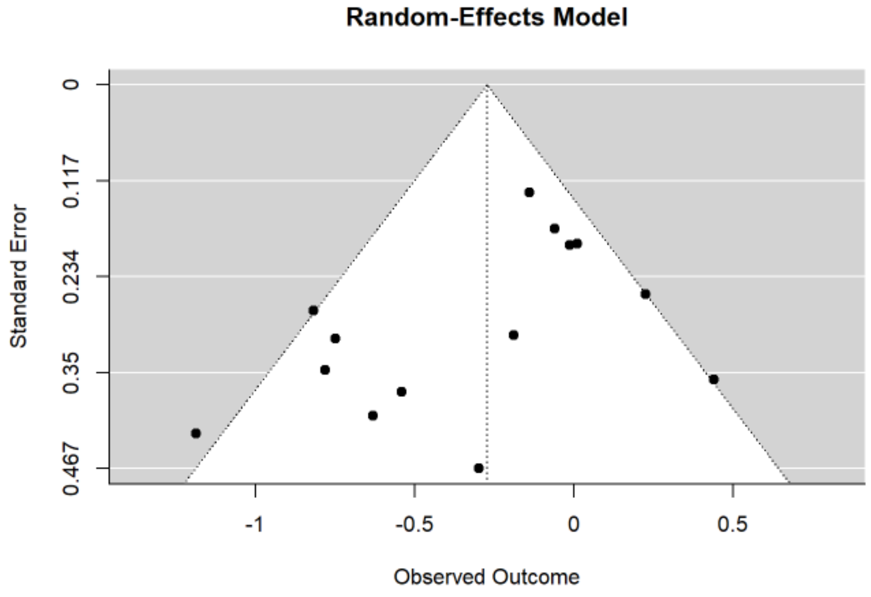

Publication bias, t = -1.182, p = 0.237

*Language – Reading*

*eFigure32. Posterior probability forest plot for reading (language)*

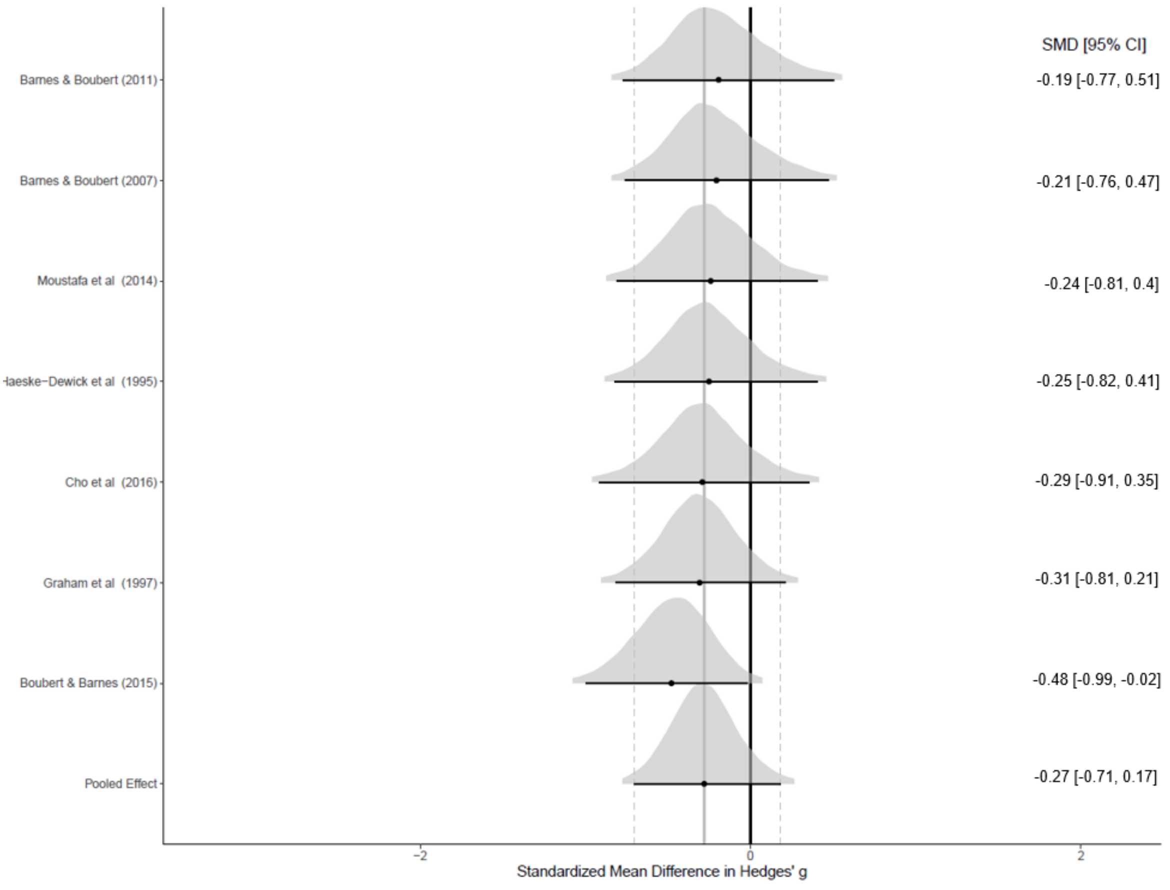

*eFigure33. Funnel plot for reading (language)*

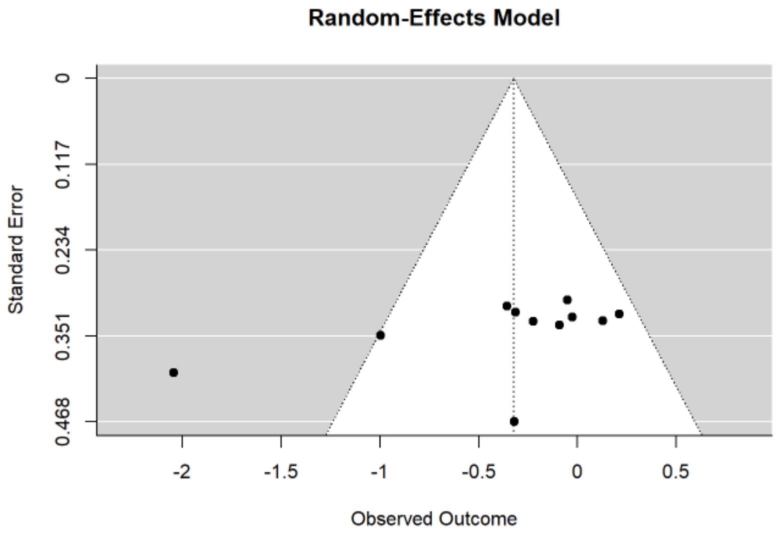

Publication bias, t = -1.787, p = 0.07

*Processing speed*

*eFigure34. Posterior probability forest plot for processing speed*

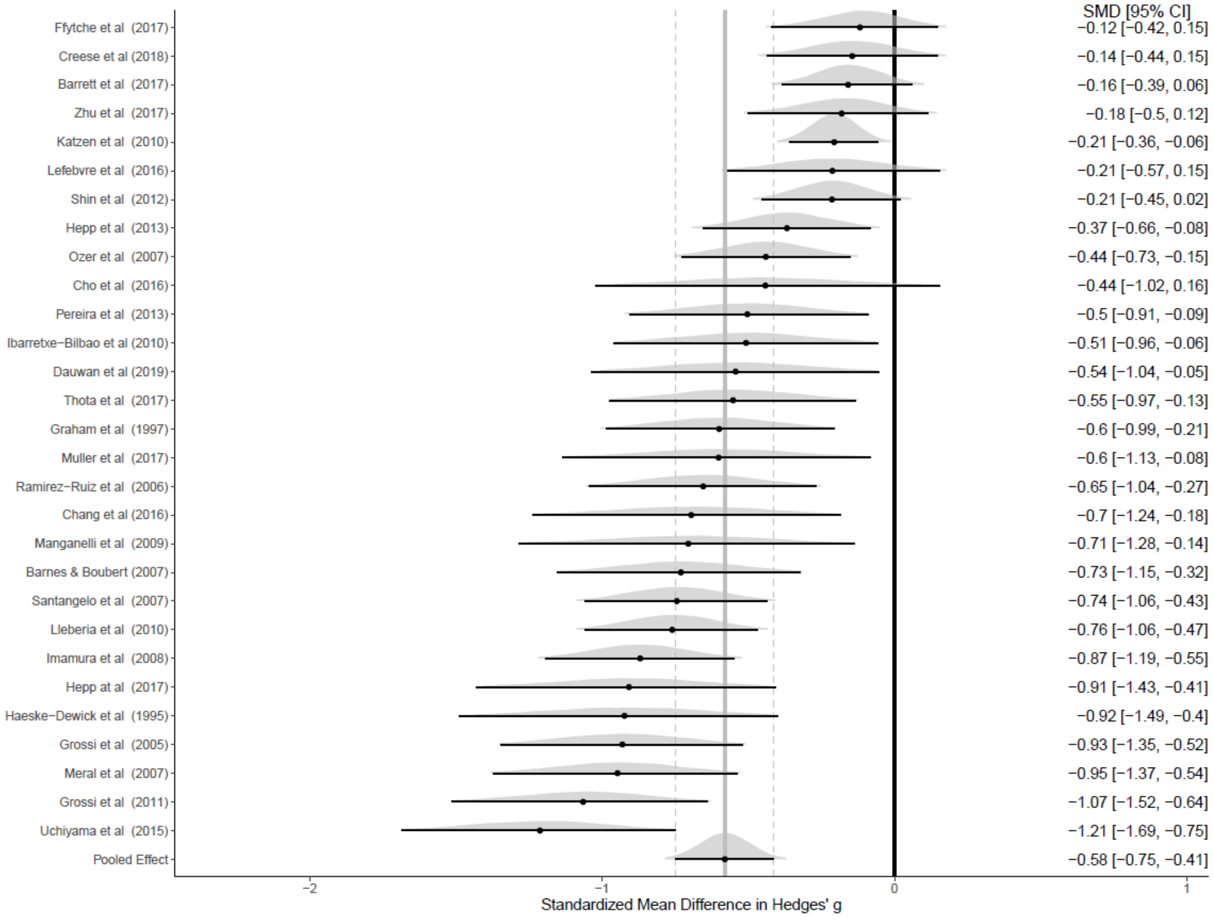

*eFigure35. Funnel plot for processing speed*

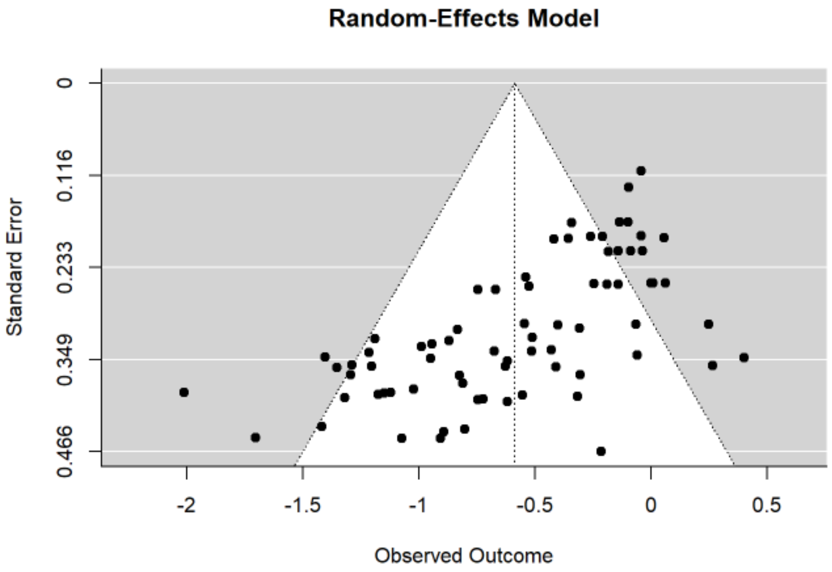

Publication bias, t = -4.470, p < 0.001

*Attention*

*eFigure36. Posterior probability forest plot for attention*

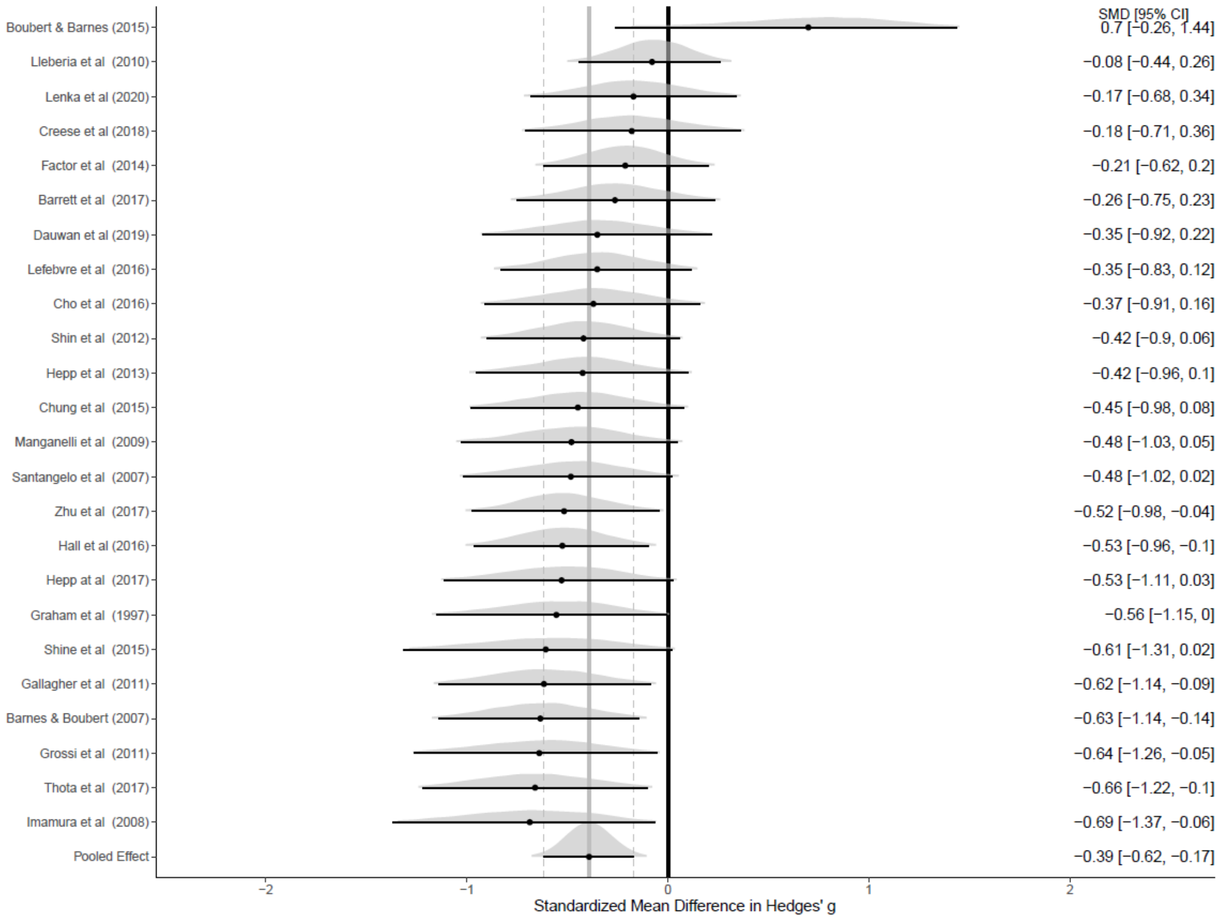

*eFigure37. Funnel plot for attention*

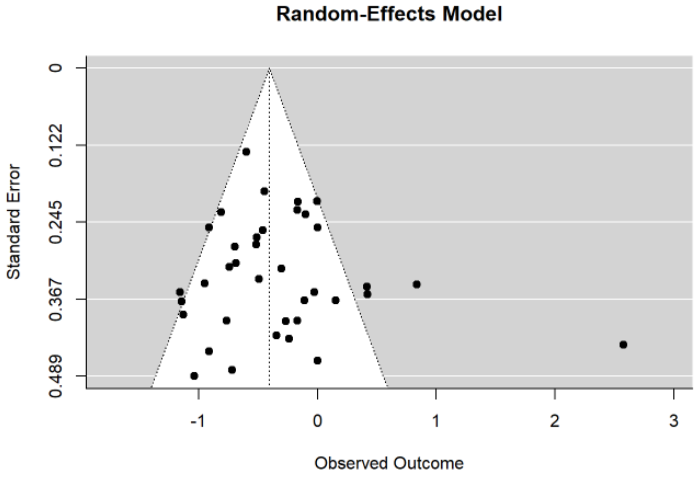

Publication bias, t = 0.353, p = 0.724

*Perception*

*eFigure38. Posterior probability forest plot for perception*

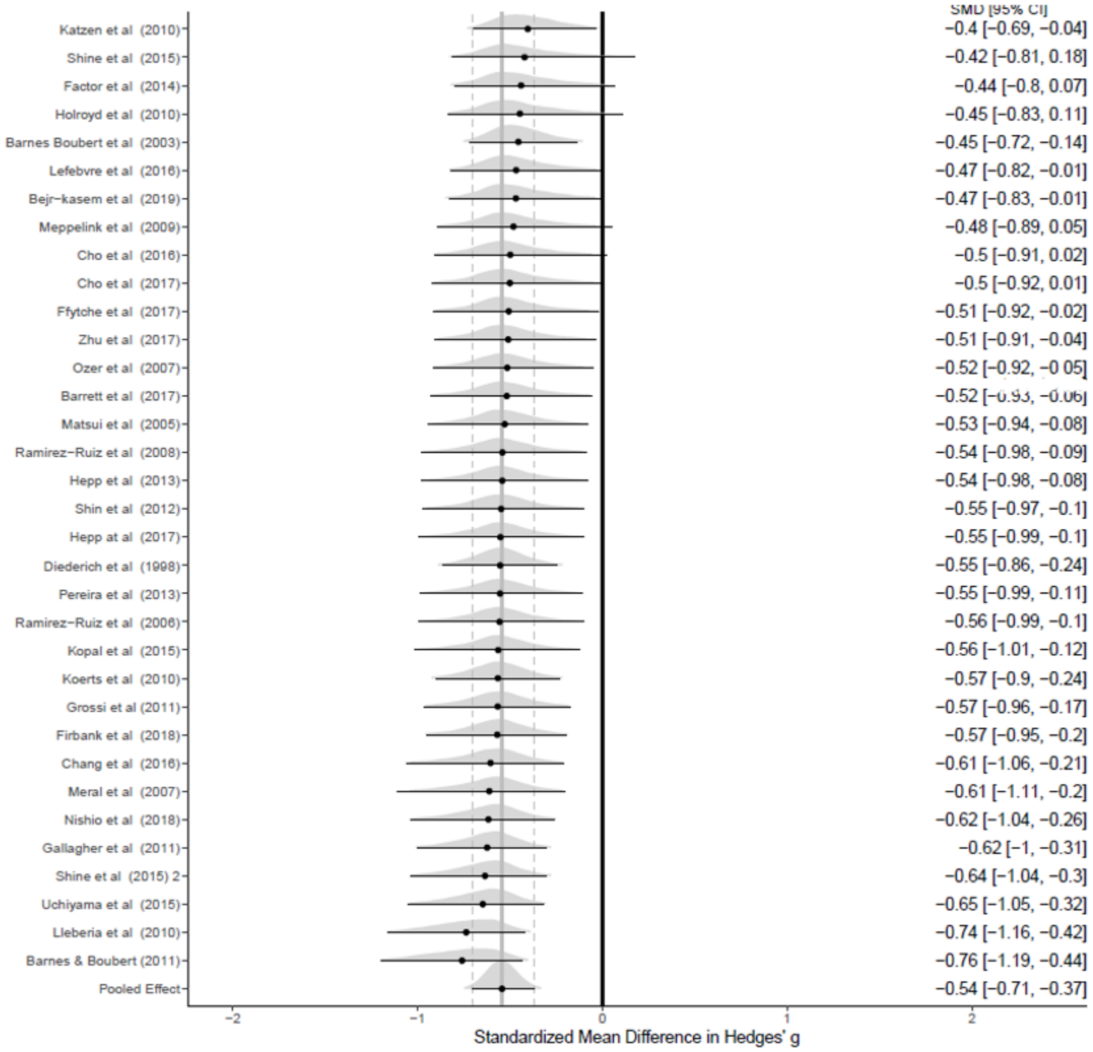

*eFigure39. Funnel plot for perception*

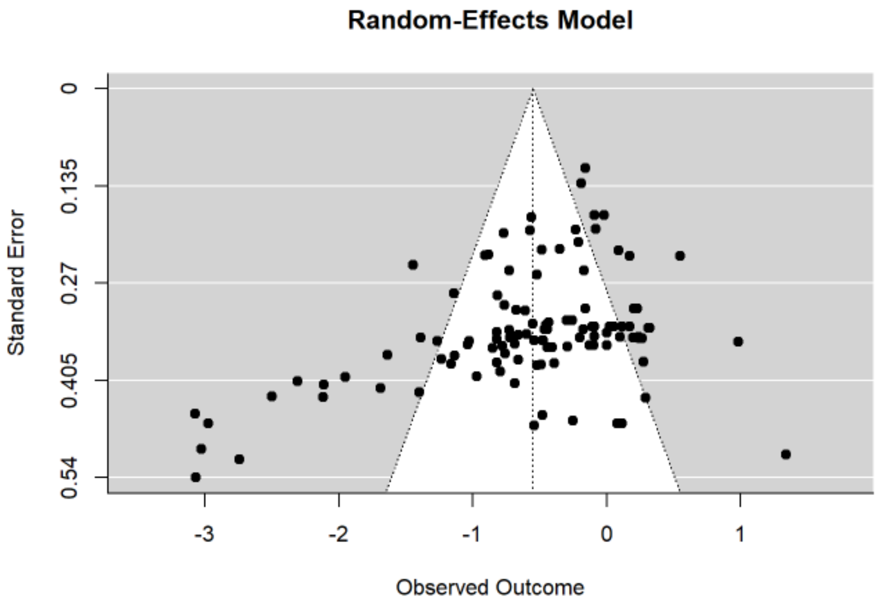

Publication bias, t = -13.191, p = 0

*Perception sub-domains*

*eFigure40. Posterior probability forest plot for perception sub-domains*

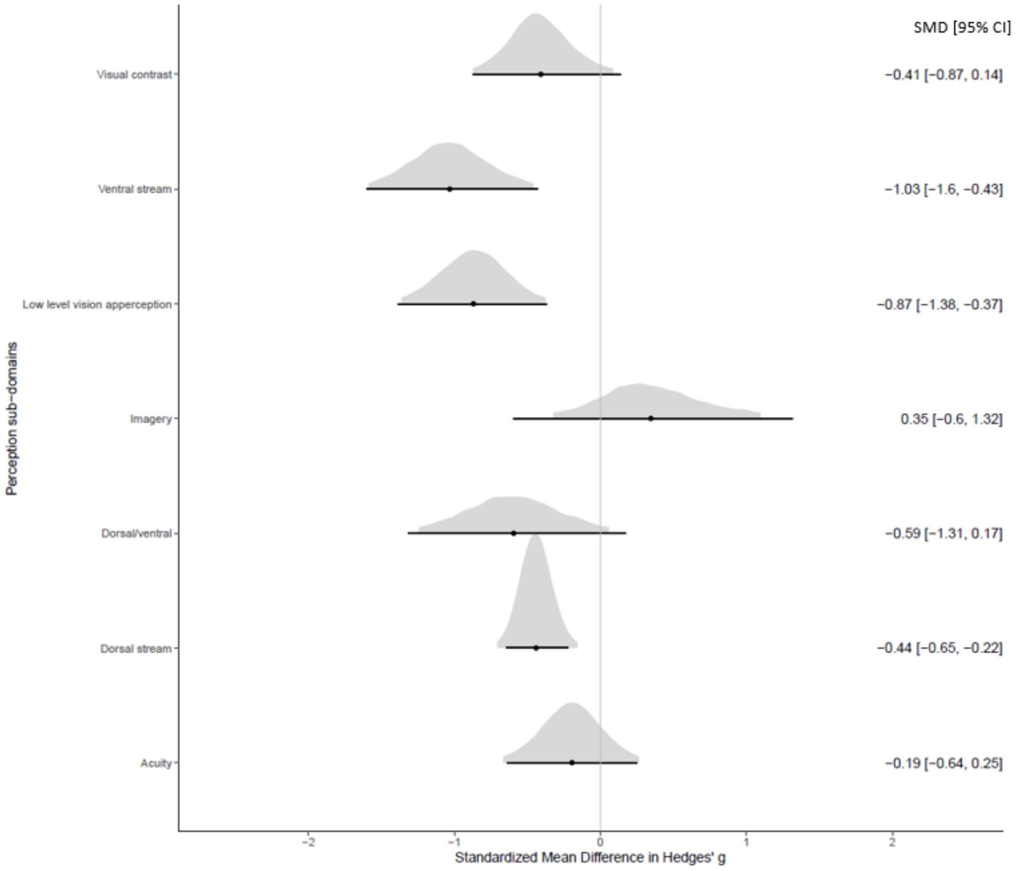

*eFigure41. Funnel plot for perception sub-domains. A) visual acuity, B) Dorsal stream, C) Ventral stream, D) Dorsal/Ventral stream, E) Low level vision apperception*

*A Visual acuity:*

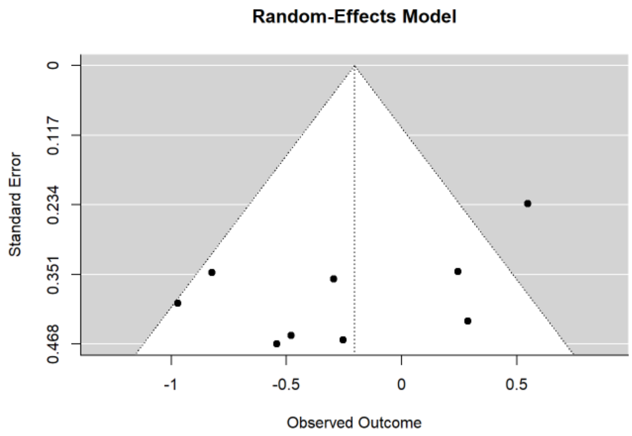

Publication bias, t = -1.113, p = 0.266

*B Dorsal stream:*

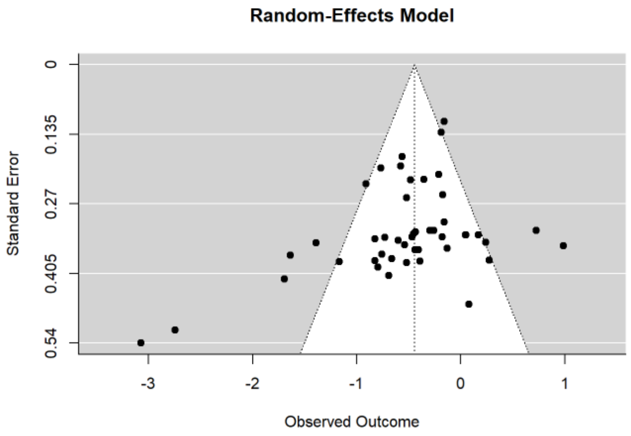

Publication bias, t = -6.224, p < 0.001

*C Ventral stream:*

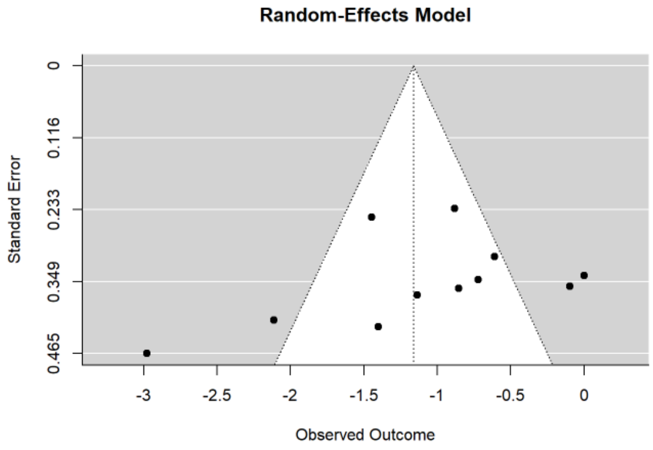

Publication bias, t = -2.277, p = 0.022

*D Dorsal stream/ventral stream:*

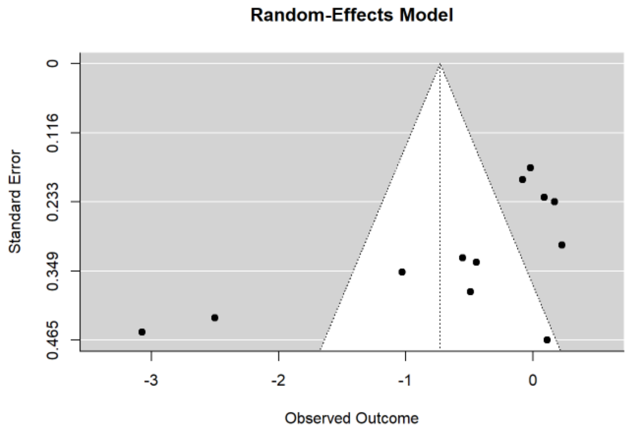

Publication bias, t = -2.349, p = 0.019

*E Low level vision apperception:*

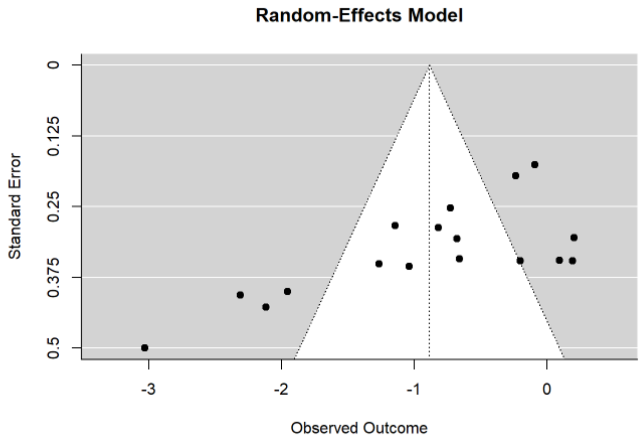

Publication bias, t = -6.141, p < 0.001

*Executive functions*

*eFigure42. Posterior probability forest plot for executive functions*

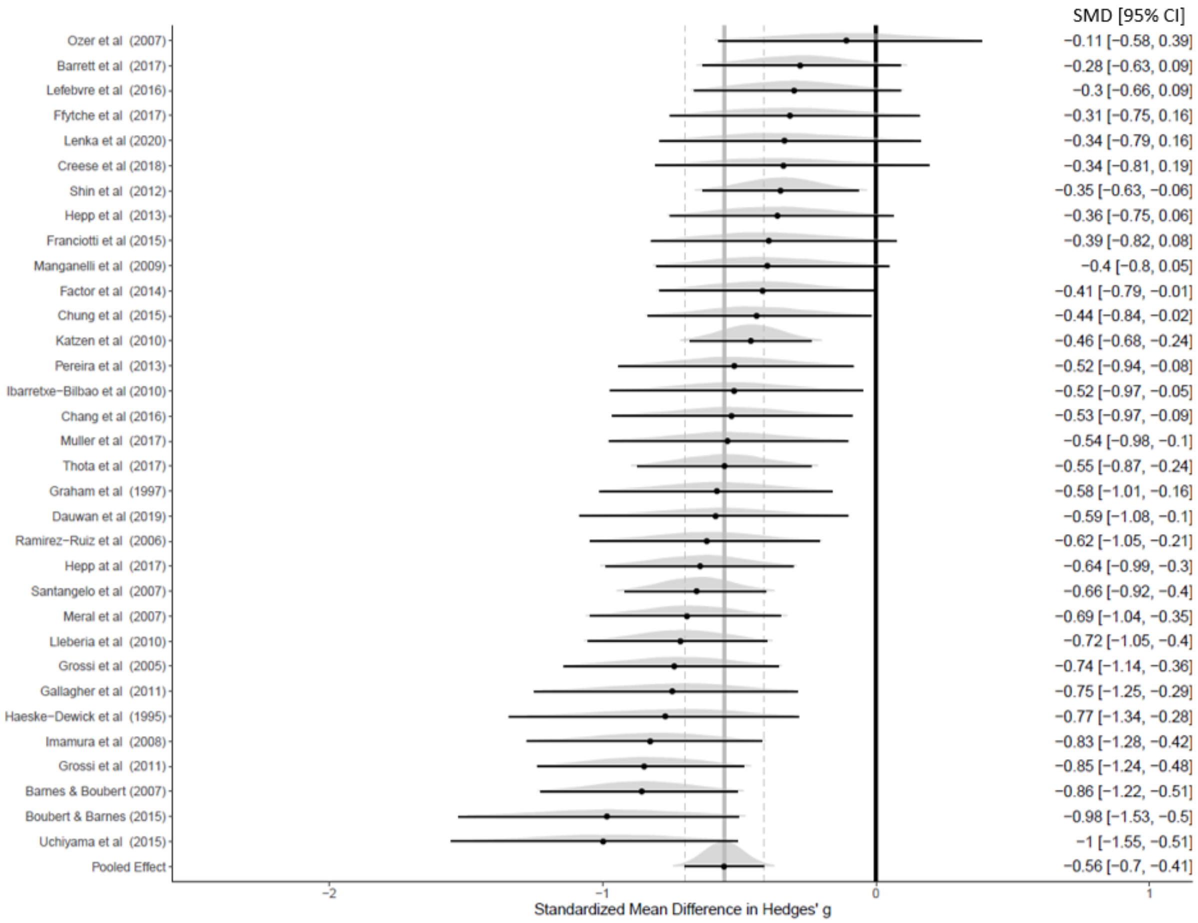

*eFigure43. Funnel plot for executive functions*

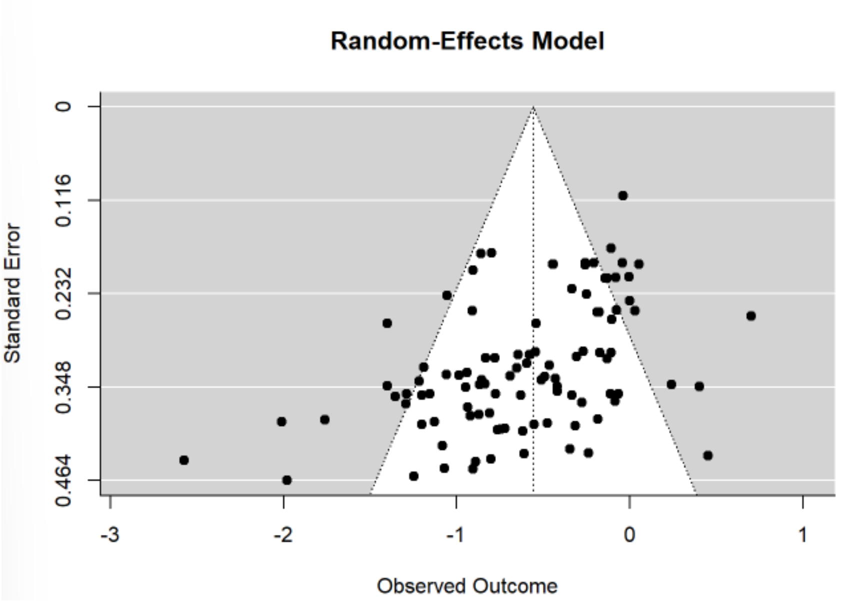

Publication bias, t = -6.132, p <0.001

### Supplementary Material 5

We conducted additional analyses to examine whether there were significant differences in main socio-demographic and clinical variables in the patients included in the same sample analysis (i.e., k=4 studies) and across all studies (k=101 studies) (PDP, n = 124, mean ± SD age = 71.15 ± 5.86 years, mean ± SD PD duration = 7.11 ± 3.53 years, mean ± SD motor scores = 25.62 ± 3.38, and PDnP, n = 139, mean ± SD age = 67.77 ± 4.70 years, mean ± SD PD duration = 6.03 ± 3.87 years, mean ± SD motor scores = 24.03 ± 2.70). There were no differences in age, PD duration, PD medications (expressed in LEDD mg/day), and motor symptoms (measured with the UPDRS part III) between these two study groups (p > 0.05).

| | **All studies (k=101)** | | **Same sample studies (k=4)** | |
|---|---|---|---|---|
| | PDP (n=2788) | PDnP (n=5931) | PDP (n=124) | PDnP (n=139) |
| Age (mean ± SD)[a] | 68.781 ± 4.231 | 66.873 ± 4.220 | 71.15 ± 5.857 | 69.415 ± 4.444 |
| UPDRS part III (mean ± SD)[b] | 28.274 ± 7.990 | 23.120 ± 6.835 | 25.617 ± 3.3784 | 24.918 ± 2.505 |
| LEDD mg/day (mean ± SD)[c] | 638.182 ± 225.394 | 564.192 ± 219.519 | 751.12 ± 206.636 | 705.892 ± 124.087 |
| PD duration (years) (mean ± SD)[d] | 9.184 ± 2.881 | 7.189 ± 2.555 | 7.106 ± 3.529 | 6.656 ± 3.147 |

[a] PDP (all studies) vs. PDP (same sample – all domains), t=-1.304, *p*=0.195. PD (all studies) vs PD (same sample – all domains), t=-1.426, *p*=0.156

[b] PDP (all studies) vs. PDP (same sample – all domains), t=0.806, *p*=0.422. PD (all studies) vs PD (same sample – all domains), t=-0.642, *p*=0.522

[c] PDP (all studies) vs. PDP (same sample – all domains), t=1.195, *p*=0.235. PD (all studies) vs PD (same sample – all domains), t=-1.560, *p* =0.122

[d] PDP (all studies) vs. PDP (same sample – all domains), t=1.694, *p*=0.093. PD (all studies) vs PD (same sample – all domains), t=0.490, *p*=0.625

### PRISMA checklist

| Section and Topic | Item # | Checklist item | Location where item is reported |
|---|---|---|---|
| **TITLE** | | | |
| Title | 1 | Identify the report as a systematic review. | Title page (page 1) |
| **ABSTRACT** | | | |
| Abstract | 2 | See the PRISMA 2020 for Abstracts checklist. | Page 3 |
| **INTRODUCTION** | | | |
| Rationale | 3 | Describe the rationale for the review in the context of existing knowledge. | Page 4 |
| Objectives | 4 | Provide an explicit statement of the objective(s) or question(s) the review addresses. | Page 4 |
| **METHODS** | | | |
| Eligibility criteria | 5 | Specify the inclusion and exclusion criteria for the review and how studies were grouped for the syntheses. | Search strategy section (page 5) |
| Information sources | 6 | Specify all databases, registers, websites, organisations, reference lists and other sources searched or consulted to identify studies. Specify the date when each source was last searched or consulted. | Search strategy section (page 5) |
| Search strategy | 7 | Present the full search strategies for all databases, registers and websites, including any filters and limits used. | Search strategy section (page 5) and supplementary material 1 |
| Selection process | 8 | Specify the methods used to decide whether a study met the inclusion criteria of the review, including how many reviewers screened each record and each report retrieved, whether they worked independently, and if applicable, details of automation tools used in the process. | Eligibility criteria (page 5-6) and PRISMA flow chart |
| Data collection process | 9 | Specify the methods used to collect data from reports, including how many reviewers collected data from each report, whether they worked independently, any processes for obtaining or confirming data from study investigators, and if applicable, details of automation tools used in the process. | Data extraction – supplementary material 1 |

| Section and Topic | Item # | Checklist item | Location where item is reported |
| --- | --- | --- | --- |
| Data items | 10a | List and define all outcomes for which data were sought. Specify whether all results that were compatible with each outcome domain in each study were sought (e.g. for all measures, time points, analyses), and if not, the methods used to decide which results to collect. | Data extraction – supplementary material 1 |
| | 10b | List and define all other variables for which data were sought (e.g. participant and intervention characteristics, funding sources). Describe any assumptions made about any missing or unclear information. | Data extraction – supplementary material 1 |
| Study risk of bias assessment | 11 | Specify the methods used to assess risk of bias in the included studies, including details of the tool(s) used, how many reviewers assessed each study and whether they worked independently, and if applicable, details of automation tools used in the process. | Assessment of study quality – supplementary material 1 |
| Effect measures | 12 | Specify for each outcome the effect measure(s) (e.g. risk ratio, mean difference) used in the synthesis or presentation of results. | Data synthesis (page 6-7) |
| Synthesis methods | 13a | Describe the processes used to decide which studies were eligible for each synthesis (e.g. tabulating the study intervention characteristics and comparing against the planned groups for each synthesis (item #5)). | Data synthesis (page 6-7) |
| | 13b | Describe any methods required to prepare the data for presentation or synthesis, such as handling of missing summary statistics, or data conversions. | Data synthesis (page 6-7) |
| | 13c | Describe any methods used to tabulate or visually display results of individual studies and syntheses. | Data synthesis (page 6-7) |
| | 13d | Describe any methods used to synthesize results and provide a rationale for the choice(s). If meta-analysis was performed, describe the model(s), method(s) to identify the presence and extent of statistical heterogeneity, and software package(s) used. | Data synthesis (page 6-7) |
| | 13e | Describe any methods used to explore possible causes of heterogeneity among study results (e.g. subgroup analysis, meta-regression). | Data synthesis (page 6-7) |
| | 13f | Describe any sensitivity analyses conducted to assess robustness of the synthesized results. | Data synthesis (page 6-7) |
| Reporting bias assessment | 14 | Describe any methods used to assess risk of bias due to missing results in a synthesis (arising from reporting biases). | Data synthesis (page 6-7) |
| Certainty assessment | 15 | Describe any methods used to assess certainty (or confidence) in the body of evidence for an outcome. | Data synthesis (page 6-7) |
| **RESULTS** | | | |
| Study selection | 16a | Describe the results of the search and selection process, from the number of records identified in the search to the number of studies included in the review, ideally using a flow diagram. | PRISMA flow chart and Results (page |

| Section and Topic | Item # | Checklist item | Location where item is reported |
|---|---|---|---|
| | | | 8-9) |
| | 16b | Cite studies that might appear to meet the inclusion criteria, but which were excluded, and explain why they were excluded. | PRISMA flowchart (page 9) |
| Study characteristics | 17 | Cite each included study and present its characteristics. | Table 1 (page 10), and study characteristics (supplementary material 2) |
| Risk of bias in studies | 18 | Present assessments of risk of bias for each included study. | Results (page 11), and supplementary material 4 |
| Results of individual studies | 19 | For all outcomes, present, for each study: (a) summary statistics for each group (where appropriate) and (b) an effect estimate and its precision (e.g. confidence/credible interval), ideally using structured tables or plots. | Results Table 1 (page 10) |
| Results of syntheses | 20a | For each synthesis, briefly summarise the characteristics and risk of bias among contributing studies. | Results (page 11), and supplementary material 4 |
| | 20b | Present results of all statistical syntheses conducted. If meta-analysis was done, present for each the summary estimate and its precision (e.g. confidence/credible interval) and measures of statistical heterogeneity. If comparing groups, describe the direction of the effect. | Results (page 10-16), table 2 (page 14-15), and figure 2 (page 9) |
| | 20c | Present results of all investigations of possible causes of heterogeneity among study results. | Results (page 10-16), table 2 (page 14-15), and supplementary material 4 |
| | 20d | Present results of all sensitivity analyses conducted to assess the robustness of the synthesized results. | Results (page 10-16), table 2 (page 14-15), and supplementary material 4-5 |

| Section and Topic | Item # | Checklist item | Location where item is reported |
|---|---|---|---|
| Reporting biases | 21 | Present assessments of risk of bias due to missing results (arising from reporting biases) for each synthesis assessed. | Supplementary material 4 |
| Certainty of evidence | 22 | Present assessments of certainty (or confidence) in the body of evidence for each outcome assessed. | Results (page 10-16), table 2 (page 14-15), and supplementary material 4-5 |
| **DISCUSSION** | | | |
| Discussion | 23a | Provide a general interpretation of the results in the context of other evidence. | Discussion (page 17) |
| | 23b | Discuss any limitations of the evidence included in the review. | Discussion (page 18-20) |
| | 23c | Discuss any limitations of the review processes used. | Discussion (page 18-20) |
| | 23d | Discuss implications of the results for practice, policy, and future research. | Discussion (page 18-20) |
| **OTHER INFORMATION** | | | |
| Registration and protocol | 24a | Provide registration information for the review, including register name and registration number, or state that the review was not registered. | Search strategy section (page 5) |
| | 24b | Indicate where the review protocol can be accessed, or state that a protocol was not prepared. | Search strategy section (page 5) |
| | 24c | Describe and explain any amendments to information provided at registration or in the protocol. | Search strategy section (page 5) |
| Support | 25 | Describe sources of financial or non-financial support for the review, and the role of the funders or sponsors in the review. | Page 2 |
| Competing interests | 26 | Declare any competing interests of review authors. | Page 2 |
| Availability of | 27 | Report which of the following are publicly available and where they can be found: template data collection forms; data extracted from included | Page 2 |

| Section and Topic | Item # | Checklist item | Location where item is reported |
|---|---|---|---|
| data, code and other materials | | studies; data used for all analyses; analytic code; any other materials used in the review. | |

### References

1. Luo D, Wan X, Liu J, Tong T. Optimally estimating the sample mean from the sample size, median, mid-range, and/or mid-quartile range. Stat Methods Med Res 2018;27(6):1785-1805.
2. Wan X, Wang W, Liu J, Tong T. Estimating the sample mean and standard deviation from the sample size, median, range and/or interquartile range. BMC Med Res Methodol 2014;14:135.
3. Goetz CG, Fahn S, Martinez-Martin P, et al. Movement Disorder Society-sponsored revision of the Unified Parkinson's Disease Rating Scale (MDS-UPDRS): Process, format, and clinimetric testing plan. Movement Disorders 2007;22(1):41-47.

*Articles included in the review (k=105)*

4. Aarsland D, Larsen JP, Cummings JL, Laake K. Prevalence and clinical correlates of psychotic symptoms in Parkinson disease - A community-based study. Archives of Neurology 1999;56(5):595-601.
5. Ballanger B, Strafella AP, van Eimeren T, et al. Serotonin 2A receptors and visual hallucinations in Parkinson disease. Arch Neurol 2010;67(4):416-421.
6. Barnes J, Boubert L. Executive functions are impaired in patients with Parkinson's disease with visual hallucinations. Journal of Neurology Neurosurgery and Psychiatry 2008;79(2):190-192.
7. Barnes J, Boubert L. Visual Memory Errors in Parkinson's Disease Patients With Visual Hallucinations. International Journal of Neuroscience 2011;121(3):159-164.
8. Barnes J, David A. Visual hallucinations in Parkinson's disease: a review and phenomenological survey. Journal of Neurology, Neurosurgery & Psychiatry 2001;70(6):727-733.
9. Barnes J, Boubert L, Harris J, Lee A, David AS. Reality monitoring and visual hallucinations in Parkinson's disease. Neuropsychologia 2003;41(5):565-574.
10. Barrett MJ, Smolkin ME, Flanigan JL, Shah BB, Harrison MB, Sperling SA. Characteristics, correlates, and assessment of psychosis in Parkinson disease without dementia. Parkinsonism Relat Disord 2017;43:56-60.
11. Bejr-kasem H, Pagonabarraga J, Martínez-Horta S, et al. Disruption of the default mode network and its intrinsic functional connectivity underlies minor hallucinations in Parkinson's disease. Movement Disorders 2019;34(1):78-86.
12. Boecker H, Ceballos-Baumann AO, Volk D, Conrad B, Forstl H, Haussermann P. Metabolic alterations in patients with Parkinson disease and visual hallucinations. Archives of Neurology 2007;64(7):984-988.
13. Boubert L, Barnes J. Phenomenology of Visual Hallucinations and Their Relationship to Cognitive Profile in Parkinson's Disease Patients: Preliminary Observations. Sage Open 2015;5(2).
14. Chang YP, Yang YH, Lai CL, Liou LM. Event-Related Potentials in Parkinson's Disease Patients with Visual Hallucination. Parkinsons Dis 2016;2016.
15. Cho SS, Strafella AP, Duff-Canning S, et al. The Relationship Between Serotonin-2A Receptor and Cognitive Functions in Nondemented Parkinson's Disease Patients with Visual Hallucinations. Movement disorders clinical practice 2017;4(5):698-709.
16. Chung EJ, Seok K, Kim SJ. A comparison of Montreal Cognitive Assessment between patients with visual hallucinations and without visual hallucinations in Parkinson's disease. Clinical Neurology and Neurosurgery 2015;130:98-100.
17. Clegg BJ, Duncan GW, Khoo TK, et al. Categorising visual hallucinations in early Parkinson's disease. Journal of Parkinson's Disease 2018;8(3):447-453.
18. Creese B, Albertyn CP, Dworkin S, Thomas RS, Wan YM, Ballard C. Executive function but not episodic memory decline associated with visual hallucinations in Parkinson's disease. J Neuropsychol 2020;14(1):85-97.
19. Dauwan M, Hoff JI, Vriens EM, Hillebrand A, Stam CJ, Sommer IE. Aberrant resting-state oscillatory brain activity in Parkinson's disease patients with visual hallucinations: An MEG source-space study. Neuroimage-Clinical 2019;22.

20. Dave S, Weintraub D, Aarsland D, Ffytche DH. Drug and Disease Effects in Parkinson's Psychosis: Revisiting the Role of Dopamine. Mov Disord Clin Pract 2020;7(1):32-36.
21. Debs R, De Cock VC, Negre-Pages L, Aristin M, Senard A, Rascol O. Thought disorders among non-demented outpatients with Parkinson's disease: prevalence and associated factors. Journal of Neural Transmission 2010;117(10):1183-1188.
22. Diederich NJ, Goetz CG, Raman R, Pappert EJ, Leurgans S, Piery V. Poor visual discrimination and visual hallucinations in Parkinson's disease. Clin Neuropharmacol 1998;21(5):289-295.
23. Doé de Maindreville A, Fénelon G, Mahieux F. Hallucinations in Parkinson's disease: a follow-up study. Movement disorders: official journal of the Movement Disorder Society 2005;20(2):212-217.
24. Factor SA, Scullin MK, Sollinger AB, et al. Cognitive correlates of hallucinations and delusions in Parkinson's disease. J Neurol Sci 2014;347(1-2):316-321.
25. Factor SA, Scullin MK, Freeman A, Bliwise DL, McDonald WM, Goldstein FC. Affective Correlates of Psychosis in Parkinson's Disease. Mov Disord Clin Pract 2017;4(2):225-230.
26. Fenelon G, Mahieux F, Huon R, Ziegler M. Hallucinations in Parkinson's disease - Prevalence, phenomenology and risk factors. Brain 2000;123:733-745.
27. Fernandez W, Stern G, Lees A. Hallucinations and parkinsonian motor fluctuations. Behavioural neurology 1992;5(2):83-86.
28. Ffytche DH, Pereira JB, Ballard C, Chaudhuri KR, Weintraub D, Aarsland D. Risk factors for early psychosis in PD: insights from the Parkinson's Progression Markers Initiative. J Neurol Neurosurg Psychiatry 2017;88(4):325-331.
29. Firbank MJ, Parikh J, Murphy N, et al. Reduced occipital GABA in Parkinson disease with visual hallucinations. Neurology 2018;91(7):e675.
30. Forsaa EB, Larsen JP, Wentzel-Larsen T, et al. A 12-year population-based study of psychosis in Parkinson disease. Arch Neurol 2010;67(8):996-1001.
31. Franciotti R, Delli Pizzi S, Perfetti B, et al. Default Mode Network Links to Visual Hallucinations: A Comparison Between Parkinson's Disease and Multiple System Atrophy. Movement Disorders 2015;30(9):1237-1247.
32. Gallagher DA, Parkkinen L, O'Sullivan SS, et al. Testing an aetiological model of visual hallucinations in Parkinson's disease. Brain 2011;134(11):3299-3309.
33. Gama RL, Bruin VM, Távora DG, Duran FL, Bittencourt L, Tufik S. Structural brain abnormalities in patients with Parkinson's disease with visual hallucinations: a comparative voxel-based analysis. Brain Cogn 2014;87:97-103.
34. Garofalo S, Justicia A, Arrondo G, et al. Cortical and Striatal Reward Processing in Parkinson's Disease Psychosis. Front Neurol 2017;8:156.
35. Gobel N, Moeller JC, Hollenstein N, et al. Face Perception and Pareidolia Production in Patients With Parkinson's Disease. Frontiers in Neurology 2021;12.
36. Goetz CG, Ouyang B, Negron A, Stebbins GT. Hallucinations in PD: Ten Year Prospective Longitudinal Assessment. Neurology 2010;74(9):A73-A73.
37. Goetz CG, Wuu J, Curgian L, Leurgans S. Age-related influences on the clinical characteristics of new-onset hallucinations in Parkinson's disease patients. Movement disorders 2006;21(2):267-270.
38. Goldman JG, Stebbins GT, Dinh V, et al. Visuoperceptive region atrophy independent of cognitive status in patients with Parkinson’s disease with hallucinations. Brain 2014;137(3):849-859.
39. Gordon PC, Kauark RB, Costa CD, de Oliveira MO, Godinho FL, Rocha MS. Clinical Implications of the National Institute of Neurological Disorders and Stroke Criteria for Diagnosing Psychosis in Parkinson's Disease. J Neuropsychiatry Clin Neurosci 2016;28(1):26-31.
40. Graham JM, Grunewald RA, Sagar HJ. Hallucinosis in idiopathic Parkinson's disease. Journal of Neurology Neurosurgery and Psychiatry 1997;63(4):434-440.
41. Grossi D, Trojano L, Pellecchia MT, Amboni M, Fragassi NA, Barone P. Frontal dysfunction contributes to the genesis of hallucinations in non-demented Parkinsonian patients. International

Journal of Geriatric Psychiatry: A journal of the psychiatry of late life and allied sciences 2005;20(7):668-673.
42. Grossi D, Carotenuto A, Trojano L, Manzo V, Fasanaro AM. Do frontal dysfunctions play a role in visual hallucinations in Alzheimer's disease as in Parkinson's disease? a comparative study. Psychology & Neuroscience 2011;4:385-389.
43. Haeske-Dewick HC. Hallucinations in Parkinson's disease: Characteristics and associated clinical features. International Journal of Geriatric Psychiatry 1995;10(6):487-495.
44. Hall JM, O'Callaghan C, Shine JM, et al. Dysfunction in attentional processing in patients with Parkinson's disease and visual hallucinations. Journal of Neural Transmission 2016;123(5):503-507.
45. Hepp DH, Foncke EMJ, Berendse HW, et al. Damaged fiber tracts of the nucleus basalis of Meynert in Parkinson's disease patients with visual hallucinations. Scientific Reports 2017;7.
46. Hepp DH, da Hora CC, Koene T, et al. Cognitive correlates of visual hallucinations in non-demented Parkinson's disease patients. Parkinsonism & Related Disorders 2013;19(9):795-799.
47. Holroyd S, Currie L, Wooten G. Prospective study of hallucinations and delusions in Parkinson's disease. Journal of Neurology, Neurosurgery & Psychiatry 2001;70(6):734-738.
48. Ibarretxe-Bilbao N, Ramirez-Ruiz B, Junque C, et al. Differential progression of brain atrophy in Parkinson's disease with and without visual hallucinations. Journal of Neurology Neurosurgery and Psychiatry 2010;81(6):650-657.
49. Ibarretxe-Bilbao N, Ramirez-Ruiz B, Tolosa E, et al. Hippocampal head atrophy predominance in Parkinson's disease with hallucinations and with dementia. Journal of neurology 2008;255(9):1324-1331.
50. Imamura K, Wada-Isoe K, Kitayama M, Nakashima K. Executive dysfunction in non-demented Parkinson's disease patients with hallucinations. Acta Neurologica Scandinavica 2008;117(4):255-259.
51. Jacobson SA, Morshed T, Dugger BN, et al. Plaques and tangles as well as Lewy-type alpha synucleinopathy are associated with formed visual hallucinations. Parkinsonism Relat Disord 2014;20(9):1009-1014.
52. Janzen J, van 't Ent D, Lemstra AW, Berendse HW, Barkhof F, Foncke EM. The pedunculopontine nucleus is related to visual hallucinations in Parkinson's disease: preliminary results of a voxel-based morphometry study. J Neurol 2012;259(1):147-154.
53. Katzen H, Myerson C, Papapetropoulos S, Nahab F, Gallo B, Levin B. Multi-Modal Hallucinations and Cognitive Function in Parkinson's Disease. Dement Geriatr Cogn Disord 2010;30(1):51-56.
54. Kiferle L, Ceravolo R, Petrozzi L, et al. Visual hallucinations in Parkinson's disease are not influenced by polymorphisms of serotonin 5-HT2A receptor and transporter genes. Neurosci Lett 2007;422(3):228-231.
55. Kiferle L, Ceravolo R, Giuntini M, et al. Caudate dopaminergic denervation and visual hallucinations: evidence from a $^{123}$I-FP-CIT SPECT study. Parkinsonism Relat Disord 2014;20(7):761-765.
56. Koerts J, Borg M, Meppelink AM, Leenders KL, van Beilen M, van Laar T. Attentional and perceptual impairments in Parkinson's disease with visual hallucinations. Parkinsonism & Related Disorders 2010;16(4):270-274.
57. Kopal A, Mejzlikova E, Preiningerova JL, et al. Changes of Retina Are Not Involved in the Genesis of Visual Hallucinations in Parkinson's Disease. Parkinsons Dis 2015;2015.
58. Lee AH, Weintraub D. Psychosis in Parkinson's disease without dementia: common and comorbid with other non-motor symptoms. Movement Disorders 2012;27(7):858-863.
59. Lee JY, Kim JM, Ahn J, Kim HJ, Jeon BS, Kim TW. Retinal Nerve Fiber Layer Thickness and Visual Hallucinations in Parkinson's Disease. Movement Disorders 2014;29(1):61-67.
60. Lee J-Y, Yoon EJ, Lee WW, Kim YK, Lee J-Y, Jeon B. Lateral geniculate atrophy in Parkinson's with visual hallucination: A trans-synaptic degeneration? Movement Disorders 2016;31(4):547-554.

61. Lee WW, Yoon EJ, Lee JY, Park SW, Kim YK. Visual Hallucination and Pattern of Brain Degeneration in Parkinsons Disease. Neurodegener Dis 2017;17(2-3):63-72.
62. Lefebvre S, Baille G, Jardri R, et al. Hallucinations and conscious access to visual inputs in Parkinson’s disease. Scientific Reports 2016;6(1):36284.
63. Lenka A, Ingalhalikar M, Shah A, et al. Abnormalities in the white matter tracts in patients with Parkinson disease and psychosis. Neurology 2020;94(18):E1876-E1884.
64. Lenka A, Ingalhalikar M, Shah A, et al. Hippocampal subfield atrophy in patients with Parkinson’s disease and psychosis. Journal of Neural Transmission 2018;125(9):1361-1372.
65. Leu-Semenescu S, De Cock VC, Le Masson VD, et al. Hallucinations in narcolepsy with and without cataplexy: contrasts with Parkinson’s disease. Sleep Medicine 2011;12(5):497-504.
66. Llebaria G, Pagonabarraga J, Martínez-Corral M, et al. Neuropsychological correlates of mild to severe hallucinations in Parkinson's disease. Mov Disord 2010;25(16):2785-2791.
67. Mack J, Rabins P, Anderson K, et al. Prevalence of psychotic symptoms in a community-based Parkinson disease sample. The American Journal of Geriatric Psychiatry 2012;20(2):123-132.
68. Manganelli F, Vitale C, Santangelo G, et al. Functional involvement of central cholinergic circuits and visual hallucinations in Parkinson's disease. Brain 2009;132(9):2350-2355.
69. Matsui H, Udaka F, Tamura A, et al. The relation between visual hallucinations and visual evoked potential in Parkinson disease. Clinical neuropharmacology 2005;28(2):79-82.
70. Matsui H, Udaka F, Tamura A, et al. Impaired visual acuity as a risk factor for visual hallucinations in Parkinson's disease. Journal of Geriatric Psychiatry and Neurology 2006;19(1):36-40.
71. Meppelink AM, de Jong BM, Renken R, Leenders KL, Cornelissen FW, van Laar T. Impaired visual processing preceding image recognition in Parkinson's disease patients with visual hallucinations. Brain 2009;132(Pt 11):2980-2993.
72. Meral H, Aydemir T, Ozer F, et al. Relationship between visual hallucinations and REM sleep behavior disorder in patients with Parkinson's disease. Clinical Neurology and Neurosurgery 2007;109(10):862-867.
73. Miloserdov K, Schmidt-Samoa C, Williams K, et al. Aberrant functional connectivity of resting state networks related to misperceptions and intra-individual variability in Parkinson's disease. Neuroimage-Clinical 2020;25.
74. Morgante L, Colosimo C, Antonini A, et al. Psychosis associated to Parkinson's disease in the early stages: relevance of cognitive decline and depression. Journal of Neurology, Neurosurgery & Psychiatry 2012;83(1):76-82.
75. Moustafa AA, Krishna R, Frank MJ, Eissa AM, Hewedi DH. Cognitive correlates of psychosis in patients with Parkinson's disease. Cognitive Neuropsychiatry 2014;19(5):381-398.
76. Muller AJ, O'Callaghan C, Walton CC, Shine JM, Lewis SJ. Retrospective Neuropsychological Profile of Patients With Parkinson Disease Prior to Developing Visual Hallucinations. J Geriatr Psychiatry Neurol 2017;30(2):90-95.
77. Nagano-Saito A, Washimi Y, Arahata Y, et al. Visual hallucination in Parkinson's disease with FDG PET. Movement disorders 2004;19(7):801-806.
78. Nishio Y, Yokoi K, Hirayama K, et al. Defining visual illusions in Parkinson's disease: Kinetopsia and object misidentification illusions. Parkinsonism and Related Disorders 2018;55:111-116.
79. Oka H, Yoshioka M, Onouchi K, et al. Impaired cardiovascular autonomic function in Parkinson's disease with visual hallucinations. Movement Disorders: Official Journal of the Movement Disorder Society 2007;22(10):1510-1514.
80. Ozer F, Meral H, Hanoglu L, et al. Cognitive impairment patterns in Parkinson's disease with visual hallucinations. J Clin Neurosci 2007;14(8):742-746.
81. Papapetropoulos S, Argyriou AA, Ellul J. Factors associated with drug-induced visual hallucinations in Parkinson's disease. J Neurol 2005;252(10):1223-1228.

82. Papapetropoulos S, Katzen H, Schrag A, et al. A questionnaire-based (UM-PDHQ) study of hallucinations in Parkinson's disease. BMC neurology 2008;8(1):1-8.
83. Park HK, Kim JS, Im KC, et al. Visual hallucinations and cognitive impairment in parkinson's disease. Canadian Journal of Neurological Sciences 2013;40(5):657-662.
84. Pereira JB, Junqué C, Bartrés-Faz D, Ramírez-Ruiz B, Marti MJ, Tolosa E. Regional vulnerability of hippocampal subfields and memory deficits in Parkinson's disease. Hippocampus 2013;23(8):720-728.
85. Porter B, Henry SR, Gray WK, Walker RW. Care requirements of a prevalent population of people with idiopathic Parkinson's disease. Age and ageing 2010;39(1):57-61.
86. Ramírez-Ruiz B, Junqué C, Martí MJ, Valldeoriola F, Tolosa E. Neuropsychological deficits in Parkinson's disease patients with visual hallucinations. Mov Disord 2006;21(9):1483-1487.
87. Ramirez-Ruiz B, Marti MJ, Tolosa E, et al. Cerebral atrophy in Parkinson's disease patients with visual hallucinations. European Journal of Neurology 2007;14(7):750-756.
88. Ramírez-Ruiz B, Martí MJ, Tolosa E, et al. Brain response to complex visual stimuli in Parkinson's patients with hallucinations: a functional magnetic resonance imaging study. Movement disorders: official journal of the Movement Disorder Society 2008;23(16):2335-2343.
89. SanchezRamos JR, Ortoll R, Paulson GW. Visual hallucinations associated with Parkinson disease. Archives of Neurology 1996;53(12):1265-1268.
90. Santangelo G, Trojano L, Vitale C, et al. A neuropsychological longitudinal study in Parkinson's patients with and without hallucinations. Movement Disorders 2007;22(16):2418-2425.
91. Sawada H, Oeda T, Umemura A, et al. Subclinical elevation of plasma C-reactive protein and illusions/hallucinations in subjects with Parkinson's disease: case-control study. PLoS One 2014;9(1):e85886.
92. Sawczak CM, Barnett AJ, Cohn M. Increased Cortical Thickness in Attentional Networks in Parkinson's Disease with Minor Hallucinations. Parkinsons Dis 2019;2019:5351749.
93. Schumacher-Schuh AF, Francisconi C, Altmann V, et al. Polymorphisms in the dopamine transporter gene are associated with visual hallucinations and levodopa equivalent dose in Brazilians with Parkinson's disease. International Journal of Neuropsychopharmacology 2013;16(6):1251-1258.
94. Schumacher-Schuh A, Altmann V, Rieck M, et al. Association of common genetic variants of HOMER1 gene with levodopa adverse effects in Parkinson’s disease patients. The Pharmacogenomics Journal 2014;14(3):289-294.
95. Shin S, Lee JE, Hong JY, Sunwoo MK, Sohn YH, Lee PH. Neuroanatomical substrates of visual hallucinations in patients with non-demented Parkinson's disease. J Neurol Neurosurg Psychiatry 2012;83(12):1155-1161.
96. Shine JM, Halliday GH, Carlos M, Naismith SL, Lewis SJ. Investigating visual misperceptions in Parkinson's disease: a novel behavioral paradigm. Mov Disord 2012;27(4):500-505.
97. Shine JM, Halliday GM, Gilat M, et al. The role of dysfunctional attentional control networks in visual misperceptions in Parkinson's disease. Human Brain Mapping 2014;35(5):2206-2219.
98. Shine JM, Keogh R, O'Callaghan C, Muller AJ, Lewis SJ, Pearson J. Imagine that: elevated sensory strength of mental imagery in individuals with Parkinson's disease and visual hallucinations. Proceedings of the Royal Society B: Biological Sciences 2015;282(1798):20142047.
99. Shine JM, Mills JMZ, Qiu J, et al. Validation of the Psychosis and Hallucinations Questionnaire in Non-demented Patients with Parkinson's Disease. Mov Disord Clin Pract 2015;2(2):175-181.
100. Shine JM, Muller AJ, O’Callaghan C, Hornberger M, Halliday GM, Lewis SJ. Abnormal connectivity between the default mode and the visual system underlies the manifestation of visual hallucinations in Parkinson’s disease: a task-based fMRI study. npj Parkinson's Disease 2015;1(1):1-8.
101. Stebbins G, Goetz C, Carrillo M, et al. Altered cortical visual processing in PD with hallucinations: an fMRI study. Neurology 2004;63(8):1409-1416.
102. Straughan S, Collerton D, Bruce V. Visual Priming and Visual Hallucinations in Parkinson's Disease. Evidence for Normal Top-Down Processes. Journal of Geriatric Psychiatry and Neurology 2016;29(1):25-30.

103. Thota N, Lenka A, George L, et al. Impaired frontal lobe functions in patients with Parkinson's disease and psychosis. Asian J Psychiatr 2017;30:192-195.
104. Uchiyama M, Nishio Y, Yokoi K, Hosokai Y, Takeda A, Mori E. Pareidolia in Parkinson's disease without dementia: A positron emission tomography study. Parkinsonism Relat Disord 2015;21(6):603-609.
105. Weintraub D, Morales KH, Duda JE, Moberg PJ, Stern MB. Frequency and correlates of co-morbid psychosis and depression in Parkinson's disease. Parkinsonism Relat Disord 2006;12(7):427-431.
106. Yao N, Cheung C, Pang S, et al. Multimodal MRI of the hippocampus in Parkinson's disease with visual hallucinations. Brain Struct Funct 2016;221(1):287-300.
107. Zarkali A, McColgan P, Ryten M, et al. Differences in network controllability and regional gene expression underlie hallucinations in Parkinson's disease. Brain : a journal of neurology 2020;29.
108. Zhu J, Shen B, Lu L, et al. Prevalence and risk factors for visual hallucinations in Chinese patients with Parkinson's disease. Journal of the Neurological Sciences 2017;372:471-476.
